# Association between Fuchs Endothelial Corneal Dystrophy, Diabetes Mellitus and Multimorbidity

**DOI:** 10.1101/2022.12.14.22283472

**Authors:** Cari L. Nealon, Christopher W. Halladay, Bryan R. Gorman, Piana Simpson, David P. Roncone, Rachael L. Canania, Scott A. Anthony, Lea R. Sawicki Rogers, Jenna N. Leber, Jacquelyn M. Dougherty, Jessica N. Cooke Bailey, Dana C. Crawford, Jack M. Sullivan, Anat Galor, Wen-Chih Wu, Paul B. Greenberg, the Million Veteran Program, Jonathan H. Lass, Sudha K. Iyengar, Neal S. Peachey

**Author notes:** Correspondence: Neal S. Peachey, Research Service (151W), VA Northeast Ohio Healthcare System, 10701 East Boulevard, Cleveland OH 44106, USA; Sudha K. Iyengar, Case Western Reserve University School of Medicine, 2103 Cornell Road, Cleveland, OH 44106, USA. These authors contributed equally to this work (Cari L. Nealon, Christopher W. Halladay, Bryan R. Gorman). These authors jointly supervised this work (Sudha K. Iyengar, Neal S. Peachey).

## Abstract

**Purpose:** To assess risk for demographic variables and other health conditions that are associated with Fuchs endothelial corneal dystrophy (FECD).

**Methods:** We developed a FECD case-control algorithm based on structured EHR data and accuracy confirmed by individual review of charts at three VA Medical Centers. This algorithm was applied to the Department of Veterans Affairs Million Veteran Program cohort from whom sex, genetic ancestry, comorbidities, diagnostic phecodes and laboratory values were extracted. Single and multiple variable logistic regression models were used to determine the association of these risk factors with FECD diagnosis.

**Results:** Being a FECD case was associated with female sex, European genetic ancestry, and a greater number of comorbidities. Of 1417 diagnostic phecodes evaluated, 213 had a significant association with FECD, falling in both ocular and non-ocular conditions, including diabetes mellitus (DM). Five of 69 laboratory values were associated with FECD, with the direction of change for four being consistent with DM. Insulin dependency and type 1 DM raised risk to a greater degree than type 2 DM, like other microvascular diabetic complications.

**Conclusions:** Female gender, European ancestry and multimorbidity increased FECD risk. Endocrine/metabolic clinic encounter codes as well as altered patterns of laboratory values support DM increasing FECD risk. Our results evoke a threshold model in which the FECD phenotype is intensified by DM and potentially other health conditions that alter corneal physiology. DM may modify FECD onset and encourage progression among susceptible individuals, suggesting that optimizing glucose control may be an effective preventative for FECD.

---

Longitudinal electronic health records (EHRs) represent an amazing opportunity to identify previously unappreciated or unrecognized relationships between complex conditions. In the present study, we focused on Fuchs endothelial corneal dystrophy (FECD) a common corneal dystrophy and a leading indication for corneal transplantation in the United States (US).^1-4^ FECD is an important healthcare challenge with an estimated prevalence in the adult population of 4%.^5^ FECD is characterized by bilateral endothelial cell pleomorphism and polymegathism, decreased endothelial cell density, guttae, and corneal edema.^1,6^ Two subtypes are recognized based on the age of onset. The early subtype, linked to mutations in the *COL8A2* gene, is rare and considered to have a more severe disease course.^7,8^ The more common late subtype has an onset during the second or third decade becoming symptomatic in the fifth to sixth decade^6^ with well-established links to mutations in *TCF4* and *SLC4A11*.^9^

The etiology of FECD involves a complex interaction between incompletely penetrant genetic factors that increase risk and biological and environmental factors that contribute to disease manifestation.^4^ The profile of genetic risk related to FECD has become clearer thanks to recent large-scale genetic analyses.^10^ Biological factors such as estrogen levels and oxidative stress, and environmental factors such as UV light exposure, have been implicated in FECD.^4,11^ This increased understanding of FECD pathogenesis prompted the present study, wherein we reasoned that additional genetic, non-genetic biological and environmental factors that relate to FECD might also be involved in other disorders, and thus lead to an increased incidence in patients with FECD. This possibility has not been a major research focus but is supported from two reports. Xu and colleagues^12^ analyzed a U.S. Medicare database to demonstrate that patients with FECD were at increased risk for breast, ovarian, thyroid and several cutaneous malignancies, while also being at decreased risk for lung and prostate cancer. Chang and colleagues^13^ analyzed the Taiwan Longitudinal Health Insurance Database 2000 and noted that patients with ocular allergic conditions were more likely to be diagnosed with FECD. These reports motivate our effort to comprehensively analyze the relationship between FECD and other disease phenotypes. To achieve this goal, we used the US Department of Veterans Affairs (VA) sponsored Million Veteran Program (MVP), an ongoing observational cohort and mega-biobank, to explore whether other disease phenotypes are associated with FECD. MVP integrates data from EHRs, health surveys, and genomic data,^14,15^ with more than 825,000 Veterans successfully enrolled across the VA healthcare system (www.mvp.va.gov). MVP has substantial ethnic diversity that parallels the US population providing research opportunities across multiple ethnic groups.^15^

Before embarking on this analysis, we first addressed an important challenge in using EHR-linked biobanks to accurately identify cases and controls through computable or electronic phenotyping. Computable phenotyping utilizes diagnostic, procedure, and office visit codes as well as prescription and/or laboratory tests to identify cases and controls for the disease of interest.^16^ Rules-based algorithms have been successfully used within MVP to identify other ocular diseases including age-related macular degeneration^17^ and primary open angle glaucoma.^18^ To support our analysis, we developed a FECD phenotype algorithm based on International Classification of Diseases Clinical Modification (ICD-CM) codes and Current Procedural Terminology (CPT) codes to accurately identify FECD cases and controls within the VA Computerized Patient Record System (CPRS).^19^

We used our FECD case-control algorithm to conduct a phenotype wide association study (PheWAS) for FECD. Although PheWAS was originally developed to understand genetic pleiotropy,^20^ the use of PheWAS has been broadened to identify relationships between a disease of interest and other conditions.^21,22^ Herein we used the PheWAS approach to identify relationships between FECD and other disease phenotypes. Considering prior reports that FECD incidence may be higher in females^4,23,24^ and may vary between racial groups^25,26^ we incorporated these factors into our analysis and noted that female sex and European ancestry were associated with FECD. Our main finding was that diagnostic codes for multiple ocular and non-ocular conditions were elevated in FECD cases. Further, these associations and laboratory value alterations identified in FECD indicate that diabetes mellitus (DM) may contribute to the development of FECD.

## Materials & Methods

### Study Participants

The MVP provides access to health, genetic and lifestyle information for U.S. Veterans who provided informed consent (https://www.research.va.gov/mvp/). When this study was conducted, MVP had provided data for 658,582 Veterans. Local chart reviews were conducted at the Eye Clinics of three VA Medical Centers (VAMCs; VA Northeast Ohio Healthcare System (Cleveland, OH), Providence VA Medical Center (Providence, RI), and VA Western New York Healthcare System (Buffalo, NY). This study adhered to the tenets of the Declaration of Helsinki and was approved by the VA Office of Research & Development Central Institutional Review Board (VA CIRB 18-41).

### Developing a Case-Control Algorithm for FECD

We designed a rules-based algorithm utilizing structured EHR data (ICD-9-CM, ICD-10-CM, and CPT codes) to identify FECD cases and controls within CPRS at each VAMC. Cases greater than 18 years old were identified based on the presence of two FECD ICD-9-CM or ICD-10-CM codes along with the absence of other ICD-9-CM or ICD-10-CM codes for corneal conditions or complicated cataract surgery that could confound the diagnosis. Controls were identified by excluding ICD-9-CM or ICD-10-CM codes for FECD, confounding corneal conditions, and complicated cataract surgery in Veterans 65 years and older. The algorithm was refined over five iterations; in each iteration, we used the algorithm to identify up to 25 cases and 25 controls, in both non-Hispanic white (NHW) and non-Hispanic black (NHB) Veterans and evaluated algorithm performance in those patients with chart reviews. Race and ethnicity were based on the consensus self-reported identify maintained by the VA Information Resource Center.

Chart review by Cleveland VAMC optometrists allowed for our FECD case-control algorithm to be tested and refined until the final version was determined. To be confirmed as a case, Veterans must have documented slit lamp findings of bilateral corneal endothelial pathology. These structural signs were accompanied by FECD-relevant descriptors: bilateral corneal guttae, a “beaten metal appearance”, stromal corneal edema, bullae, and bullous keratopathy.^1,6^ Case exclusion criteria included prior complicated ocular surgeries or traumas causing secondary corneal endothelial disease. Cases of postoperative intraocular surgery with pseudophakic corneal edema, pseudophakic bullous keratopathy, or corneal decompensation were included if FECD was documented pre-operatively. Clinicians were instructed to use best clinical judgment and available clinical documentation to determine inclusion status.

To be confirmed as controls, Veterans must have no signs of FECD or any other corneal endothelial disease. Finalized inclusion criteria codes included H18.51 for ICD-10-CM and 371.57 for ICD-9-CM. Exclusion criteria codes are displayed in **Supplementary Table 1**.

Figure 1. presents an overview of the process used to evaluate algorithm performance. We first calculated positive and negative predictive values (PPV, NPV) for the Cleveland VAMC site, in parallel for NHB and NHW Veterans. Once we achieved >85% PPV and NPV at the Cleveland VAMC, the algorithm was also evaluated by Eye Clinic staff at the Buffalo and Providence VAMCs.

**Figure 1:**
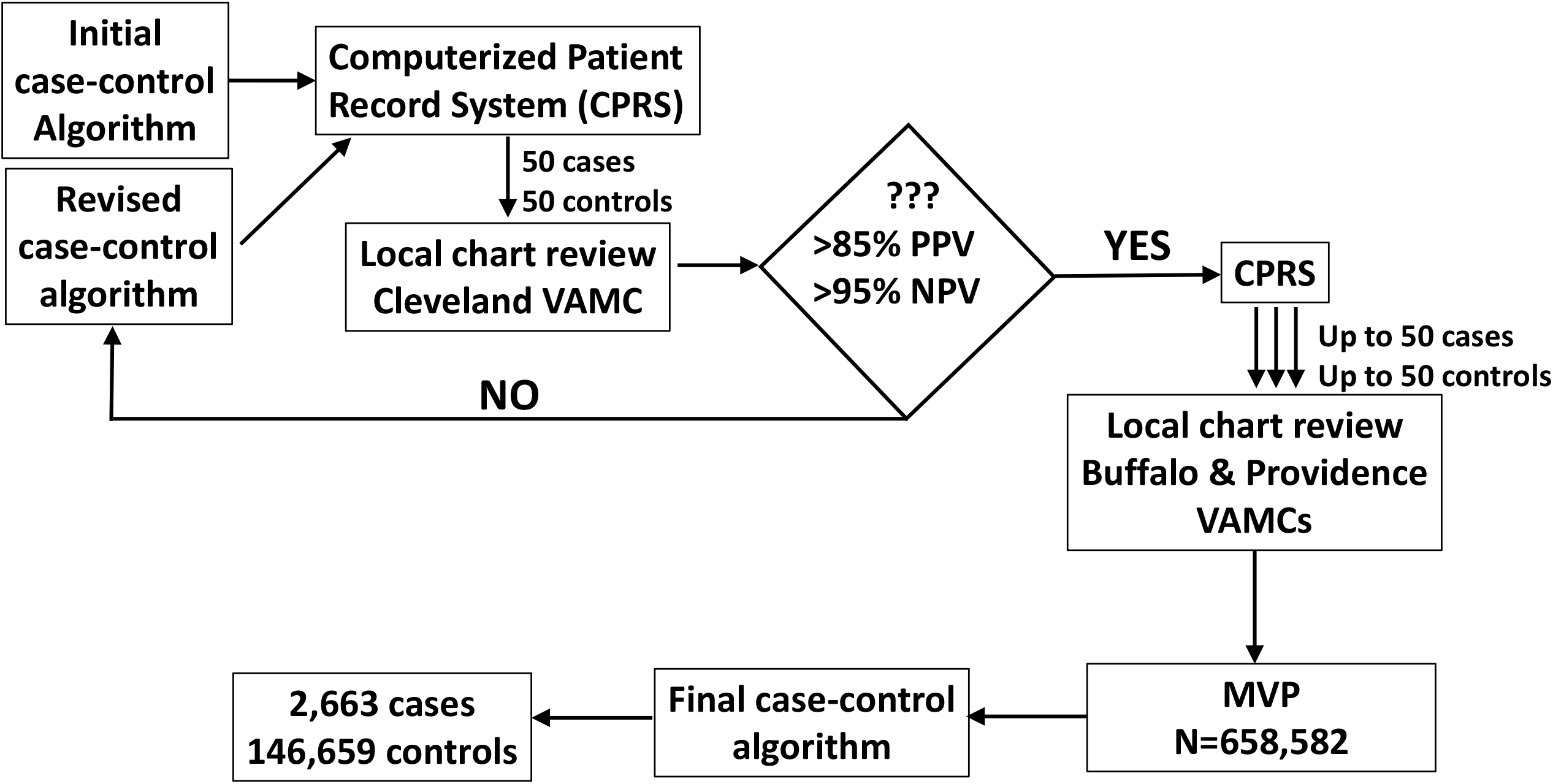
Overview of the algorithm refinement process and application to MVP. FECD case-control algorithms were evaluated and revised at the Cleveland VAMC Eye Clinic. Upon reaching >85% PPV and >95% NPV, algorithm performance was confirmed at Buffalo and Providence VAMC Eye Clinics, and then applied to the MVP.

### Analyses Within the Million Veteran Program Biobank

We applied the final FECD case-control algorithm to the available MVP cohort of 658,582 Veterans with available genetic data. Following identification of cases and controls, data on sex and current age were extracted from medical records or the MVP baseline lifestyle survey. Using participant genotypes^27^ we classified individuals by genetic ancestry into admixed African (AFR), Asian (ASN), European (EUR), and admixed American (HIS) populations via the HARE (harmonized ancestry and race/ethnicity) machine learning algorithm.^28^

We used single and multiple variable logistic regression to understand the impact of sex, genetic ancestry (coded as a categorical variable) and Charlson Comorbidity Index (CCI) on FECD. Since our algorithm used age to select cases and controls, we could not test the effect of age, but did use this variable to adjust for confounding. In our multiple regression model, we also examined comorbidity in cases and controls using the CCI.^29^ Based on prior reports that US adults have an average of 2.2 comorbid conditions,^30,31^ we compared Veterans without comorbid conditions to those with either 1-3 or 4 or more comorbidities, coded as categorical variables.

### Association of FECD Diagnosis with other Phenotypes and Laboratory Values

Phecodes combine ICD codes into clinically meaningful phenotypes^32,33^ and have traditionally been used to perform PheWAS in the context of genetic risk factors.^20^ In our study we used phecodes to understand phenotypic relationships between FECD and multiple conditions. We also determined if relationships existed between FECD and laboratory values. Phenotype and laboratory analyses were both implemented in the R PheWAS package.^34^ For each ICD code, cases were defined as at least 2 counts of the code on separate days, and controls were 0 counts. Subjects with 1 count were excluded. Additionally, we applied phenotype-based control exclusion (i.e., exclusion of controls that were cases in similar phenotypes) and we excluded females in male-specific phenotypes and vice versa. We only considered phenotypes with 100 or more cases. For laboratory values, we used the median value in regression analyses, and required that everyone had at least two lab measurements to be included. Labs with measurements in at least 50 individuals were considered. All ICD code-based phenotypes were tested with logistic regression, and all lab-based phenotypes were tested with linear regression, adjusting for age, age-squared, sex (for non-sex specific phenotypes), and 20 Principal Components of genome-wide genetic markers representing population structure in both.

## Results

### Developing an Accurate FECD Case-Control Algorithm

Initial algorithm development and refinement via chart review was completed at the Cleveland VAMC Eye Clinic. At each iteration, the algorithm identified 50 putative cases (25 NHW, 25 NHB) and 50 putative controls (25 NHW, 25 NHB) whose charts were then individually reviewed. The first four iterations identified errors which were used to refine the algorithm. For example, during early algorithm development, multiple putative cases were identified as epithelial basement membrane dystrophy which affects the corneal epithelium rather than the endothelium which is involved in FECD. Control accuracy was also affected by coding errors. For example, some putative controls had clear changes to the corneal endothelium (guttae) but this was not coded in the medical record.

For the fifth and final iteration, review of the cases identified six errors in which postoperative complications made it difficult to determine if FECD was present before surgical intervention, yielding a PPV of 88%. Review of the 50 controls identified three errors where guttae were present, but a FECD ICD-9-CM or ICD-10-CM was not recorded in the chart, yielding a NPV of 94%.

We next evaluated algorithm performance at the Buffalo VAMC and Providence VAMC Eye Clinics. At Buffalo, 42 FECD cases (25 NHW, 17 NHB) and 50 controls (25 NHW, 25 NHB) were identified by the algorithm. After chart review by eye care providers, a PPV of 78% and NPV of 98% were determined. At Providence, 26 FECD cases (25 NHW, 1 NHB) and 49 (24 NHW, 25 NHB) controls were reviewed by eye care providers. The PPV was 77% while the NPV was 100%.

Combined across all three VA sites, the FECD algorithm performed with an 82% PPV and 97% NPV (**Table 1**). The PPV was higher in NHB Veterans (93%) than in NHW Veterans (76%). NPV was similar between NHB and NHW Veterans at 98% and 96%, respectively.

**Table 1:** Summary of Cases and Controls reviewed in final FECD algorithm.

|  |  | Algorithm Identified |  |  |  |
| --- | --- | --- | --- | --- | --- |
|  |  | **FECD cases**<br>**(female)** | **Controls**<br>**(female)** | **PPV** | **NPV** |
| **Non-Hispanic**<br>**Black** | Cleveland | 25 (4) | 25 (0) | 96% | 96% |
|  | Providence | 1 (0) | 25 (0) | 100% | 100% |
|  | Buffalo | 17 (3) | 25 (1) | 88% | 100% |
|  | Combined | 43 (7) | 75 (1) | 93% | 98% |
| **Non-Hispanic**<br>**White** | Cleveland | 25 (2) | 25 (0) | 80% | 92% |
|  | Providence | 25 (0) | 24 (0) | 76% | 100% |
|  | Buffalo | 25 (2) | 25 (0) | 72% | 96% |
|  | Combined | 75 (4) | 74 (0) | 76% | 96% |
| **Total** |  | 118 (11) | 149 (1) | 82% | 97% |

FECD: Fuchs endothelial corneal dystrophy
PPV: positive predictive value
NPV: negative predictive value

### FECD Risk Factors include Female Sex and European Ancestry

When applied to the genotyped MVP cohort, the algorithm identified 2,663 cases and 146,659 controls (**Table 2**), exceeding the sizes of the case and control cohorts analyzed in the largest reported genome-wide association study for FECD.^10^ While this seems like a large drop-off from the original MVP cohort of 658,582, a major factor is the requirement that controls be 65 years of age or older and presence of an eye exam. Most cases and controls were male, reflecting the historical prevalence of males in the U.S. military and that >90% of MVP enrollees are male.^35^ Stratified by ancestry, our algorithm identified 2,264 EUR, 315 AFR, 80 HIS, and 4 ASN cases, partially reflecting the relative ancestry proportions in MVP (∼70% EUR overall).

**Table 2:** Genotyped MVP participants who meet the criteria for case with FECD or control.

| **Genetic ancestry** | **Cases**<br>**all / male / female** | **Controls**<br>**all / male / female** | **% FECD**<br>**all / male / female** |
| --- | --- | --- | --- |
| EUR | 2,264 / 2,057 / 207 | 114,408 / 111,935 / 2,473 | 1.94 / 1.80 / 7.72 |
| AFR | 315 / 225 / 90 | 16,367 / 15,907 / 460 | 1.89 / 1.39 / 16.36 |
| HIS | 80 / 75 / 5 | 7,105 / 6,962 / 143 | 1.11 / 1.07 / 3.38 |
| ASN | 4 / 4 / 0 | 1,064 / 1,035 / 29 | 0.37 / 0.38 / not done |
| Total | 2,663 / 2,361 / 302 | 138,944 / 135,839 / 3,105 | 1.88 / 1.71 / 8.86 |

MVP: Million Veteran Program
FECD: Fuchs endothelial corneal dystrophy
EUR: European ancestry
AFR: African ancestry
ASN: Asian ancestry
HIS: admixed American ancestry

Prior reports indicate the frequency of FECD is elevated in females as compared to males, although the estimate of the magnitude of this difference varies^4,23,24^ and has not been examined in diverse populations. We examined the frequency of females *vs*. males among cases and noted that the frequency of FECD varied across race and was much higher in females (**Table 2**). Single variable logistic regression (**Supplementary Table 2**) showed that FECD cases had a 5.6X greater odds of being female (OR=5.60; CI: 4.93-6.33; p = 1.07 × 10^−160^). Single variable analysis of genetic ancestry as a categorical variable (EUR *vs* AFR, EUR *vs* HIS, and EUR *vs* ASN) showed that individuals of EUR ancestry were at greatest risk for FECD (**Supplementary Table 2**). The mean (± S.D.) age of FECD cases was significantly younger (69.52 ± 10.04 years) than controls (71.68 ± 6.05 years) (**Fig. 2**). As our algorithm implemented different age criteria for FECD cases and controls, we used logistic regression to adjust for this age difference in subsequent multivariable logistic regression models (**Table 3**) but did not formally test for age differences. We incorporated the CCI in the model to determine whether the non-ocular conditions as ascertained by higher scores demonstrated association with FECD.

**Table 3:** Multivariable logistic regression examining various factors influencing risk to FECD (N = 141,607).

| Variable | OR | 95% CI | P value |
| --- | --- | --- | --- |
| Age | 0.92 | 0.92, 0.93 | < 0.001 |
| Sex | 4.60 | 4.03, 5.22 | < 0.001 |
| Ancestry – AFR vs EUR | 0.81 | 0.71, 0.91 | < 0.001 |
| Ancestry – HIS vs EUR | 0.53 | 0.42, 0.66 | < 0.001 |
| Ancestry – ASN vs EUR | 0.18 | 0.06, 0.42 | < 0.001 |
| CCI Cat 1-3 | 1.21 | 1.06, 1.40 | 0.005 |
| CCI Cat 4+ | 1.40 | 1.11, 1.75 | 0.003 |

FECD: Fuchs endothelial corneal dystrophy
AFR: admixed African ancestry
EUR: European ancestry
HIS: admixed American ancestry (Hispanic/Latino)
ASN: Asian ancestry
CCI: Charlson Comorbidity Index
OR: odds ratio
CI: confidence interval

**Figure 2:**
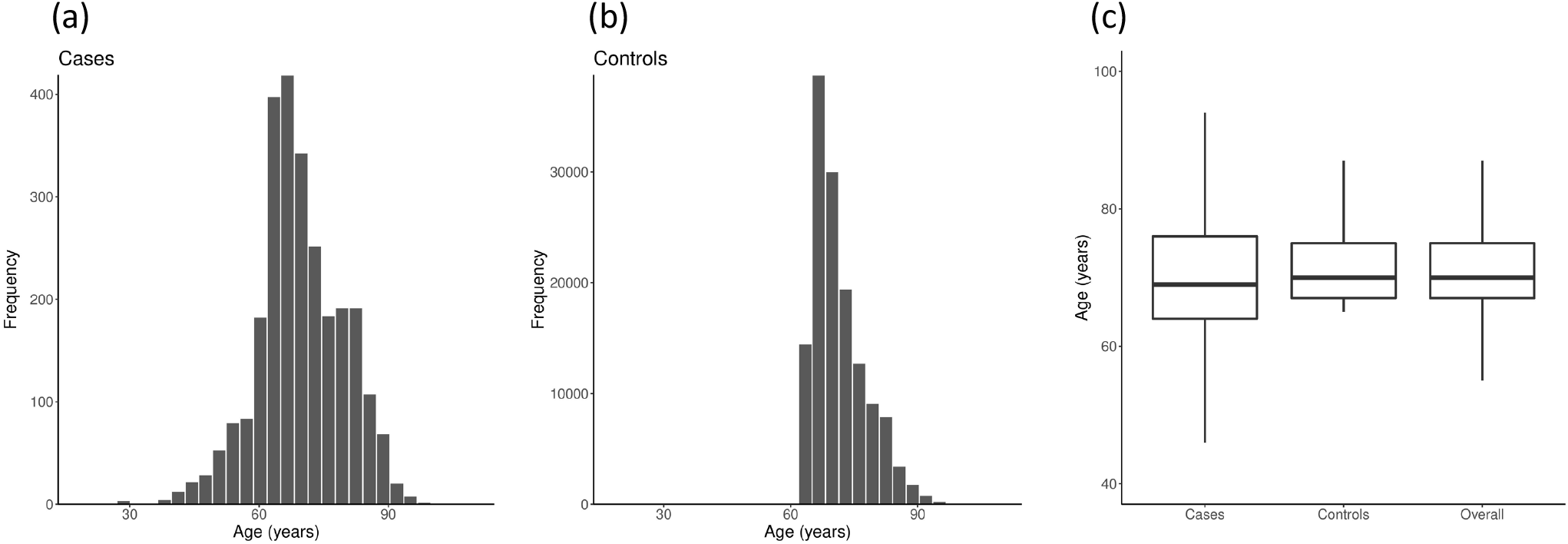
Age distribution of FECD cases (N=2,663) (a) and controls (N=138,944) (b). (c) Box-plot representation.

Although women comprise only ∼9% of MVP enrollees, we found a much higher risk for females after adjusting for age, and ancestry group (OR=4.60; CI: 4.03 – 5.22; p < 0.001). In fact, female sex was the most significant effect in our analyses. Non-European ancestry (AFR, ASN, HIS) was associated with reduced risk (OR=0.81, 95% CI: 0.71 - 0.91, p < 0.001; OR=0.18, 95% CI: 0.06 - 0.42, p < 0.001; OR=0.53, 95% CI: 0.42 – 0.66, p < 0.001, respectively).

### FECD is associated with multiple other disease conditions

The CCI predicts long-term mortality for multiple medical conditions and can be used to identify the impact of overall health on diagnostic and prognostic factors.^36^ After adjusting for other covariates (age, sex and ancestry group), we found that cases with FECD were 1.21 times more likely to have 1-3 comorbid conditions identified via the CCI, compared with a control without FECD (95% CI: 1.06 - 1.39, p = 0.005). A similar analysis of an increasing number of comorbid conditions, 4 or greater, elevated the odds ratio to 1.40 (95% CI: 1.11 - 1.75; p = 0.003).

We used a PheWAS to better understand the nature of the elevated number of comorbidities noted in our more numerous Veterans of European ancestry. Of the total number of 1,814 diagnostic phecodes, 1,417 met our criteria of including at least 100 cases. Of these, 213 (15.0%) were more prevalent in FECD cases than controls after Bonferroni correction (**Supplementary Table 3**). Significant associations were noted in virtually every phecode group (**Figure 3; Table 4**). The phegroup **‘Sense Organs**’ had the highest number and rate of significant phecodes (51 out of 118; 43.22%); the five most significant phecodes were for corneal dystrophy (***cornea replaced by transplant, corneal dystrophy, corneal opacity and other disorders, Fuchs dystrophy, corneal edema)***. Other highly significant phecodes were ***inflammation of the eye*** (OR=2.25; 95% CI: 2.03 – 2.49; p <2.59 X 10^−51^), ***dry eyes*** (OR=1.95; 95% CI: 1.78 – 2.15; p <1.24 X 10^−44^), and ***uveitis, noninfectious or NOS*** (OR=4.43; 95% CI: 3.60 – 5.39; p <8.86 X 10^−34^). The phegroup **‘Dermatologic’** also had a high rate of significant phecodes (19 out of 73; 26.0%) and these included ***corns and callosities*** (OR=1.56; 95% CI: 1.36 – 1.77; p <1.99 X 10^−10^), ***other hypertrophic and atrophic conditions*** (OR=1.53; 95% CI: 1.36 – 1.72; p <5.78 X 10^−12^) and ***disturbances of skin sensation*** (OR=1.54; 95% CI: 1.34 – 1.76; p <2.59 X 10^−9^). The ‘**Musculoskeletal’** phegroup had 16 out of 109 (14.7%) phecodes that were significantly higher in cases than controls, and these included a number related to arthritis (***peripheral enthesopathies and allied syndromes*** (OR=1.45; 95 CI: 1.32 – 1.59; p<8.86 X 10^−14^), ***osteoarthritis NOS*** (OR=1.35; 95% CI: 1.23 – 1.48; p<1.58 X 10^−10^), and abnormal wear of spinal disks (***spondylosis and allied disorders*** (OR=1.42; 95% CI: 1.27 – 1.58; p<1.68 X 10^−9^), ***spondylosis without myelopathy*** (OR=1.40; 95% CI: 1.25 – 1.57; p<1.58 X 10^−8^).

**Figure 3:**
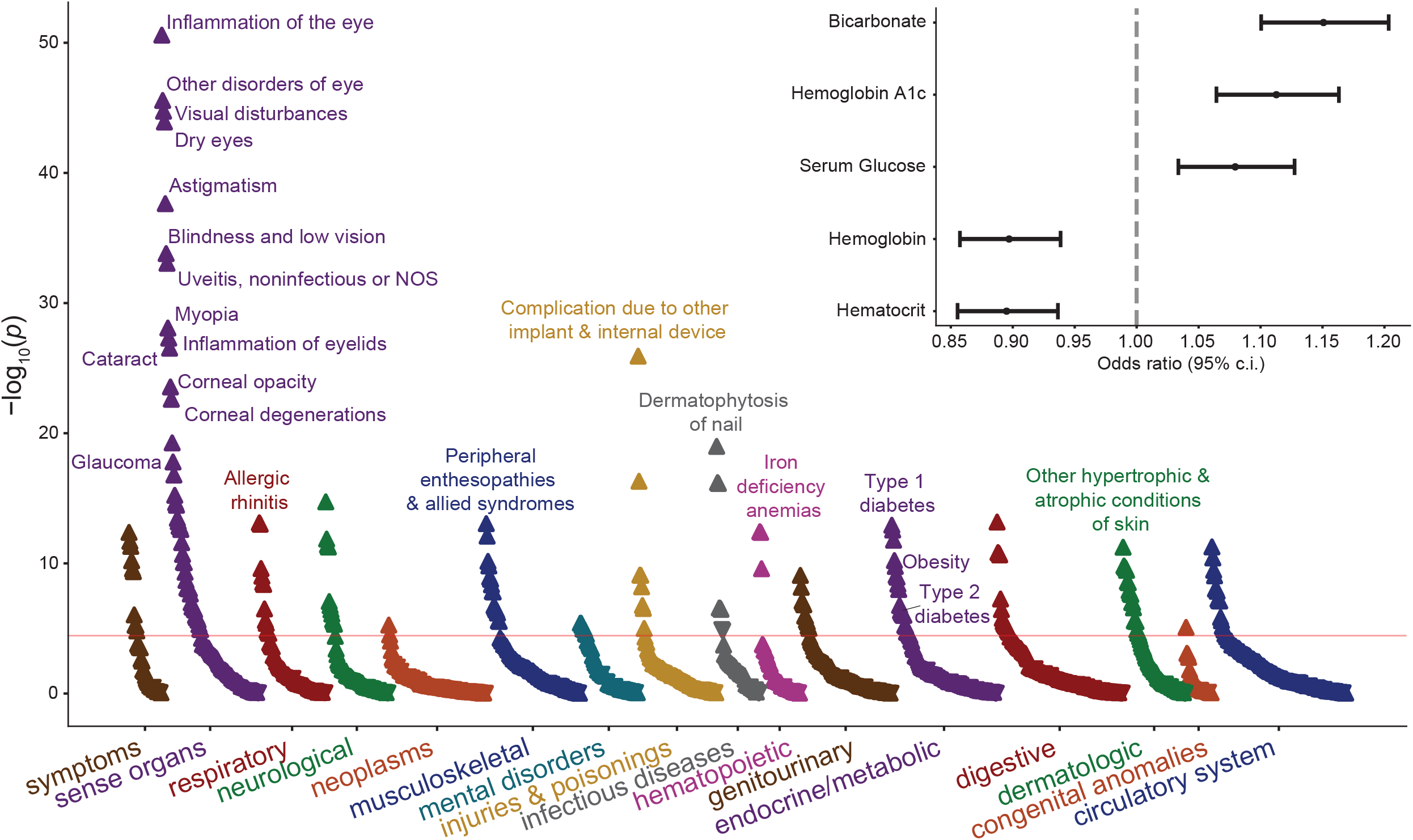
Phenome-wide association scan of EHR-derived phenotypes (main panel) and normalized laboratory values (inset) with FECD. Upwards-pointing arrows indicate positive association (OR > 1) and downwards-pointing arrows indicate negative association (OR < 1). Red line indicates Bonferroni-corrected significance threshold. Phenotypes highly similar to FECD are not plotted. Labs shown are those significant after multiple hypothesis testing correction.

**Table 4:** Summary of PheWAS organized by phecode groups. For each phecode group, the total number of phecodes that included at least 100 cases and those that achieved statistical significance after Bonferroni correction are noted.

| **Phecode group** | **Total phecodes** | **Significant phecodes** | **% significant** |
| --- | --- | --- | --- |
| circulatory system | 17 | 156 | 10.90 |
| congenital anomalies | 1 | 30 | 3.33 |
| dermatologic | 19 | 73 | 26.03 |
| digestive | 15 | 146 | 10.27 |
| endocrine/metabolic | 21 | 122 | 17.21 |
| genitourinary | 11 | 106 | 10.38 |
| hematopoietic | 3 | 47 | 6.38 |
| infectious diseases | 8 | 50 | 16.00 |
| injuries & poisonings | 8 | 91 | 8.79 |
| mental disorders | 8 | 67 | 11.94 |
| musculoskeletal | 16 | 109 | 14.68 |
| neoplasms | 2 | 113 | 1.77 |
| neurological | 11 | 73 | 15.07 |
| pregnancy complications | 0 | 1 | 0.00 |
| respiratory | 12 | 77 | 15.58 |
| sense organs | 51 | 118 | 43.22 |
| symptoms | 10 | 38 | 26.32 |
| **TOTAL** | **213** | **305** | **15.03** |

Within the ‘**Endocrine/Metabolic**’ phegroup, we noted a number of phecodes related to DM. For example, the odds of having the ***insulin pump user*** phecode was 2.2 times higher among FECD cases versus controls (95% CI: 1.82 – 2.68; p<2.75 X 10^−13^). Phecodes for ***Type 1 diabetes*** (OR=2.01; 95% CI: 1.69 – 2.37; p<1.18 X 10^−13^) and ***Type 2 diabetes*** (OR=1.25; 95% CI: 1.15 – 1.37; p<4.55 X 10^−7^) were both more prevalent in FECD cases, along with a number of phecodes related to associations of Type 1 or Type 2 DM and specific systems (neurological, ophthalmic, renal). The phecode ***obesity*** was also more prevalent in FECD cases (OR=1.34; 95% CI: 1.22 – 1.47; p<1.73 X 10^−10^) and this may be related to the multiple ‘**Digestive**’ (15 out of 146; 10.3%) and ‘**Respiratory**’ (12 out of 77; 15.6%) phecodes that that were significantly more prevalent in FECD cases than controls.

The only phegroups where FECD cases were not distinguished from controls were ‘**pregnancy’** (0 out of 1; 0%), ‘**congenital anomalies**’ (1 out of 30; 3.3%) and ‘**neoplasms**’ (3 out of 113; 2.7%). Disorders of pregnancy and congenital anomalies have relative low relevance to the adult Veteran population that we examined.

In our PheWAS for Veterans of AFR ancestry, forty out of 942 (4.25%) phecodes were more prevalent in FECD cases than controls. While this analysis had less power due to the smaller sample size, many of the phecodes that were significant reflected our EUR analysis, including ***Fuchs*’ *dystrophy, corneal dystrophy, corneal opacity and other disorders of cornea, corneal edema, dry eyes***, and ***osteoarthrosis***. Both Type 1 and Type 2 DM were more frequent in AFR cases, attaining nominal significance that did not withstand multiple testing correction, but displaying the same direction of effect. In sum, the effects were consistent across groups, but lacked adequate sample sizes in AFR.

While more work is needed to fully understand this higher rate of non-ocular phecodes in FECD cases than controls, our analysis of laboratory data were consistent with an important role for DM. Out of the 69 available measures, five were significantly altered in FECD cases of EUR ancestry after Bonferroni correction (**Supplementary Table 4**). For four of these, the change direction was consistent with DM. FECD cases had elevated levels of hemoglobin A1C (OR=1.11; CI: 1.06 – 1.16; p<2.47 X 10^−6^) and serum glucose (OR=1.08; CI: 1.03 – 1.13; p=5.54 X 10^−4^) while hematocrit (OR=0.90; CI: 0.86 – 0.94; p<1.57 X 10^−6^) and hemoglobin (OR=0.90; CI: 0.86 – 0.94; p<2.60 X 10^−6^) were decreased. Changes in bicarbonate levels had the strongest association, showing elevation in FECD cases (OR=1.15; CI: 1.10 – 1.20; p<7.31 X 10^−10^), but the cause of this pH increase in blood and relationship to FECD is unknown. Hemoglocin A1C and serum glucose were also elevated in FECD cases of AFR ancestry, but these changes did not reach statistical significance due to the smaller sample size. Effect estimates were consistent with results from the EUR ancestry analysis.

In summary, the most important demographic risk factors for FECD are female sex and European ancestry. Further, we note that FECD risk increases with increasing multi-morbidity and multiple systemic conditions. The associations with phecodes related to endocrine/metabolic conditions including DM are supported by the significant laboratory values that are associated with FECD.

## Discussion

PheWAS has traditionally been used to identify pleiotropy, where a genetic trait is associated with multiple conditions.^20^ In this study, we applied the PheWAS approach to the MVP biobank to comprehensively investigate whether FECD may be associated with other conditions. We noted that Veterans with a CCI score of 0 (zero comorbidities) were less likely to have a diagnosis of FECD as compared to those with at least one comorbidity. To explore this observation, we examined the association between FECD status and the entire library of phecodes. In comparison to controls, we noted that FECD cases had higher rates of many conditions, including endocrine/metabolic, infectious disease, neurological, and musculoskeletal phegroups and indicating that as a group FECD cases have many more healthcare challenges than controls. Among these, a primary association converged on DM, where we noted that FECD cases had elevated rates of DM-related phecodes and laboratory profiles consistent with DM.

What underlies the many associations noted between FECD and other conditions? We favor a model in which these other conditions render the cornea more vulnerable to developing the FECD phenotype. FECD genetic risk factors are incompletely penetrant, so an additional factor is thought to be needed to induce the disease phenotype. A number of such factors that contribute to the FECD phenotype have been identified, including estrogen, oxidative stress, and exposure to UV light.^4,11^ Our results indicate that DM may be a particularly important factor that contributes to FECD etiology. This concept is consistent with a broad literature. For example, DM is known to alter corneal physiology^37-40^ leading to complications that increase keratopathy. These include abnormal tear secretion perhaps secondary to reduced goblet cell density, decreased corneal sensitivity, delayed corneal reepithelialization, and altered corneal morphology, function and biomechanics.^37,41-46^ Consistent with these observations, the Cornea Preservation Time Study documented elevated risk for graft failure for eyes that received a cornea transplant from donors with DM.^47^ The mechanism underling the association between DM and FECD may involve TCF4. Expansion of a common trinucleotide repeat within the *TCF4* locus is strongly linked to FECD,^48^ and *TCF4* had the strongest signal in a large-scale genome wide analysis of FECD.^10^ TCF4 (also referred to as E2-2) is involved in the regulation of insulin genes^49^ and is required to maintain the fate of mature plasmacytoid dendritic cells,^50^ a cell type which has been implicated in Type 1 DM.^51^

We also noted that the link between FECD and DM was stronger for the earlier onset Type 1 than the Type 2 form, consistent with the relationship between DM and microvascular complications of the retina, kidney, peripheral nervous system and other systems. It will be interesting to determine whether the presence of DM results in a more severe FECD phenotype in patients with comparable risk factors, and whether DM also impacts other corneal dystrophies. It will also be interesting to determine, for example, if FECD is reduced in prevalence or severity in patients with well-managed DM and whether the beneficial effects of topical insulin observed in animal models^38^ will alleviate FECD severity in patients with DM.

Diabetes mellitus may also be central to many of the associations between FECD and other phecodes. For example, anemia can be induced by the shorter life span and greater fragility of red blood cells in DM^52-56^ In addition, DM has been linked to multiple dermatologic conditions, as well as enthesopathy, osteoarthritis and spinal disk disorders.^57-59^. In comparison, while DM increases the risk of gout, this may reflect other comorbid conditions and not a direct effect of DM.^60^

We found that FECD had higher rates of other ocular conditions including cataracts and dry eye. Both are more common in DM,^37^ suggestive of a central role for DM in the etiology of FECD. However, we cannot rule out the possibility that the elevated rate of diagnostic codes for these and potentially other ocular conditions may simply reflect these patients undergoing more intensive ophthalmic care. Further, the association with cataract may reflect that cataract surgery alone can induce a phenotype involving damage to the corneal endothelium and resembling FECD.^61-65^ The association with dry eye may also reflect that FECD symptoms related to corneal pain and/or sensitivity inspire clinicians to add codes related to dry eye to the medical record. Prior studies have reported that patients with FECD are more likely to have glaucoma.^66^ We cannot exclude the possibility that pleiotropy underlies some of the FECD associations that we observed. A more complete understanding of the genetic factors that contribute to FECD and conditions associated with FECD will be needed to understand the relative roles played by pleiotropy versus the physiological impacts of diabetes and potentially other disorders on the cornea in FECD etiology.

FECD has been reported to be more common in women than in men.^4,24,25^ While >90% of U.S. Veterans are male, 1.80% of EUR males (=2,057/111,935) and 7.72% (=207/2,473) of EUR females were classified as cases, consistent with excess risk to females. Using our case-control algorithm, we observed a 1.78% prevalence of FECD in the MVP (2,663/149,322). This is somewhat lower than prior reports that FECD prevalence is approximately 4%.^5,64^ A lower estimate of FECD prevalence could reflect that MVP enrollees are >90% male^35^ and that males have a lower occurrence of FECD. A lower estimate of FECD prevalence could also reflect that our FECD algorithm (a) eliminated individuals with a single positive code and (b) included many exclusion codes that could potentially co-exist with FECD. Our estimate of FECD prevalence is, however, much higher than an analysis of data from a national managed-care network which found that the prevalence of all corneal dystrophies was less than 0.1%.^68^ This study did not have an age requirement and therefore included data from young patients not yet at risk for FECD. Additional studies of FECD prevalence in large healthcare databases should help to define a precise estimate of FECD prevalence.

We further found that FECD frequency was higher in EUR Veterans than in other ancestries, after adjustment for age and sex. This finding agrees with that of Eghrari et al.^69^ who reported a modest reduction in the incidence of FECD in NHB as compared to NHW patients. In comparison, Mahr and colleagues^25^ did not observe a difference in FECD incidence between NHW and NHB patients based on their analysis of U.S Medicare claims. Any comparative increase in FECD incidence in NHW may reflect previously reported average differences in corneal thickness across populations.^70^

## Limitations and Strengths

As MVP does not provide imaging data, this study was limited to clinical coding, billing, and prescription data. We addressed this limitation by developing an algorithm to accurately identify FECD cases and controls within CPRS. Through a local chart review, involving three VAMC Eye Clinics, we were able to define a final algorithm that had high PPV (probability of a case identified by the algorithm having FECD) and high NPV (probability of a control identified by the algorithm not having FECD). Our algorithm thus provides the opportunity to pursue FECD genetic studies within MVP and additional largescale biobanks.

In contrast to our algorithm for FECD, we did not use a similar approach to validate the many phecodes that were examined in the PheWAS. These phecodes could suffer from coding and diagnostic errors, although we would expect these to be random such errors will reduce power by introducing noise.

This study was limited to the population served by the Veterans Health Administration. While it is unclear that the Veteran and general population are markedly different, it will be valuable to examine these associations in additional patient cohorts, including those including a larger fraction of women enrollees.

The statistical model used to conduct our PheWAS and our analysis of laboratory values adjusted for obvious confounders (age, sex, genetic ancestry). We cannot rule out however, that some hidden residual confounding may remain.

## Conclusions

By developing a rules-based algorithm to identify FECD and applying this to a large EHR-based biobank, we provide new insights into the etiology of FECD. In the future it will be important to better understand the relationship between FECD and DM. Insights regarding this relationship may identify opportunities for slowing FECD progression. We anticipate that our case-control algorithm will open the door for further FECD gene discovery.

## Data Availability

All data produced in the present work are contained in the manuscript

## Acknowledgments

We are grateful to the VINCI and GENISIS support teams, and to the MVP Core Statistical Analysis team. This publication is solely the responsibility of the authors and does not necessarily represent the official views of the NIH. This publication does not represent the views of the Department of Veterans Affairs or the United States Government.

