## Supplemental Tables 1-4 for "Association between Fuchs Endothelial Corneal Dystrophy, Diabetes Mellitus and Multimorbidity"

**Supplementary Table 1:** Exclusion codes used in final FECD case-control algorithm.

| Condition | ICD-10-CM | ICD-9-CM |
|---|---|---|
| Epithelial corneal dystrophy | H18.52 | |
| Cogan's corneal dystrophy | | 371.52 |
| Crystalline corneal dystrophy | | 371.56 |
| Granular corneal dystrophy | H18.53 | 371.53 |
| Lattice corneal dystrophy | H18.54 | 371.54 |
| Macular corneal dystrophy | H18.55 | 371.54 |
| Marginal corneal degeneration (Terrien's)/Peripheral corneal degeneration | H18.46X | 371.52 |
| Meesman's/epithelial | | 371.51 |
| Nodular, Salzmann's | | 371.46 |
| Ring-like corneal degeneration | | 371.52 |
| Corneal edema secondary to contactlens | H18.21X | |
| Keratoconus | H18.6XXX | 371.60 |
| Other and unspecified corneal deformities | H18.7XXX | |
| Other specified disorders of cornea | H18.8XXX | |
| Complicated cataract surgery | T85.29XA, H59.XXX | |

**Supplementary Table 2:** Single regression models examining various factors individually imparting risk for FECD.

| Variable | OR | 95% CI | P value | N |
|---|---|---|---|---|
| Sex | 5.60 | 4.94, 6.34 | $1.62 \times 10^{-160}$ | 141,607 |
| Ancestry – AFR vs EUR | 0.95 | 0.83, 1.05 | $2.75 \times 10^{-01}$ | 133,354 |
| Ancestry – HIS vs EUR | 0.57 | 0.45, 0.71 | $1.17 \times 10^{-06}$ | 123,857 |
| Ancestry – ASN vs EUR | 0.19 | 0.06, 0.45 | $7.63 \times 10^{-04}$ | 117,740 |

AFR: admixed African ancestry (African American)
EUR: European ancestry
HIS: admixed American ancestry (Hispanic/Latino)
ASN: Asian ancestry
OR: odds ratio
CI: confidence interval

**Supplementary Table 3:** Phecode association study comparing EUR FECD cases and controls.

| PHECODE | GROUP | DESCRIPTION | OR | beta | se | CI 2.5% | CI 97.5% | p | n cases | n controls | n total |
|---|---|---|---|---|---|---|---|---|---|---|---|
| 364.9 | sense organs | Cornea replaced by transplant | 76.13 | 4.33 | 0.09 | 4.16 | 4.50 | <2.2E-308 | 618 | 121838 | 122456 |
| 364.5 | sense organs | Corneal dystrophy | 5001684.01 | 15.43 | 1.28 | 13.48 | 20.19 | <2.2E-308 | 2404 | 119784 | 122188 |
| 364 | sense organs | Corneal opacity and other disorders of cornea | 395133.23 | 12.89 | 1.41 | 10.96 | 17.72 | <2.2E-308 | 4562 | 114364 | 118926 |
| 364.51 | sense organs | Fuchs' dystrophy | 5359296.50 | 15.49 | 1.28 | 13.54 | 20.33 | <2.2E-308 | 2400 | 119804 | 122204 |
| 364.2 | sense organs | Corneal edema | 53.08 | 3.97 | 0.07 | 3.83 | 4.11 | <2.2E-308 | 1013 | 120610 | 121623 |
| 371 | sense organs | Inflammation of the eye | 2.25 | 0.81 | 0.05 | 0.71 | 0.91 | 2.593E-51 | 21674 | 81813 | 103487 |
| 379 | sense organs | Other disorders of eye | 2.05 | 0.72 | 0.05 | 0.62 | 0.82 | 2.728E-46 | 29621 | 72379 | 102000 |
| 368 | sense organs | Visual disturbances | 2.17 | 0.78 | 0.05 | 0.67 | 0.88 | 1.825E-45 | 19079 | 83549 | 102628 |
| 375.1 | sense organs | Dry eyes | 1.95 | 0.67 | 0.05 | 0.58 | 0.76 | 1.236E-44 | 36976 | 65370 | 102346 |
| 367.2 | sense organs | Astigmatism | 1.87 | 0.63 | 0.05 | 0.53 | 0.72 | 2.304E-38 | 44292 | 53174 | 97466 |
| 367.9 | sense organs | Blindness and low vision | 2.43 | 0.89 | 0.07 | 0.75 | 1.01 | 1.584E-34 | 9425 | 103713 | 113138 |
| 371.1 | sense organs | Uveitis, noninfectious or NOS | 4.43 | 1.49 | 0.10 | 1.28 | 1.68 | 8.855E-34 | 1606 | 119738 | 121344 |
| 367.1 | sense organs | Myopia | 1.81 | 0.59 | 0.05 | 0.49 | 0.69 | 8.266E-29 | 20804 | 84200 | 105004 |
| 371.3 | sense organs | Inflammation of eyelids | 1.91 | 0.65 | 0.06 | 0.54 | 0.76 | 4.957E-28 | 17282 | 87604 | 104886 |
| 366 | sense organs | Cataract | 4.31 | 1.46 | 0.16 | 1.15 | 1.80 | 2.935E-27 | 110133 | 4966 | 115099 |
| 859 | injuries & poisonings | Complication due to other implant and internal device | 5.82 | 1.76 | 0.13 | 1.49 | 2.02 | 1.200E-26 | 713 | 120166 | 120879 |
| 364.1 | sense organs | Corneal opacity | 2.86 | 1.05 | 0.09 | 0.87 | 1.23 | 2.784E-24 | 3072 | 116403 | 119475 |
| 364.4 | sense organs | Corneal degenerations | 4.80 | 1.57 | 0.13 | 1.30 | 1.82 | 2.479E-23 | 946 | 119609 | 120555 |
| 362 | sense organs | Other retinal disorders | 1.56 | 0.44 | 0.05 | 0.35 | 0.54 | 5.528E-20 | 45252 | 62705 | 107957 |
| 110.11 | infectious diseases | Dermatophytosis of nail | 1.58 | 0.46 | 0.05 | 0.36 | 0.55 | 1.026E-19 | 29684 | 81669 | 111353 |
| 365 | sense organs | Glaucoma | 1.51 | 0.41 | 0.05 | 0.32 | 0.50 | 1.624E-18 | 36521 | 78918 | 115439 |
| 379.3 | sense organs | Aphakia and other disorders of lens | 2.32 | 0.84 | 0.09 | 0.66 | 1.01 | 1.634E-17 | 4273 | 112475 | 116748 |
| 851 | injuries & poisonings | Complications of transplants and reattached limbs | 3.41 | 1.23 | 0.13 | 0.97 | 1.46 | 4.933E-17 | 1232 | 119083 | 120315 |

| | | | | | | | | | | | |
|---|---|---|---|---|---|---|---|---|---|---|---|
| 110 | infectious diseases | Dermatophytosis / Dermatomycosis | 1.50 | 0.40 | 0.05 | 0.31 | 0.50 | 6.260E-17 | 36215 | 71162 | 107377 |
| 110.1 | infectious diseases | Dermatophytosis | 1.50 | 0.40 | 0.05 | 0.31 | 0.50 | 7.273E-17 | 35705 | 71933 | 107638 |
| 362.9 | sense organs | Retinal edema | 2.65 | 0.98 | 0.11 | 0.76 | 1.18 | 5.923E-16 | 2457 | 117515 | 119972 |
| 351 | neurological | Other peripheral nerve disorders | 1.52 | 0.42 | 0.05 | 0.32 | 0.51 | 1.872E-15 | 21933 | 89829 | 111762 |
| 370 | sense organs | Keratitis | 2.24 | 0.81 | 0.09 | 0.62 | 0.98 | 2.252E-15 | 3660 | 112865 | 116525 |
| 362.26 | sense organs | Macular puckering of retina | 1.71 | 0.54 | 0.06 | 0.41 | 0.66 | 3.432E-15 | 11586 | 105009 | 116595 |
| 379.2 | sense organs | Disorders of vitreous body | 1.51 | 0.41 | 0.05 | 0.31 | 0.52 | 3.545E-14 | 22124 | 83907 | 106031 |
| 369 | sense organs | Infection of the eye | 2.04 | 0.71 | 0.09 | 0.54 | 0.88 | 6.481E-14 | 4597 | 109478 | 114075 |
| 561 | digestive | Symptoms involving digestive system | 1.54 | 0.43 | 0.06 | 0.32 | 0.54 | 6.676E-14 | 17270 | 89817 | 107087 |
| 476 | respiratory | Allergic rhinitis | 1.47 | 0.38 | 0.05 | 0.28 | 0.48 | 7.652E-14 | 25311 | 83821 | 109132 |
| 726 | musculoskeletal | Peripheral enthesopathies and allied syndromes | 1.45 | 0.37 | 0.05 | 0.28 | 0.47 | 8.859E-14 | 28542 | 76549 | 105091 |
| 465 | respiratory | Acute upper respiratory infections of multiple or unspecified sites | 1.52 | 0.42 | 0.05 | 0.31 | 0.53 | 9.879E-14 | 19204 | 81248 | 100452 |
| 250.1 | endocrine/metabolic | Type 1 diabetes | 2.01 | 0.70 | 0.09 | 0.53 | 0.86 | 1.179E-13 | 4825 | 112464 | 117289 |
| 367 | sense organs | Disorders of refraction and accommodation; blindness and low vision | 2.37 | 0.86 | 0.13 | 0.61 | 1.13 | 1.420E-13 | 106403 | 6620 | 113023 |
| 365.1 | sense organs | Open-angle glaucoma | 1.46 | 0.38 | 0.05 | 0.28 | 0.48 | 1.588E-13 | 26397 | 89490 | 115887 |
| 379.5 | sense organs | Disorders of iris and ciliary body | 2.81 | 1.03 | 0.12 | 0.78 | 1.26 | 2.516E-13 | 1711 | 118044 | 119755 |
| 250.3 | endocrine/metabolic | Insulin pump user | 2.22 | 0.80 | 0.10 | 0.60 | 0.99 | 2.753E-13 | 3291 | 116382 | 119673 |
| 285 | hematopoietic | Other anemias | 1.44 | 0.37 | 0.05 | 0.27 | 0.46 | 3.691E-13 | 31816 | 77111 | 108927 |
| 280 | hematopoietic | Iron deficiency anemias | 1.61 | 0.48 | 0.06 | 0.35 | 0.60 | 4.189E-13 | 12529 | 102772 | 115301 |
| 782.3 | symptoms | Edema | 1.47 | 0.39 | 0.05 | 0.28 | 0.49 | 4.202E-13 | 23996 | 83546 | 107542 |
| 735 | musculoskeletal | Acquired foot deformities | 1.48 | 0.39 | 0.05 | 0.29 | 0.50 | 8.071E-13 | 19870 | 91873 | 111743 |
| 350 | neurological | Abnormal movement | 1.43 | 0.36 | 0.05 | 0.26 | 0.45 | 1.356E-12 | 30999 | 76078 | 107077 |
| 250.24 | endocrine/metabolic | Type 2 diabetes with neurological manifestations | 1.47 | 0.39 | 0.05 | 0.28 | 0.49 | 1.659E-12 | 20957 | 95643 | 116600 |
| 760 | symptoms | Back pain | 1.38 | 0.33 | 0.05 | 0.23 | 0.42 | 2.053E-12 | 56805 | 51464 | 108269 |

| | | | | | | | | | | | |
|---|---|---|---|---|---|---|---|---|---|---|---|
| 362.2 | sense organs | Degeneration of macula and posterior pole of retina | 1.43 | 0.36 | 0.05 | 0.26 | 0.46 | 2.563E-12 | 33963 | 78288 | 112251 |
| 339 | neurological | Other headache syndromes | 1.56 | 0.44 | 0.06 | 0.32 | 0.56 | 3.985E-12 | 11800 | 99417 | 111217 |
| 770 | symptoms | Myalgia and myositis unspecified | 1.80 | 0.59 | 0.08 | 0.43 | 0.74 | 4.942E-12 | 5302 | 110335 | 115637 |
| 455 | circulatory system | Hemorrhoids | 1.51 | 0.41 | 0.06 | 0.30 | 0.53 | 5.414E-12 | 16406 | 85058 | 101464 |
| 350.2 | neurological | Abnormality of gait | 1.44 | 0.37 | 0.05 | 0.26 | 0.47 | 5.573E-12 | 25421 | 83130 | 108551 |
| 701 | dermatologic | Other hypertrophic and atrophic conditions of skin | 1.53 | 0.43 | 0.06 | 0.31 | 0.54 | 5.779E-12 | 14580 | 90952 | 105532 |
| 530.1 | digestive | Esophagitis, GERD and related diseases | 1.36 | 0.31 | 0.05 | 0.22 | 0.39 | 1.484E-11 | 54072 | 58136 | 112208 |
| 530.11 | digestive | GERD | 1.36 | 0.30 | 0.05 | 0.22 | 0.39 | 1.687E-11 | 51395 | 60633 | 112028 |
| 362.23 | sense organs | Cystoid macular degeneration of retina | 2.06 | 0.72 | 0.10 | 0.53 | 0.91 | 2.078E-11 | 3737 | 116171 | 119908 |
| 530 | digestive | Diseases of esophagus | 1.35 | 0.30 | 0.05 | 0.21 | 0.39 | 2.239E-11 | 55318 | 56836 | 112154 |
| 418 | circulatory system | Nonspecific chest pain | 1.37 | 0.31 | 0.05 | 0.22 | 0.41 | 3.812E-11 | 37759 | 66834 | 104593 |
| 250.6 | endocrine/metabolic | Polyneuropathy in diabetes | 1.77 | 0.57 | 0.08 | 0.41 | 0.72 | 6.106E-11 | 6267 | 112190 | 118457 |
| 785 | symptoms | Abdominal pain | 1.41 | 0.35 | 0.05 | 0.24 | 0.45 | 6.326E-11 | 22818 | 81853 | 104671 |
| 716.9 | musculoskeletal | Arthropathy NOS | 1.56 | 0.45 | 0.07 | 0.32 | 0.57 | 7.050E-11 | 11170 | 100138 | 111308 |
| 368.3 | sense organs | Anisometropia | 2.64 | 0.97 | 0.13 | 0.70 | 1.22 | 8.259E-11 | 1479 | 117441 | 118920 |
| 378 | sense organs | Strabismus and other disorders of binocular eye movements | 1.76 | 0.57 | 0.08 | 0.40 | 0.72 | 8.908E-11 | 5908 | 112428 | 118336 |
| 740.9 | musculoskeletal | Osteoarthrosis NOS | 1.35 | 0.30 | 0.05 | 0.21 | 0.39 | 1.576E-10 | 45367 | 63386 | 108753 |
| 681 | dermatologic | Superficial cellulitis and abscess | 1.42 | 0.35 | 0.05 | 0.24 | 0.45 | 1.589E-10 | 20903 | 86945 | 107848 |
| 278.1 | endocrine/metabolic | Obesity | 1.34 | 0.29 | 0.05 | 0.20 | 0.38 | 1.731E-10 | 45115 | 66768 | 111883 |
| 700 | dermatologic | Corns and callosities | 1.56 | 0.44 | 0.07 | 0.31 | 0.57 | 1.993E-10 | 10936 | 104661 | 115597 |
| 689 | dermatologic | Disorder of skin and subcutaneous tissue NOS | 1.48 | 0.39 | 0.06 | 0.27 | 0.51 | 2.356E-10 | 15614 | 86497 | 102111 |
| 512 | respiratory | Other symptoms of respiratory system | 1.35 | 0.30 | 0.05 | 0.21 | 0.40 | 2.498E-10 | 54986 | 46937 | 101923 |
| 280.1 | hematopoietic | Iron deficiency anemias, unspecified or not due to blood loss | 1.55 | 0.44 | 0.07 | 0.30 | 0.56 | 2.581E-10 | 11400 | 104555 | 115955 |

| | | | | | | | | | | | |
|---|---|---|---|---|---|---|---|---|---|---|---|
| 761 | symptoms | Cervicalgia | 1.41 | 0.35 | 0.05 | 0.24 | 0.45 | 2.742E-10 | 19418 | 91856 | 111274 |
| 456 | circulatory system | Chronic venous insufficiency [CVI] | 1.58 | 0.46 | 0.07 | 0.32 | 0.59 | 3.383E-10 | 9809 | 106259 | 116068 |
| 706 | dermatologic | Diseases of sebaceous glands | 1.42 | 0.35 | 0.05 | 0.24 | 0.46 | 3.437E-10 | 21089 | 84178 | 105267 |
| 798 | symptoms | Malaise and fatigue | 1.40 | 0.33 | 0.05 | 0.23 | 0.44 | 4.350E-10 | 24447 | 80371 | 104818 |
| 368.9 | sense organs | Subjective visual disturbances | 2.03 | 0.71 | 0.10 | 0.50 | 0.90 | 4.828E-10 | 3216 | 112205 | 115421 |
| 916 | injuries & poisonings | Contusion | 1.68 | 0.52 | 0.08 | 0.36 | 0.67 | 7.987E-10 | 6505 | 102183 | 108688 |
| 840 | injuries & poisonings | Sprains and strains | 1.51 | 0.41 | 0.06 | 0.28 | 0.54 | 8.257E-10 | 11179 | 97067 | 108246 |
| 599 | genitourinary | Other symptoms/disorders or the urinary system | 1.37 | 0.31 | 0.05 | 0.21 | 0.41 | 8.413E-10 | 30441 | 73867 | 104308 |
| 276 | endocrine/metabolic | Disorders of fluid, electrolyte, and acid-base balance | 1.39 | 0.33 | 0.05 | 0.22 | 0.43 | 8.710E-10 | 24830 | 80987 | 105817 |
| 454 | circulatory system | Varicose veins | 1.69 | 0.52 | 0.08 | 0.36 | 0.68 | 9.056E-10 | 6285 | 110695 | 116980 |
| 512.8 | respiratory | Cough | 1.39 | 0.33 | 0.05 | 0.23 | 0.43 | 9.743E-10 | 23444 | 77809 | 101253 |
| 726.1 | musculoskeletal | Enthesopathy | 1.49 | 0.40 | 0.06 | 0.27 | 0.52 | 1.162E-09 | 11960 | 98002 | 109962 |
| 250.23 | endocrine/metabolic | Type 2 diabetes with ophthalmic manifestations | 1.55 | 0.44 | 0.07 | 0.30 | 0.57 | 1.165E-09 | 10434 | 104176 | 114610 |
| 721 | musculoskeletal | Spondylosis and allied disorders | 1.42 | 0.35 | 0.06 | 0.24 | 0.46 | 1.685E-09 | 17069 | 92875 | 109944 |
| 365.11 | sense organs | Primary open angle glaucoma | 1.54 | 0.43 | 0.07 | 0.30 | 0.57 | 2.046E-09 | 11133 | 108166 | 119299 |
| 687.4 | dermatologic | Disturbance of skin sensation | 1.54 | 0.43 | 0.07 | 0.29 | 0.57 | 2.592E-09 | 9126 | 99514 | 108640 |
| 512.9 | respiratory | Other dyspnea | 1.37 | 0.32 | 0.05 | 0.21 | 0.42 | 2.993E-09 | 24292 | 79982 | 104274 |
| 706.8 | dermatologic | Other specified diseases of sebaceous glands | 1.58 | 0.46 | 0.07 | 0.31 | 0.60 | 3.079E-09 | 9257 | 101366 | 110623 |
| 278 | endocrine/metabolic | Overweight, obesity and other hyperalimentation | 1.31 | 0.27 | 0.05 | 0.18 | 0.36 | 3.991E-09 | 50173 | 59481 | 109654 |
| 497 | respiratory | Bronchitis | 1.68 | 0.52 | 0.08 | 0.35 | 0.68 | 4.189E-09 | 5906 | 105309 | 111215 |
| 716 | musculoskeletal | Other arthropathies | 1.44 | 0.37 | 0.06 | 0.25 | 0.48 | 4.449E-09 | 14534 | 94598 | 109132 |
| 979 | injuries & poisonings | Adverse drug events and drug allergies | 2.22 | 0.80 | 0.12 | 0.55 | 1.03 | 6.346E-09 | 1921 | 113705 | 115626 |
| 611 | genitourinary | Abnormal findings on mammogram or breast exam | 2.09 | 0.74 | 0.12 | 0.50 | 0.97 | 6.936E-09 | 1862 | 118891 | 120753 |
| 370.1 | sense organs | Corneal ulcer | 3.84 | 1.35 | 0.19 | 0.94 | 1.71 | 7.834E-09 | 439 | 121801 | 122240 |

| | | | | | | | | | | | |
|---|---|---|---|---|---|---|---|---|---|---|---|
| 362.3 | sense organs | Other nondiabetic retinopathy | 1.75 | 0.56 | 0.09 | 0.38 | 0.74 | 8.646E-09 | 4901 | 111633 | 116534 |
| 367.8 | sense organs | Hypermetropia | 1.33 | 0.29 | 0.05 | 0.19 | 0.38 | 9.031E-09 | 32148 | 68931 | 101079 |
| 366.1 | sense organs | Nonsenile Cataract | 2.51 | 0.92 | 0.14 | 0.63 | 1.19 | 9.288E-09 | 1484 | 118241 | 119725 |
| 401.1 | circulatory system | Essential hypertension | 1.50 | 0.41 | 0.07 | 0.26 | 0.56 | 9.495E-09 | 104215 | 13636 | 117851 |
| 401 | circulatory system | Hypertension | 1.51 | 0.41 | 0.08 | 0.27 | 0.56 | 9.613E-09 | 104773 | 13248 | 118021 |
| 250.7 | endocrine/metabolic | Diabetic retinopathy | 1.50 | 0.41 | 0.07 | 0.27 | 0.54 | 1.021E-08 | 10994 | 103839 | 114833 |
| 939 | dermatologic | Atopic/contact dermatitis due to other or unspecified | 1.37 | 0.32 | 0.05 | 0.21 | 0.42 | 1.261E-08 | 21679 | 82761 | 104440 |
| 401.2 | circulatory system | Hypertensive heart and/or renal disease | 1.36 | 0.30 | 0.05 | 0.20 | 0.41 | 1.394E-08 | 24955 | 86028 | 110983 |
| 735.2 | musculoskeletal | Acquired toe deformities | 1.47 | 0.39 | 0.07 | 0.26 | 0.51 | 1.461E-08 | 11791 | 102852 | 114643 |
| 721.1 | musculoskeletal | Spondylosis without myelopathy | 1.40 | 0.34 | 0.06 | 0.22 | 0.45 | 1.578E-08 | 15752 | 94543 | 110295 |
| 703.1 | dermatologic | Ingrowing nail | 1.54 | 0.43 | 0.07 | 0.29 | 0.57 | 2.022E-08 | 8553 | 108301 | 116854 |
| 386.9 | sense organs | Dizziness and giddiness (Light-headedness and vertigo) | 1.36 | 0.31 | 0.05 | 0.20 | 0.42 | 2.273E-08 | 22063 | 84012 | 106075 |
| 703 | dermatologic | Diseases of nail, NOS | 1.41 | 0.34 | 0.06 | 0.23 | 0.46 | 2.473E-08 | 16919 | 95935 | 112854 |
| 562.1 | digestive | Diverticulosis | 1.37 | 0.31 | 0.06 | 0.20 | 0.42 | 5.464E-08 | 20174 | 82390 | 102564 |
| 427 | circulatory system | Cardiac dysrhythmias | 1.29 | 0.26 | 0.05 | 0.16 | 0.35 | 5.800E-08 | 53831 | 53484 | 107315 |
| 454.11 | circulatory system | Varicose veins of lower extremity, symptomtic | 1.82 | 0.60 | 0.10 | 0.39 | 0.79 | 7.454E-08 | 3489 | 115702 | 119191 |
| 454.1 | circulatory system | Varicose veins of lower extremity | 1.62 | 0.49 | 0.08 | 0.32 | 0.65 | 7.820E-08 | 5746 | 112198 | 117944 |
| 585.1 | genitourinary | Acute renal failure | 1.39 | 0.33 | 0.06 | 0.21 | 0.45 | 8.135E-08 | 15762 | 97809 | 113571 |
| 340 | neurological | Migraine | 1.69 | 0.52 | 0.09 | 0.34 | 0.70 | 8.610E-08 | 3689 | 116069 | 119758 |
| 707 | dermatologic | Chronic ulcer of skin | 1.50 | 0.41 | 0.07 | 0.26 | 0.55 | 1.016E-07 | 9251 | 107328 | 116579 |
| 368.4 | sense organs | Visual field defects | 1.80 | 0.59 | 0.10 | 0.38 | 0.78 | 1.072E-07 | 3786 | 113811 | 117597 |
| 687.1 | dermatologic | Rash and other nonspecific skin eruption | 1.41 | 0.34 | 0.06 | 0.22 | 0.46 | 1.078E-07 | 13670 | 89465 | 103135 |
| 599.5 | genitourinary | Frequency of urination and polyuria | 1.44 | 0.36 | 0.07 | 0.23 | 0.49 | 1.123E-07 | 12781 | 96330 | 109111 |
| 600 | genitourinary | Hyperplasia of prostate | 1.29 | 0.25 | 0.05 | 0.16 | 0.35 | 1.526E-07 | 56522 | 51954 | 108476 |

| | | | | | | | | | | | |
|---|---|---|---|---|---|---|---|---|---|---|---|
| 870 | injuries & poisonings | Open wounds of head; neck; and trunk | 1.72 | 0.54 | 0.10 | 0.35 | 0.73 | 1.741E-07 | 4086 | 110706 | 114792 |
| 276.5 | endocrine/metabolic | Hypovolemia | 1.54 | 0.43 | 0.08 | 0.27 | 0.58 | 1.747E-07 | 7703 | 103868 | 111571 |
| 327.3 | neurological | Sleep apnea | 1.27 | 0.24 | 0.05 | 0.15 | 0.33 | 1.781E-07 | 39640 | 77404 | 117044 |
| 745 | musculoskeletal | Pain in joint | 1.31 | 0.27 | 0.05 | 0.17 | 0.38 | 1.813E-07 | 74592 | 32592 | 107184 |
| 369.5 | sense organs | Conjunctivitis, infectious | 1.86 | 0.62 | 0.11 | 0.40 | 0.83 | 2.294E-07 | 2959 | 111911 | 114870 |
| 369.2 | sense organs | Eye infection, viral | 2.44 | 0.89 | 0.15 | 0.58 | 1.18 | 2.303E-07 | 1170 | 120008 | 121178 |
| 740 | musculoskeletal | Osteoarthrosis | 1.27 | 0.24 | 0.05 | 0.15 | 0.33 | 2.546E-07 | 60249 | 48271 | 108520 |
| 698 | dermatologic | Pruritus and related conditions | 1.70 | 0.53 | 0.10 | 0.34 | 0.71 | 2.625E-07 | 4329 | 110913 | 115242 |
| 41 | infectious diseases | Bacterial infection NOS | 1.50 | 0.40 | 0.07 | 0.25 | 0.55 | 2.671E-07 | 8206 | 104618 | 112824 |
| 78 | infectious diseases | Viral warts & HPV | 1.60 | 0.47 | 0.09 | 0.30 | 0.63 | 2.773E-07 | 5430 | 108886 | 114316 |
| 379.51 | sense organs | Pigmentary iris degeneration | 3.56 | 1.27 | 0.21 | 0.83 | 1.66 | 2.838E-07 | 410 | 121858 | 122268 |
| 871 | injuries & poisonings | Open wounds of extremities | 1.50 | 0.40 | 0.07 | 0.25 | 0.55 | 2.933E-07 | 7948 | 104376 | 112324 |
| 512.7 | respiratory | Shortness of breath | 1.29 | 0.25 | 0.05 | 0.16 | 0.35 | 3.159E-07 | 31417 | 73338 | 104755 |
| 41.1 | infectious diseases | Staphylococcus infections | 1.88 | 0.63 | 0.11 | 0.40 | 0.85 | 3.421E-07 | 2564 | 116521 | 119085 |
| 562 | digestive | Diverticulosis and diverticulitis | 1.33 | 0.28 | 0.05 | 0.18 | 0.39 | 3.565E-07 | 22539 | 80102 | 102641 |
| 741 | musculoskeletal | Symptoms and disorders of the joints | 1.38 | 0.32 | 0.06 | 0.20 | 0.44 | 3.595E-07 | 14647 | 93169 | 107816 |
| 356 | neurological | Hereditary and idiopathic peripheral neuropathy | 1.40 | 0.33 | 0.06 | 0.21 | 0.46 | 3.747E-07 | 12999 | 100652 | 113651 |
| 381 | sense organs | Otitis media and Eustachian tube disorders | 1.54 | 0.43 | 0.08 | 0.27 | 0.59 | 3.791E-07 | 6422 | 107549 | 113971 |
| 727 | musculoskeletal | Other disorders of synovium, tendon, and bursa | 1.35 | 0.30 | 0.06 | 0.19 | 0.41 | 4.272E-07 | 16256 | 94212 | 110468 |
| 250.2 | endocrine/metabolic | Type 2 diabetes | 1.25 | 0.22 | 0.04 | 0.14 | 0.31 | 4.549E-07 | 53490 | 63101 | 116591 |
| 272.1 | endocrine/metabolic | Hyperlipidemia | 1.43 | 0.35 | 0.07 | 0.21 | 0.50 | 6.371E-07 | 103981 | 13783 | 117764 |
| 728.7 | musculoskeletal | Fasciitis | 1.42 | 0.35 | 0.07 | 0.22 | 0.48 | 6.545E-07 | 10158 | 105490 | 115648 |
| 338.2 | neurological | Chronic pain | 1.39 | 0.33 | 0.06 | 0.20 | 0.45 | 7.936E-07 | 11444 | 100487 | 111931 |
| 272 | endocrine/metabolic | Disorders of lipoid metabolism | 1.42 | 0.35 | 0.07 | 0.21 | 0.50 | 8.168E-07 | 104044 | 13725 | 117769 |
| 788 | symptoms | Syncope and collapse | 1.40 | 0.34 | 0.07 | 0.20 | 0.46 | 9.086E-07 | 11893 | 102156 | 114049 |

| | | | | | | | | | | | |
|---|---|---|---|---|---|---|---|---|---|---|---|
| 250.14 | endocrine/metabolic | Type 1 diabetes with neurological manifestations | 2.45 | 0.89 | 0.16 | 0.56 | 1.20 | 9.773E-07 | 1000 | 120341 | 121341 |
| 250 | endocrine/metabolic | Diabetes mellitus | 1.24 | 0.22 | 0.04 | 0.13 | 0.31 | 9.840E-07 | 53756 | 62405 | 116161 |
| 381.1 | sense organs | Otitis media | 1.63 | 0.49 | 0.09 | 0.30 | 0.67 | 1.050E-06 | 4329 | 111206 | 115535 |
| 627 | genitourinary | Menopausal and postmenopausal disorders | 2.30 | 0.83 | 0.18 | 0.49 | 1.18 | 1.110E-06 | 1210 | 1269 | 2479 |
| 627.2 | genitourinary | Symptomatic menopause | 2.29 | 0.83 | 0.17 | 0.50 | 1.16 | 1.201E-06 | 843 | 1696 | 2539 |
| 401.22 | circulatory system | Hypertensive chronic kidney disease | 1.38 | 0.32 | 0.06 | 0.19 | 0.44 | 1.430E-06 | 14519 | 100783 | 115302 |
| 374 | sense organs | Other disorders of eyelids | 1.35 | 0.30 | 0.06 | 0.18 | 0.42 | 1.492E-06 | 16258 | 95037 | 111295 |
| 361.1 | sense organs | Retinal detachment with retinal defect | 1.97 | 0.68 | 0.13 | 0.42 | 0.92 | 1.560E-06 | 2048 | 119331 | 121379 |
| 521 | digestive | Diseases of hard tissues of teeth | 1.30 | 0.27 | 0.05 | 0.16 | 0.37 | 1.695E-06 | 18950 | 96620 | 115570 |
| 337.1 | neurological | Peripheral autonomic neuropathy | 1.59 | 0.47 | 0.09 | 0.28 | 0.64 | 2.236E-06 | 4762 | 113045 | 117807 |
| 361 | sense organs | Retinal detachments and defects | 1.55 | 0.44 | 0.09 | 0.26 | 0.61 | 2.335E-06 | 5582 | 114063 | 119645 |
| 585 | genitourinary | Renal failure | 1.26 | 0.23 | 0.05 | 0.14 | 0.33 | 2.361E-06 | 34646 | 78521 | 113167 |
| 495 | respiratory | Asthma | 1.42 | 0.35 | 0.07 | 0.21 | 0.49 | 2.497E-06 | 8739 | 109485 | 118224 |
| 426 | circulatory system | Cardiac conduction disorders | 1.29 | 0.25 | 0.05 | 0.15 | 0.36 | 2.550E-06 | 26102 | 80587 | 106689 |
| 722 | musculoskeletal | Intervertebral disc disorders | 1.30 | 0.26 | 0.05 | 0.15 | 0.37 | 2.599E-06 | 19825 | 90433 | 110258 |
| 681.1 | dermatologic | Cellulitis and abscess of fingers/toes | 1.71 | 0.54 | 0.11 | 0.32 | 0.74 | 2.661E-06 | 3360 | 113890 | 117250 |
| 702.2 | dermatologic | Seborrheic keratosis | 1.26 | 0.23 | 0.05 | 0.13 | 0.32 | 2.791E-06 | 37465 | 65820 | 103285 |
| 741.3 | musculoskeletal | Difficulty in walking | 1.44 | 0.36 | 0.07 | 0.21 | 0.50 | 2.917E-06 | 8929 | 106016 | 114945 |
| 687 | dermatologic | Symptoms affecting skin | 1.61 | 0.48 | 0.10 | 0.28 | 0.66 | 2.986E-06 | 4428 | 106598 | 111026 |
| 521.1 | digestive | Dental caries | 1.30 | 0.26 | 0.05 | 0.15 | 0.37 | 3.107E-06 | 18455 | 97695 | 116150 |
| 337 | neurological | Disorders of the autonomic nervous system | 1.56 | 0.45 | 0.09 | 0.26 | 0.62 | 3.210E-06 | 5097 | 112365 | 117462 |
| 611.1 | genitourinary | Abnormal mammogram | 2.41 | 0.88 | 0.18 | 0.52 | 1.23 | 3.317E-06 | 517 | 121411 | 121928 |
| 301 | mental disorders | Personality disorders | 1.81 | 0.59 | 0.12 | 0.35 | 0.82 | 3.910E-06 | 2002 | 119107 | 121109 |
| 300.4 | mental disorders | Dysthymic disorder | 1.46 | 0.38 | 0.08 | 0.22 | 0.53 | 4.042E-06 | 6643 | 112021 | 118664 |
| 480 | respiratory | Pneumonia | 1.33 | 0.28 | 0.06 | 0.17 | 0.40 | 4.092E-06 | 15979 | 97283 | 113262 |

| | | | | | | | | | | | |
|---|---|---|---|---|---|---|---|---|---|---|---|
| 535 | digestive | Gastritis and duodenitis | 1.52 | 0.42 | 0.09 | 0.25 | 0.59 | 4.581E-06 | 5810 | 106969 | 112779 |
| 327.32 | neurological | Obstructive sleep apnea | 1.25 | 0.22 | 0.05 | 0.13 | 0.31 | 4.796E-06 | 32976 | 84134 | 117110 |
| 379.9 | sense organs | Pain, swelling or discharge of eye | 2.28 | 0.83 | 0.16 | 0.49 | 1.13 | 5.178E-06 | 1056 | 116430 | 117486 |
| 208 | neoplasms | Benign neoplasm of colon | 1.24 | 0.22 | 0.05 | 0.12 | 0.31 | 5.496E-06 | 41585 | 64840 | 106425 |
| 523 | digestive | Gingival and periodontal diseases | 1.31 | 0.27 | 0.06 | 0.15 | 0.38 | 5.702E-06 | 15827 | 101005 | 116832 |
| 766 | symptoms | Neuralgia, neuritis, and radiculitis NOS | 1.69 | 0.52 | 0.11 | 0.31 | 0.73 | 5.746E-06 | 3065 | 114319 | 117384 |
| 272.12 | endocrine/metabolic | Hyperglyceridemia | 1.60 | 0.47 | 0.10 | 0.27 | 0.66 | 5.939E-06 | 4105 | 114694 | 118799 |
| 292.2 | mental disorders | Mild cognitive impairment | 1.47 | 0.38 | 0.08 | 0.22 | 0.54 | 6.029E-06 | 7473 | 110452 | 117925 |
| 789 | symptoms | Nausea and vomiting | 1.45 | 0.37 | 0.08 | 0.21 | 0.52 | 6.110E-06 | 7103 | 105207 | 112310 |
| 611.3 | genitourinary | Lump or mass in breast | 1.89 | 0.64 | 0.13 | 0.37 | 0.89 | 6.575E-06 | 1429 | 119490 | 120919 |
| 251.1 | endocrine/metabolic | Hypoglycemia | 2.05 | 0.72 | 0.14 | 0.42 | 0.99 | 6.904E-06 | 1512 | 118279 | 119791 |
| 420 | circulatory system | Carditis | 1.76 | 0.57 | 0.12 | 0.33 | 0.79 | 6.944E-06 | 2720 | 116598 | 119318 |
| 525 | digestive | Other diseases of the teeth and supporting structures | 1.28 | 0.24 | 0.05 | 0.14 | 0.35 | 6.950E-06 | 20453 | 94517 | 114970 |
| 536.3 | digestive | Gastroparesis | 2.73 | 1.00 | 0.19 | 0.60 | 1.37 | 7.770E-06 | 580 | 121430 | 122010 |
| 757 | congenital anomalies | Congenital anomalies of the integument | 3.46 | 1.24 | 0.23 | 0.75 | 1.67 | 8.340E-06 | 352 | 121564 | 121916 |
| 411 | circulatory system | Ischemic Heart Disease | 1.23 | 0.21 | 0.05 | 0.12 | 0.30 | 8.823E-06 | 54296 | 58220 | 112516 |
| 591 | genitourinary | Urinary tract infection | 1.31 | 0.27 | 0.06 | 0.15 | 0.39 | 9.851E-06 | 17042 | 93861 | 110903 |
| 842 | injuries & poisonings | Other sprains and strains | 2.59 | 0.95 | 0.19 | 0.56 | 1.31 | 1.008E-05 | 663 | 118463 | 119126 |
| 290.3 | mental disorders | Other persistent mental disorders due to conditions classified elsewhere | 1.51 | 0.41 | 0.09 | 0.23 | 0.58 | 1.085E-05 | 6093 | 113088 | 119181 |
| 300.1 | mental disorders | Anxiety disorder | 1.26 | 0.23 | 0.05 | 0.13 | 0.33 | 1.099E-05 | 22979 | 90253 | 113232 |
| 368.1 | sense organs | Amblyopia | 1.76 | 0.57 | 0.12 | 0.32 | 0.79 | 1.146E-05 | 2538 | 118759 | 121297 |
| 70 | infectious diseases | Viral hepatitis | 0.55 | -0.60 | 0.15 | -0.91 | -0.32 | 1.211E-05 | 3162 | 118116 | 121278 |
| 378.1 | sense organs | Strabismus (not specified as paralytic) | 1.59 | 0.46 | 0.10 | 0.26 | 0.66 | 1.226E-05 | 4135 | 115045 | 119180 |
| 573.7 | digestive | Abnormal results of function study of liver | 1.46 | 0.38 | 0.08 | 0.21 | 0.54 | 1.278E-05 | 6023 | 111052 | 117075 |
| 292 | mental disorders | Neurological disorders | 1.29 | 0.25 | 0.06 | 0.14 | 0.36 | 1.302E-05 | 19671 | 92491 | 112162 |

| | | | | | | | | | | | |
|---|---|---|---|---|---|---|---|---|---|---|---|
| 707.2 | dermatologic | Chronic ulcer of leg or foot | 1.49 | 0.40 | 0.09 | 0.22 | 0.56 | 1.306E-05 | 6087 | 113127 | 119214 |
| 427.3 | circulatory system | Other specified cardiac dysrhythmias | 1.31 | 0.27 | 0.06 | 0.15 | 0.39 | 1.308E-05 | 17367 | 90151 | 107518 |
| 475 | respiratory | Chronic sinusitis | 1.38 | 0.32 | 0.07 | 0.18 | 0.46 | 1.390E-05 | 9620 | 102273 | 111893 |
| 70.3 | infectious diseases | Viral hepatitis C | 0.53 | -0.64 | 0.16 | -0.97 | -0.34 | 1.535E-05 | 2666 | 119101 | 121767 |
| 763 | symptoms | Thoracic or lumbosacral neuritis or radiculitis, unspecified | 1.34 | 0.29 | 0.07 | 0.16 | 0.42 | 1.555E-05 | 11349 | 104020 | 115369 |
| 524 | digestive | Dentofacial anomalies, including malocclusion | 1.96 | 0.67 | 0.14 | 0.38 | 0.94 | 1.686E-05 | 1467 | 118345 | 119812 |
| 249 | endocrine/metabolic | Secondary diabetes mellitus | 1.82 | 0.60 | 0.13 | 0.34 | 0.84 | 1.908E-05 | 2076 | 117359 | 119435 |
| 465.2 | respiratory | Acute pharyngitis | 1.69 | 0.53 | 0.11 | 0.29 | 0.74 | 1.954E-05 | 2655 | 111613 | 114268 |
| 578 | digestive | Gastrointestinal hemorrhage | 1.31 | 0.27 | 0.06 | 0.15 | 0.39 | 2.011E-05 | 14712 | 96996 | 111708 |
| 276.1 | endocrine/metabolic | Electrolyte imbalance | 1.29 | 0.25 | 0.06 | 0.14 | 0.36 | 2.077E-05 | 18228 | 90445 | 108673 |
| 250.25 | endocrine/metabolic | Diabetes type 2 with peripheral circulatory disorders | 1.61 | 0.48 | 0.10 | 0.26 | 0.67 | 2.084E-05 | 3908 | 115335 | 119243 |
| 690 | dermatologic | Erythematosquamous dermatosis | 1.34 | 0.29 | 0.07 | 0.16 | 0.42 | 2.352E-05 | 12292 | 100918 | 113210 |
| 296.2 | mental disorders | Depression | 1.22 | 0.20 | 0.05 | 0.10 | 0.28 | 2.382E-05 | 39139 | 76015 | 115154 |
| 470 | respiratory | Septal Deviations/Turbinate Hypertrophy | 1.66 | 0.51 | 0.11 | 0.28 | 0.72 | 2.401E-05 | 2906 | 116470 | 119376 |
| 691 | dermatologic | Congenital anomalies of skin | 2.50 | 0.92 | 0.19 | 0.52 | 1.27 | 2.504E-05 | 674 | 120538 | 121212 |
| 416 | circulatory system | Cardiomegaly | 1.59 | 0.46 | 0.10 | 0.25 | 0.66 | 2.688E-05 | 4051 | 110686 | 114737 |
| 296 | mental disorders | Mood disorders | 1.21 | 0.19 | 0.05 | 0.10 | 0.28 | 2.850E-05 | 41389 | 73845 | 115234 |
| 415.2 | circulatory system | Chronic pulmonary heart disease | 1.53 | 0.43 | 0.10 | 0.23 | 0.61 | 2.882E-05 | 4762 | 113049 | 117811 |
| 278.11 | endocrine/metabolic | Morbid obesity | 1.34 | 0.29 | 0.07 | 0.16 | 0.42 | 2.896E-05 | 10659 | 106757 | 117416 |
| 216 | neoplasms | Benign neoplasm of skin | 1.32 | 0.28 | 0.06 | 0.15 | 0.40 | 3.172E-05 | 13117 | 92391 | 105508 |
| 272.11 | endocrine/metabolic | Hypercholesterolemia | 1.26 | 0.23 | 0.05 | 0.12 | 0.33 | 3.816E-05 | 22503 | 89481 | 111984 |
| 327 | neurological | Sleep disorders | 1.23 | 0.20 | 0.05 | 0.11 | 0.30 | 4.068E-05 | 29688 | 77604 | 107292 |
| 690.1 | dermatologic | Seborrheic dermatitis | 1.33 | 0.29 | 0.07 | 0.15 | 0.42 | 4.247E-05 | 12228 | 101182 | 113410 |
| 459.9 | circulatory system | Circulatory disease NEC | 1.52 | 0.42 | 0.10 | 0.22 | 0.60 | 4.313E-05 | 4938 | 108864 | 113802 |
| 426.21 | circulatory system | First degree AV block | 1.68 | 0.52 | 0.12 | 0.28 | 0.74 | 4.356E-05 | 3071 | 114931 | 118002 |

| | | | | | | | | | | | |
|---|---|---|---|---|---|---|---|---|---|---|---|
| 574 | digestive | Cholelithiasis and cholecystitis | 1.41 | 0.34 | 0.08 | 0.18 | 0.50 | 4.490E-05 | 7327 | 111334 | 118661 |
| 938.2 | injuries & poisonings | Chronic dermatitis due to solar radiation | 1.64 | 0.49 | 0.11 | 0.27 | 0.71 | 4.599E-05 | 3164 | 114837 | 118001 |
| 522 | digestive | Diseases of pulp and periapical tissues | 1.50 | 0.41 | 0.09 | 0.22 | 0.59 | 4.684E-05 | 4355 | 113909 | 118264 |
| 596 | genitourinary | Other disorders of bladder | 1.34 | 0.30 | 0.07 | 0.16 | 0.43 | 5.014E-05 | 10900 | 103323 | 114223 |
| 574.1 | digestive | Cholelithiasis | 1.46 | 0.38 | 0.09 | 0.20 | 0.55 | 5.091E-05 | 5647 | 112870 | 118517 |
| 743 | musculoskeletal | Osteoporosis, osteopenia and pathological fracture | 1.31 | 0.27 | 0.07 | 0.14 | 0.40 | 5.590E-05 | 13693 | 101754 | 115447 |
| 370.2 | sense organs | Superficial keratitis | 2.42 | 0.88 | 0.19 | 0.48 | 1.25 | 5.791E-05 | 700 | 119850 | 120550 |
| 735.1 | musculoskeletal | Flat foot | 1.53 | 0.42 | 0.10 | 0.22 | 0.62 | 6.759E-05 | 4173 | 114697 | 118870 |
| 395.1 | circulatory system | Nonrheumatic mitral valve disorders | 1.49 | 0.40 | 0.10 | 0.21 | 0.58 | 7.484E-05 | 5010 | 111619 | 116629 |
| 707.3 | dermatologic | Chronic ulcer of unspecified site | 1.88 | 0.63 | 0.15 | 0.33 | 0.90 | 7.496E-05 | 1636 | 117950 | 119586 |
| 257.1 | endocrine/metabolic | Testicular hypofunction | 1.44 | 0.36 | 0.09 | 0.19 | 0.53 | 7.550E-05 | 5938 | 111647 | 117585 |
| 458 | circulatory system | Hypotension | 1.29 | 0.26 | 0.06 | 0.13 | 0.38 | 7.863E-05 | 15179 | 94036 | 109215 |
| 578.2 | digestive | Blood in stool | 1.41 | 0.35 | 0.08 | 0.18 | 0.51 | 8.057E-05 | 6813 | 106600 | 113413 |
| 303 | mental disorders | Psychogenic and somatoform disorders | 1.77 | 0.57 | 0.13 | 0.30 | 0.83 | 8.604E-05 | 1579 | 119138 | 120717 |
| 599.4 | genitourinary | Urinary incontinence | 1.33 | 0.28 | 0.07 | 0.14 | 0.42 | 8.720E-05 | 10578 | 104158 | 114736 |
| 433 | circulatory system | Cerebrovascular disease | 1.23 | 0.21 | 0.05 | 0.10 | 0.31 | 8.799E-05 | 27958 | 83343 | 111301 |
| 447.1 | circulatory system | Stricture of artery | 3.00 | 1.10 | 0.24 | 0.59 | 1.54 | 9.098E-05 | 371 | 120740 | 121111 |
| 743.9 | musculoskeletal | Osteopenia or other disorder of bone and cartilage | 1.41 | 0.34 | 0.08 | 0.17 | 0.51 | 9.228E-05 | 6450 | 110863 | 117313 |
| 228.1 | neoplasms | Hemangioma of skin and subcutaneous tissue | 1.67 | 0.52 | 0.12 | 0.27 | 0.75 | 9.487E-05 | 2570 | 114216 | 116786 |
| 296.1 | mental disorders | Bipolar | 1.52 | 0.42 | 0.10 | 0.21 | 0.62 | 9.670E-05 | 3231 | 118196 | 121427 |
| 726.4 | musculoskeletal | Calcaneal spur; Exostosis NOS | 1.60 | 0.47 | 0.11 | 0.24 | 0.69 | 9.833E-05 | 3050 | 113878 | 116928 |
| 257 | endocrine/metabolic | Testicular dysfunction | 1.42 | 0.35 | 0.09 | 0.18 | 0.52 | 9.915E-05 | 6113 | 110952 | 117065 |
| 564 | digestive | Functional digestive disorders | 1.40 | 0.33 | 0.08 | 0.17 | 0.49 | 9.955E-05 | 6417 | 106682 | 113099 |
| 523.3 | digestive | Periodontitis (acute or chronic) | 1.34 | 0.29 | 0.07 | 0.15 | 0.43 | 1.080E-04 | 8615 | 108880 | 117495 |

| | | | | | | | | | | | |
|---|---|---|---|---|---|---|---|---|---|---|---|
| 938 | injuries & poisonings | Dermatitis due to solar radiation | 1.48 | 0.39 | 0.10 | 0.20 | 0.58 | 1.149E-04 | 4822 | 110489 | 115311 |
| 626 | genitourinary | Disorders of menstruation and other abnormal bleeding from female genital tract | 3.12 | 1.14 | 0.27 | 0.58 | 1.65 | 1.236E-04 | 121 | 2639 | 2760 |
| 809 | injuries & poisonings | Fracture of unspecified bones | 1.76 | 0.57 | 0.14 | 0.29 | 0.82 | 1.272E-04 | 1835 | 117922 | 119757 |
| 522.1 | digestive | Pulpitis and necrosis of tooth pulp | 1.62 | 0.48 | 0.12 | 0.24 | 0.71 | 1.275E-04 | 2542 | 117248 | 119790 |
| 464 | respiratory | Acute sinusitis | 1.41 | 0.34 | 0.09 | 0.17 | 0.51 | 1.297E-04 | 5621 | 106071 | 111692 |
| 605 | genitourinary | Erectile dysfunction [ED] | 1.21 | 0.19 | 0.05 | 0.09 | 0.28 | 1.459E-04 | 35361 | 73704 | 109065 |
| 296.22 | mental disorders | Major depressive disorder | 1.21 | 0.19 | 0.05 | 0.09 | 0.28 | 1.479E-04 | 28331 | 87316 | 115647 |
| 694 | dermatologic | Dyschromia and Vitiligo | 1.44 | 0.36 | 0.09 | 0.18 | 0.54 | 1.485E-04 | 5336 | 108148 | 113484 |
| 772.3 | symptoms | Muscle weakness | 1.29 | 0.25 | 0.06 | 0.12 | 0.38 | 1.596E-04 | 13981 | 98064 | 112045 |
| 565 | digestive | Anal and rectal conditions | 1.51 | 0.41 | 0.10 | 0.20 | 0.61 | 1.630E-04 | 3979 | 111457 | 115436 |
| 509 | respiratory | Respiratory failure, insufficiency, arrest | 1.30 | 0.27 | 0.07 | 0.13 | 0.40 | 1.682E-04 | 11385 | 104671 | 116056 |
| 736.6 | musculoskeletal | Unequal leg length (acquired) | 1.87 | 0.62 | 0.15 | 0.31 | 0.91 | 1.718E-04 | 1422 | 119883 | 121305 |
| 377.3 | sense organs | Optic neuritis/neuropathy | 1.79 | 0.58 | 0.14 | 0.29 | 0.85 | 1.722E-04 | 1779 | 118834 | 120613 |
| 287 | hematopoietic | Purpura and other hemorrhagic conditions | 1.37 | 0.32 | 0.08 | 0.16 | 0.47 | 1.723E-04 | 7503 | 108822 | 116325 |
| 735.21 | musculoskeletal | Hammer toe (acquired) | 1.37 | 0.32 | 0.08 | 0.15 | 0.47 | 1.734E-04 | 7632 | 109648 | 117280 |
| 380.4 | sense organs | Impacted cerumen | 1.24 | 0.21 | 0.06 | 0.10 | 0.32 | 1.809E-04 | 23058 | 83003 | 106061 |
| 428 | circulatory system | Congestive heart failure; nonhypertensive | 1.23 | 0.20 | 0.05 | 0.10 | 0.31 | 1.872E-04 | 24575 | 90264 | 114839 |
| 41.12 | infectious diseases | Methicillin resistant Staphylococcus aureus | 1.98 | 0.68 | 0.17 | 0.34 | 0.99 | 1.893E-04 | 1094 | 119810 | 120904 |
| 428.4 | circulatory system | Heart failure with preserved EF [Diastolic heart failure] | 1.35 | 0.30 | 0.08 | 0.15 | 0.45 | 1.898E-04 | 8751 | 108690 | 117441 |
| 281.9 | hematopoietic | Deficiency anemias | 1.91 | 0.65 | 0.16 | 0.32 | 0.94 | 1.934E-04 | 1398 | 118792 | 120190 |
| 550.2 | digestive | Diaphragmatic hernia | 1.40 | 0.33 | 0.09 | 0.16 | 0.50 | 1.942E-04 | 6532 | 108302 | 114834 |
| 428.1 | circulatory system | Congestive heart failure (CHF) NOS | 1.26 | 0.23 | 0.06 | 0.11 | 0.35 | 1.953E-04 | 17769 | 97454 | 115223 |
| 374.6 | sense organs | Dermatochalasis | 1.37 | 0.32 | 0.08 | 0.15 | 0.48 | 1.978E-04 | 7506 | 109050 | 116556 |

| | | | | | | | | | | | |
|---|---|---|---|---|---|---|---|---|---|---|---|
| 722.6 | musculoskeletal | Degeneration of intervertebral disc | 1.25 | 0.23 | 0.06 | 0.11 | 0.34 | 2.116E-04 | 15626 | 95420 | 111046 |
| 764 | symptoms | Sciatica | 1.38 | 0.32 | 0.08 | 0.15 | 0.48 | 2.165E-04 | 6546 | 108866 | 115412 |
| 395 | circulatory system | Heart valve disorders | 1.28 | 0.25 | 0.06 | 0.12 | 0.37 | 2.167E-04 | 14663 | 98928 | 113591 |
| 303.4 | mental disorders | Somatoform disorder | 1.77 | 0.57 | 0.14 | 0.28 | 0.84 | 2.193E-04 | 1365 | 119928 | 121293 |
| 382 | sense organs | Otalgia | 1.70 | 0.53 | 0.13 | 0.26 | 0.79 | 2.243E-04 | 1937 | 115041 | 116978 |
| 458.9 | circulatory system | Hypotension NOS | 1.35 | 0.30 | 0.08 | 0.14 | 0.45 | 2.292E-04 | 8697 | 103251 | 111948 |
| 483 | respiratory | Acute bronchitis and bronchiolitis | 1.31 | 0.27 | 0.07 | 0.13 | 0.41 | 2.298E-04 | 10096 | 97020 | 107116 |
| 720 | musculoskeletal | Spinal stenosis | 1.27 | 0.24 | 0.06 | 0.11 | 0.36 | 2.342E-04 | 14083 | 101752 | 115835 |
| 381.2 | sense organs | Eustachian tube disorders | 1.69 | 0.52 | 0.13 | 0.25 | 0.77 | 2.374E-04 | 1980 | 117403 | 119383 |
| 244.1 | endocrine/metabolic | Secondary hypothyroidism | 1.89 | 0.64 | 0.16 | 0.31 | 0.94 | 2.445E-04 | 1146 | 120836 | 121982 |
| 596.5 | genitourinary | Functional disorders of bladder | 1.41 | 0.34 | 0.09 | 0.16 | 0.52 | 2.452E-04 | 5600 | 112708 | 118308 |
| 558 | digestive | Noninfectious gastroenteritis | 1.51 | 0.41 | 0.11 | 0.20 | 0.62 | 2.587E-04 | 3477 | 112640 | 116117 |
| 514 | respiratory | Abnormal findings examination of lungs | 1.28 | 0.24 | 0.07 | 0.11 | 0.37 | 2.630E-04 | 13407 | 97845 | 111252 |
| 300.3 | mental disorders | Obsessive-compulsive disorders | 2.12 | 0.75 | 0.19 | 0.37 | 1.10 | 2.639E-04 | 754 | 121225 | 121979 |
| 694.2 | dermatologic | Other dyschromia | 1.44 | 0.37 | 0.10 | 0.17 | 0.55 | 2.707E-04 | 4784 | 108978 | 113762 |
| 790.6 | symptoms | Other abnormal blood chemistry | 1.35 | 0.30 | 0.08 | 0.14 | 0.45 | 2.857E-04 | 8324 | 101090 | 109414 |
| 525.1 | digestive | Loss of teeth or edentulism | 1.25 | 0.23 | 0.06 | 0.10 | 0.34 | 2.923E-04 | 14295 | 103943 | 118238 |
| 701.1 | dermatologic | Keratoderma, acquired | 1.45 | 0.37 | 0.10 | 0.17 | 0.55 | 2.949E-04 | 4955 | 110050 | 115005 |
| 772.2 | symptoms | Spasm of muscle | 1.61 | 0.47 | 0.12 | 0.22 | 0.71 | 3.063E-04 | 2234 | 114954 | 117188 |
| 411.4 | circulatory system | Coronary atherosclerosis | 1.18 | 0.17 | 0.05 | 0.08 | 0.26 | 3.082E-04 | 48814 | 65062 | 113876 |
| 381.11 | sense organs | Suppurative and unspecified otitis media | 1.57 | 0.45 | 0.12 | 0.21 | 0.67 | 3.095E-04 | 2812 | 113953 | 116765 |
| 426.3 | circulatory system | Bundle branch block | 1.41 | 0.35 | 0.09 | 0.16 | 0.52 | 3.148E-04 | 5816 | 110543 | 116359 |
| 713 | musculoskeletal | Arthropathy associated with other disorders classified elsewhere | 2.38 | 0.87 | 0.21 | 0.42 | 1.26 | 3.170E-04 | 586 | 121457 | 122043 |
| 368.2 | sense organs | Diplopia and disorders of binocular vision | 1.49 | 0.40 | 0.10 | 0.19 | 0.59 | 3.192E-04 | 4168 | 114259 | 118427 |

| | | | | | | | | | | | |
|---|---|---|---|---|---|---|---|---|---|---|---|
| 380.1 | sense organs | Otitis externa | 1.44 | 0.36 | 0.10 | 0.17 | 0.55 | 3.234E-04 | 4947 | 111686 | 116633 |
| 427.7 | circulatory system | Tachycardia NOS | 1.47 | 0.39 | 0.10 | 0.18 | 0.58 | 3.244E-04 | 3934 | 110941 | 114875 |
| 522.5 | digestive | Periapical abscess | 1.87 | 0.62 | 0.16 | 0.30 | 0.93 | 3.346E-04 | 1208 | 118075 | 119283 |
| 427.9 | circulatory system | Palpitations | 1.39 | 0.33 | 0.09 | 0.15 | 0.49 | 3.380E-04 | 5449 | 110493 | 115942 |
| 564.1 | digestive | Irritable Bowel Syndrome | 1.55 | 0.44 | 0.12 | 0.20 | 0.66 | 3.424E-04 | 2466 | 118163 | 120629 |
| 304 | mental disorders | Adjustment reaction | 1.25 | 0.22 | 0.06 | 0.10 | 0.34 | 3.544E-04 | 14532 | 99128 | 113660 |
| 250.22 | endocrine/metabolic | Type 2 diabetes with renal manifestations | 1.32 | 0.28 | 0.08 | 0.13 | 0.43 | 3.802E-04 | 9593 | 108288 | 117881 |
| 355.1 | neurological | Chronic pain syndrome | 1.41 | 0.34 | 0.09 | 0.16 | 0.52 | 3.901E-04 | 4543 | 114440 | 118983 |
| 947 | injuries & poisonings | Urticaria | 1.82 | 0.60 | 0.15 | 0.28 | 0.89 | 3.954E-04 | 1338 | 118508 | 119846 |
| 426.7 | circulatory system | Abnormal electrocardiogram [ECG] [EKG] | 1.29 | 0.26 | 0.07 | 0.12 | 0.39 | 3.973E-04 | 10985 | 97344 | 108329 |
| 523.1 | digestive | Gingivitis | 1.36 | 0.31 | 0.08 | 0.14 | 0.47 | 4.087E-04 | 6185 | 112367 | 118552 |
| 277 | endocrine/metabolic | Other disorders of metabolism | 1.78 | 0.58 | 0.15 | 0.27 | 0.86 | 4.187E-04 | 1467 | 118498 | 119965 |
| 704.8 | dermatologic | Other specified diseases of hair and hair follicles | 1.66 | 0.50 | 0.13 | 0.23 | 0.76 | 4.289E-04 | 2156 | 115661 | 117817 |
| 285.2 | hematopoietic | Anemia of chronic disease | 1.38 | 0.33 | 0.09 | 0.15 | 0.50 | 4.477E-04 | 6582 | 110720 | 117302 |
| 736 | musculoskeletal | Other acquired deformities of limbs | 1.52 | 0.42 | 0.11 | 0.19 | 0.63 | 4.505E-04 | 3134 | 115465 | 118599 |
| 371.2 | sense organs | Conjunctivitis, noninfectious | 1.52 | 0.42 | 0.11 | 0.19 | 0.63 | 4.514E-04 | 3107 | 114779 | 117886 |
| 459 | circulatory system | Other disorders of circulatory system | 1.38 | 0.32 | 0.09 | 0.15 | 0.50 | 4.857E-04 | 6348 | 106031 | 112379 |
| 722.9 | musculoskeletal | Other and unspecified disc disorder | 1.80 | 0.59 | 0.15 | 0.27 | 0.88 | 4.890E-04 | 1357 | 118278 | 119635 |
| 172 | neoplasms | Skin cancer | 1.19 | 0.17 | 0.05 | 0.08 | 0.27 | 4.968E-04 | 36108 | 79463 | 115571 |
| 281 | hematopoietic | Other deficiency anemia | 1.37 | 0.32 | 0.09 | 0.14 | 0.48 | 4.969E-04 | 6360 | 110739 | 117099 |
| 597 | genitourinary | Other disorders of urethra and urinary tract | 1.70 | 0.53 | 0.14 | 0.24 | 0.80 | 4.994E-04 | 1954 | 118862 | 120816 |
| 962 | injuries & poisonings | Poisoning by hormones and synthetic substitutes | 2.68 | 0.98 | 0.24 | 0.46 | 1.44 | 5.343E-04 | 380 | 120638 | 121018 |
| 599.2 | genitourinary | Retention of urine | 1.29 | 0.25 | 0.07 | 0.11 | 0.39 | 5.348E-04 | 11661 | 102211 | 113872 |
| 379.1 | sense organs | Scleritis and episcleritis | 2.77 | 1.02 | 0.25 | 0.48 | 1.49 | 5.524E-04 | 333 | 121919 | 122252 |

| | | | | | | | | | | | |
|---|---|---|---|---|---|---|---|---|---|---|---|
| 571.5 | digestive | Other chronic nonalcoholic liver disease | 1.35 | 0.30 | 0.08 | 0.13 | 0.46 | 5.668E-04 | 6075 | 111931 | 118006 |
| 427.22 | circulatory system | Atrial flutter | 1.30 | 0.26 | 0.07 | 0.11 | 0.40 | 5.796E-04 | 10182 | 107777 | 117959 |
| 287.31 | hematopoietic | Primary thrombocytopenia | 2.38 | 0.87 | 0.22 | 0.40 | 1.28 | 5.894E-04 | 532 | 121524 | 122056 |
| 420.3 | circulatory system | Endocarditis | 1.85 | 0.61 | 0.16 | 0.28 | 0.92 | 6.050E-04 | 1343 | 119427 | 120770 |
| 447 | circulatory system | Other disorders of arteries and arterioles | 1.77 | 0.57 | 0.15 | 0.25 | 0.86 | 6.065E-04 | 1569 | 117066 | 118635 |
| 585.3 | genitourinary | Chronic renal failure [CKD] | 1.20 | 0.19 | 0.05 | 0.08 | 0.29 | 6.220E-04 | 26630 | 89237 | 115867 |
| 571 | digestive | Chronic liver disease and cirrhosis | 1.34 | 0.29 | 0.08 | 0.13 | 0.45 | 6.355E-04 | 6443 | 111309 | 117752 |
| 704 | dermatologic | Diseases of hair and hair follicles | 1.51 | 0.41 | 0.12 | 0.18 | 0.63 | 7.125E-04 | 3017 | 113451 | 116468 |
| 550 | digestive | Abdominal hernia | 1.20 | 0.19 | 0.05 | 0.08 | 0.29 | 7.509E-04 | 23044 | 88767 | 111811 |
| 528 | digestive | Diseases of the oral soft tissues, excluding lesions specific for gingiva and tongue | 1.49 | 0.40 | 0.11 | 0.17 | 0.62 | 7.916E-04 | 3169 | 113284 | 116453 |
| 365.2 | sense organs | Primary angle-closure glaucoma | 1.49 | 0.40 | 0.11 | 0.17 | 0.61 | 7.920E-04 | 3512 | 116904 | 120416 |
| 755 | congenital anomalies | Congenital anomalies of limbs | 1.48 | 0.39 | 0.11 | 0.17 | 0.61 | 8.137E-04 | 3218 | 115546 | 118764 |
| 443 | circulatory system | Peripheral vascular disease | 1.23 | 0.21 | 0.06 | 0.09 | 0.33 | 8.178E-04 | 16931 | 95924 | 112855 |
| 911 | injuries & poisonings | Blister | 2.16 | 0.77 | 0.21 | 0.34 | 1.16 | 8.860E-04 | 686 | 119832 | 120518 |
| 536 | digestive | Disorders of function of stomach | 1.48 | 0.39 | 0.11 | 0.17 | 0.60 | 8.965E-04 | 3373 | 114086 | 117459 |
| 713.5 | musculoskeletal | Arthropathy associated with neurological disorders | 2.40 | 0.88 | 0.23 | 0.39 | 1.31 | 9.091E-04 | 484 | 121852 | 122336 |
| 426.32 | circulatory system | Left bundle branch block | 1.60 | 0.47 | 0.13 | 0.20 | 0.73 | 9.699E-04 | 2287 | 117326 | 119613 |
| 753 | congenital anomalies | Congenital anomalies of the eye | 2.52 | 0.93 | 0.24 | 0.40 | 1.38 | 9.722E-04 | 426 | 120883 | 121309 |
| 426.91 | circulatory system | Cardiac pacemaker in situ | 1.32 | 0.27 | 0.08 | 0.11 | 0.43 | 9.727E-04 | 9007 | 111363 | 120370 |
| 613 | genitourinary | Other nonmalignant breast conditions | 1.71 | 0.54 | 0.15 | 0.22 | 0.83 | 9.918E-04 | 1021 | 119735 | 120756 |
| 395.3 | circulatory system | Nonrheumatic tricuspid valve disorders | 2.08 | 0.73 | 0.20 | 0.31 | 1.10 | 9.957E-04 | 776 | 119005 | 119781 |

| | | | | | | | | | | | |
|---|---|---|---|---|---|---|---|---|---|---|---|
| 371.21 | sense organs | Allergic conjunctivitis | 1.51 | 0.41 | 0.12 | 0.17 | 0.64 | 1.013E-03 | 2777 | 115541 | 118318 |
| 8 | infectious diseases | Intestinal infection | 1.52 | 0.42 | 0.12 | 0.17 | 0.65 | 1.016E-03 | 2777 | 115419 | 118196 |
| 327.1 | neurological | Hypersomnia | 2.00 | 0.69 | 0.19 | 0.29 | 1.05 | 1.023E-03 | 722 | 119400 | 120122 |
| 715 | musculoskeletal | Other inflammatory spondylopathies | 1.64 | 0.49 | 0.14 | 0.21 | 0.76 | 1.080E-03 | 1737 | 118881 | 120618 |
| 377 | sense organs | Disorders of optic nerve and visual pathways | 1.42 | 0.35 | 0.10 | 0.14 | 0.54 | 1.080E-03 | 4549 | 113628 | 118177 |
| 597.1 | genitourinary | Urethral stricture (not specified as infectious) | 1.69 | 0.53 | 0.15 | 0.22 | 0.81 | 1.089E-03 | 1774 | 119461 | 121235 |
| 428.3 | circulatory system | Heart failure with reduced EF [Systolic or combined heart failure] | 1.25 | 0.23 | 0.07 | 0.09 | 0.36 | 1.108E-03 | 13132 | 104365 | 117497 |
| 90 | infectious diseases | Sexually transmitted infections (not HIV or hepatitis) | 2.27 | 0.82 | 0.22 | 0.35 | 1.23 | 1.147E-03 | 529 | 121239 | 121768 |
| 411.8 | circulatory system | Other chronic ischemic heart disease, unspecified | 1.18 | 0.17 | 0.05 | 0.07 | 0.27 | 1.181E-03 | 29341 | 85460 | 114801 |
| 508 | respiratory | Pulmonary collapse; interstitial and compensatory emphysema | 1.51 | 0.41 | 0.12 | 0.17 | 0.64 | 1.226E-03 | 2920 | 113111 | 116031 |
| 300.11 | mental disorders | Generalized anxiety disorder | 1.33 | 0.28 | 0.09 | 0.11 | 0.45 | 1.244E-03 | 5796 | 113608 | 119404 |
| 495.1 | respiratory | Chronic obstructive asthma | 1.78 | 0.58 | 0.16 | 0.24 | 0.88 | 1.257E-03 | 1307 | 119668 | 120975 |
| 287.3 | hematopoietic | Thrombocytopenia | 1.33 | 0.29 | 0.09 | 0.11 | 0.45 | 1.271E-03 | 6830 | 111033 | 117863 |
| 365.5 | sense organs | Pseudoexfoliation glaucoma | 2.11 | 0.74 | 0.21 | 0.31 | 1.13 | 1.275E-03 | 749 | 121473 | 122222 |
| 433.31 | circulatory system | Transient cerebral ischemia | 1.26 | 0.23 | 0.07 | 0.09 | 0.36 | 1.282E-03 | 12354 | 103695 | 116049 |
| 523.31 | digestive | Acute periodontitis | 1.34 | 0.29 | 0.09 | 0.12 | 0.46 | 1.288E-03 | 5544 | 113060 | 118604 |
| 261 | endocrine/metabolic | Vitamin deficiency | 1.18 | 0.17 | 0.05 | 0.07 | 0.27 | 1.313E-03 | 26046 | 84853 | 110899 |
| 608 | genitourinary | Other disorders of male genital organs | 1.51 | 0.41 | 0.12 | 0.16 | 0.64 | 1.342E-03 | 2949 | 111842 | 114791 |
| 681.5 | dermatologic | Cellulitis and abscess of leg, except foot | 1.34 | 0.30 | 0.09 | 0.12 | 0.47 | 1.355E-03 | 6075 | 110636 | 116711 |
| 715.1 | musculoskeletal | Sacroiliitis NEC | 1.77 | 0.57 | 0.16 | 0.23 | 0.88 | 1.362E-03 | 1141 | 120215 | 121356 |
| 250.13 | endocrine/metabolic | Type 1 diabetes with ophthalmic manifestations | 2.22 | 0.80 | 0.22 | 0.33 | 1.21 | 1.366E-03 | 551 | 120931 | 121482 |
| 38 | infectious diseases | Septicemia | 1.31 | 0.27 | 0.08 | 0.11 | 0.43 | 1.385E-03 | 7560 | 110000 | 117560 |

| | | | | | | | | | | | |
|---|---|---|---|---|---|---|---|---|---|---|---|
| 386 | sense organs | Vertiginous syndromes and other disorders of vestibular system | 1.35 | 0.30 | 0.09 | 0.12 | 0.47 | 1.408E-03 | 5774 | 110833 | 116607 |
| 297 | mental disorders | Suicidal ideation or attempt | 1.48 | 0.39 | 0.12 | 0.15 | 0.61 | 1.427E-03 | 2580 | 116925 | 119505 |
| 727.1 | musculoskeletal | Synovitis and tenosynovitis | 1.30 | 0.27 | 0.08 | 0.10 | 0.42 | 1.463E-03 | 7072 | 109879 | 116951 |
| 599.9 | genitourinary | Other abnormality of urination | 1.35 | 0.30 | 0.09 | 0.12 | 0.47 | 1.464E-03 | 5912 | 107415 | 113327 |
| 425.1 | circulatory system | Primary/intrinsic cardiomyopathies | 1.32 | 0.27 | 0.08 | 0.11 | 0.43 | 1.485E-03 | 7519 | 110437 | 117956 |
| 479 | respiratory | Other upper respiratory disease | 1.39 | 0.33 | 0.10 | 0.13 | 0.52 | 1.583E-03 | 4422 | 108347 | 112769 |
| 276.12 | endocrine/metabolic | Hyposmolality and/or hyponatremia | 1.35 | 0.30 | 0.09 | 0.12 | 0.48 | 1.650E-03 | 5290 | 111729 | 117019 |
| 755.1 | congenital anomalies | Congenital deformities of feet | 1.46 | 0.38 | 0.11 | 0.15 | 0.59 | 1.671E-03 | 3081 | 116030 | 119111 |
| 875 | injuries & poisonings | Non-healing surgical wound | 1.87 | 0.63 | 0.18 | 0.25 | 0.97 | 1.671E-03 | 962 | 120374 | 121336 |
| 374.3 | sense organs | Ptosis of eyelid | 1.37 | 0.32 | 0.10 | 0.12 | 0.50 | 1.679E-03 | 5312 | 113473 | 118785 |
| 250.4 | endocrine/metabolic | Abnormal glucose | 1.19 | 0.17 | 0.05 | 0.07 | 0.28 | 1.687E-03 | 21187 | 88818 | 110005 |
| 930 | injuries & poisonings | Allergic reaction to food | 3.10 | 1.13 | 0.30 | 0.46 | 1.70 | 1.716E-03 | 194 | 121835 | 122029 |
| 286.9 | hematopoietic | Abnormal coagulation profile | 1.67 | 0.51 | 0.15 | 0.20 | 0.80 | 1.796E-03 | 1683 | 117907 | 119590 |
| 379.4 | sense organs | Anomalies of pupillary function | 2.02 | 0.70 | 0.20 | 0.28 | 1.08 | 1.880E-03 | 741 | 120422 | 121163 |
| 228 | neoplasms | Hemangioma and lymphangioma, any site | 1.42 | 0.35 | 0.11 | 0.13 | 0.56 | 1.890E-03 | 3877 | 111129 | 115006 |
| 41.11 | infectious diseases | Methicillin sensitive Staphylococcus aureus | 1.85 | 0.61 | 0.18 | 0.24 | 0.95 | 1.895E-03 | 1016 | 119977 | 120993 |
| 707.1 | dermatologic | Decubitus ulcer | 1.45 | 0.37 | 0.11 | 0.14 | 0.59 | 1.940E-03 | 3587 | 115927 | 119514 |
| 532 | digestive | Dysphagia | 1.19 | 0.18 | 0.06 | 0.07 | 0.29 | 1.945E-03 | 20567 | 93407 | 113974 |
| 443.7 | circulatory system | Peripheral angiopathy in diseases classified elsewhere | 3.39 | 1.22 | 0.33 | 0.49 | 1.82 | 2.016E-03 | 184 | 122103 | 122287 |
| 702 | dermatologic | Degenerative skin conditions and other dermatoses | 1.15 | 0.14 | 0.05 | 0.05 | 0.24 | 2.027E-03 | 55037 | 51766 | 106803 |
| 510 | respiratory | Other diseases of lung | 1.30 | 0.26 | 0.08 | 0.10 | 0.42 | 2.038E-03 | 7681 | 106675 | 114356 |
| 710.12 | musculoskeletal | Chronic osteomyelitis | 1.70 | 0.53 | 0.16 | 0.20 | 0.83 | 2.124E-03 | 1390 | 120420 | 121810 |
| 433.3 | circulatory system | Cerebral ischemia | 1.24 | 0.21 | 0.07 | 0.08 | 0.34 | 2.171E-03 | 12985 | 102342 | 115327 |

| | | | | | | | | | | | |
|---|---|---|---|---|---|---|---|---|---|---|---|
| 425 | circulatory system | Cardiomyopathy | 1.30 | 0.26 | 0.08 | 0.10 | 0.42 | 2.183E-03 | 7832 | 109904 | 117736 |
| 523.32 | digestive | Chronic periodontitis | 1.34 | 0.29 | 0.09 | 0.11 | 0.47 | 2.198E-03 | 5170 | 113755 | 118925 |
| 965.1 | injuries & poisonings | Opiates and related narcotics causing adverse effects in therapeutic use | 2.72 | 1.00 | 0.28 | 0.39 | 1.52 | 2.209E-03 | 263 | 120966 | 121229 |
| 411.1 | circulatory system | Unstable angina (intermediate coronary syndrome) | 1.34 | 0.29 | 0.09 | 0.11 | 0.47 | 2.259E-03 | 5797 | 111038 | 116835 |
| 722.1 | musculoskeletal | Displacement of intervertebral disc | 1.42 | 0.35 | 0.11 | 0.13 | 0.56 | 2.316E-03 | 3420 | 114331 | 117751 |
| 771.1 | symptoms | Swelling of limb | 1.45 | 0.37 | 0.12 | 0.14 | 0.59 | 2.317E-03 | 3193 | 110596 | 113789 |
| 727.4 | musculoskeletal | Ganglion and cyst of synovium, tendon, and bursa | 1.53 | 0.42 | 0.13 | 0.16 | 0.67 | 2.402E-03 | 2114 | 117733 | 119847 |
| 362.8 | sense organs | Retinal hemorrhage/ischemia | 1.52 | 0.42 | 0.13 | 0.15 | 0.67 | 2.426E-03 | 2558 | 115395 | 117953 |
| 300.12 | mental disorders | Agorophobia, social phobia, and panic disorder | 1.46 | 0.38 | 0.12 | 0.14 | 0.60 | 2.474E-03 | 2493 | 118612 | 121105 |
| 580.3 | genitourinary | Nephritis and nephropathy without mention of glomerulonephritis | 1.72 | 0.54 | 0.16 | 0.20 | 0.85 | 2.482E-03 | 1378 | 119557 | 120935 |
| 580 | genitourinary | Nephritis; nephrosis; renal sclerosis | 1.59 | 0.46 | 0.14 | 0.17 | 0.73 | 2.534E-03 | 1972 | 118190 | 120162 |
| 172.2 | neoplasms | Other non-epithelial cancer of skin | 1.17 | 0.16 | 0.05 | 0.05 | 0.26 | 2.708E-03 | 30336 | 84118 | 114454 |
| 79 | infectious diseases | Viral infection | 1.67 | 0.51 | 0.16 | 0.19 | 0.81 | 2.748E-03 | 1398 | 115716 | 117114 |
| 395.2 | circulatory system | Nonrheumatic aortic valve disorders | 1.27 | 0.24 | 0.08 | 0.08 | 0.39 | 2.784E-03 | 9777 | 107413 | 117190 |
| 272.13 | endocrine/metabolic | Mixed hyperlipidemia | 1.16 | 0.15 | 0.05 | 0.05 | 0.25 | 2.857E-03 | 29565 | 82016 | 111581 |
| 353 | neurological | Nerve root and plexus disorders | 1.64 | 0.50 | 0.16 | 0.18 | 0.79 | 3.010E-03 | 1408 | 118747 | 120155 |
| 742 | musculoskeletal | Derangement of joint, non-traumatic | 1.54 | 0.43 | 0.14 | 0.15 | 0.69 | 3.130E-03 | 1851 | 116295 | 118146 |
| 457 | circulatory system | Encounter for long-term (current) use of anticoagulants, antithrombotics, aspirin | 1.46 | 0.38 | 0.12 | 0.13 | 0.60 | 3.235E-03 | 3214 | 113576 | 116790 |

| | | | | | | | | | | | |
|---|---|---|---|---|---|---|---|---|---|---|---|
| 288 | hematopoietic | Diseases of white blood cells | 1.32 | 0.28 | 0.09 | 0.09 | 0.45 | 3.245E-03 | 5760 | 108753 | 114513 |
| 765 | symptoms | Cervical radiculitis | 1.32 | 0.28 | 0.09 | 0.10 | 0.46 | 3.259E-03 | 5124 | 112979 | 118103 |
| 394 | circulatory system | Rheumatic disease of the heart valves | 1.50 | 0.40 | 0.13 | 0.14 | 0.65 | 3.261E-03 | 2731 | 114344 | 117075 |
| 360.3 | sense organs | Hypotony of eye | 3.76 | 1.32 | 0.36 | 0.49 | 1.99 | 3.291E-03 | 135 | 122275 | 122410 |
| 81 | infectious diseases | Infection/inflammation of internal prosthetic device; implant; and graft | 1.84 | 0.61 | 0.19 | 0.21 | 0.96 | 3.331E-03 | 914 | 120589 | 121503 |
| 585.33 | genitourinary | Chronic Kidney Disease, Stage III | 1.20 | 0.18 | 0.06 | 0.06 | 0.30 | 3.391E-03 | 18321 | 98972 | 117293 |
| 601 | genitourinary | Inflammatory diseases of prostate | 1.32 | 0.27 | 0.09 | 0.09 | 0.45 | 3.425E-03 | 6493 | 105836 | 112329 |
| 737 | musculoskeletal | Curvature of spine | 1.60 | 0.47 | 0.15 | 0.16 | 0.75 | 3.448E-03 | 1558 | 118053 | 119611 |
| 443.9 | circulatory system | Peripheral vascular disease, unspecified | 1.21 | 0.19 | 0.06 | 0.06 | 0.31 | 3.488E-03 | 16051 | 97248 | 113299 |
| 415 | circulatory system | Pulmonary heart disease | 1.27 | 0.24 | 0.08 | 0.08 | 0.39 | 3.619E-03 | 8617 | 108759 | 117376 |
| 426.2 | circulatory system | Atrioventricular [AV] block | 1.30 | 0.26 | 0.09 | 0.09 | 0.43 | 3.651E-03 | 7509 | 109714 | 117223 |
| 507 | respiratory | Pleurisy; pleural effusion | 1.30 | 0.26 | 0.09 | 0.09 | 0.43 | 3.656E-03 | 6872 | 109118 | 115990 |
| 599.3 | genitourinary | Dysuria | 1.47 | 0.39 | 0.13 | 0.13 | 0.62 | 3.747E-03 | 2566 | 113302 | 115868 |
| 695.7 | dermatologic | Prurigo and Lichen | 1.49 | 0.40 | 0.13 | 0.13 | 0.65 | 3.750E-03 | 2301 | 116677 | 118978 |
| 720.1 | musculoskeletal | Spinal stenosis of lumbar region | 1.24 | 0.22 | 0.07 | 0.07 | 0.36 | 4.016E-03 | 10057 | 107327 | 117384 |
| 598 | genitourinary | Abnormal findings on examination of urine | 1.52 | 0.42 | 0.14 | 0.14 | 0.68 | 4.019E-03 | 2187 | 115335 | 117522 |
| 110.12 | infectious diseases | Althete's foot | 1.32 | 0.27 | 0.09 | 0.09 | 0.45 | 4.041E-03 | 6022 | 108217 | 114239 |
| 513 | respiratory | Respiratory abnormalities | 1.41 | 0.34 | 0.11 | 0.11 | 0.56 | 4.079E-03 | 3523 | 113287 | 116810 |
| 580.32 | genitourinary | Nephritis and nephropathy with pathological lesion | 2.10 | 0.74 | 0.23 | 0.25 | 1.17 | 4.312E-03 | 535 | 121297 | 121832 |
| 313.2 | mental disorders | Tics and stuttering | 3.16 | 1.15 | 0.34 | 0.40 | 1.78 | 4.343E-03 | 158 | 122217 | 122375 |
| 261.2 | endocrine/metabolic | Vitamin B-complex deficiencies | 1.27 | 0.24 | 0.08 | 0.08 | 0.39 | 4.356E-03 | 8180 | 109728 | 117908 |
| 740.1 | musculoskeletal | Osteoarthritis; localized | 1.15 | 0.14 | 0.05 | 0.04 | 0.23 | 4.563E-03 | 35456 | 72432 | 107888 |
| 729 | musculoskeletal | Other disorders of soft tissues | 1.54 | 0.43 | 0.14 | 0.14 | 0.70 | 4.566E-03 | 1781 | 115355 | 117136 |
| 427.6 | circulatory system | Premature beats | 1.39 | 0.33 | 0.11 | 0.10 | 0.54 | 4.566E-03 | 3864 | 111975 | 115839 |

| | | | | | | | | | | | |
|---|---|---|---|---|---|---|---|---|---|---|---|
| 8.6 | infectious diseases | Viral Enteritis | 2.28 | 0.83 | 0.26 | 0.27 | 1.31 | 4.807E-03 | 361 | 120453 | 120814 |
| 290 | mental disorders | Delirium dementia and amnestic and other cognitive disorders | 1.21 | 0.19 | 0.07 | 0.06 | 0.32 | 4.815E-03 | 15375 | 100027 | 115402 |
| 624.9 | genitourinary | stress incontinence, female | 1.90 | 0.64 | 0.22 | 0.20 | 1.06 | 4.842E-03 | 323 | 2350 | 2673 |
| 340.1 | neurological | Migrain with aura | 1.95 | 0.67 | 0.22 | 0.21 | 1.07 | 4.908E-03 | 507 | 121128 | 121635 |
| 386.2 | sense organs | Peripheral or central vertigo | 1.34 | 0.29 | 0.10 | 0.09 | 0.48 | 4.960E-03 | 4834 | 112405 | 117239 |
| 286 | hematopoietic | Coagulation defects | 1.36 | 0.30 | 0.10 | 0.09 | 0.50 | 4.966E-03 | 4572 | 113628 | 118200 |
| 840.2 | injuries & poisonings | Rotator cuff (capsule) sprain | 1.44 | 0.36 | 0.12 | 0.11 | 0.59 | 4.998E-03 | 2922 | 116616 | 119538 |
| 702.1 | dermatologic | Actinic keratosis | 1.15 | 0.14 | 0.05 | 0.04 | 0.23 | 5.025E-03 | 43026 | 66412 | 109438 |
| 594.3 | genitourinary | Calculus of ureter | 1.43 | 0.36 | 0.12 | 0.11 | 0.59 | 5.169E-03 | 2757 | 118124 | 120881 |
| 907 | injuries & poisonings | Injuries to the nervous system | 1.65 | 0.50 | 0.17 | 0.16 | 0.81 | 5.231E-03 | 1262 | 119307 | 120569 |
| 41.4 | infectious diseases | E. coli | 1.73 | 0.55 | 0.18 | 0.17 | 0.89 | 5.323E-03 | 1035 | 119082 | 120117 |
| 870.3 | injuries & poisonings | Other open wound of head and face | 1.82 | 0.60 | 0.20 | 0.19 | 0.96 | 5.403E-03 | 889 | 118798 | 119687 |
| 440 | circulatory system | Atherosclerosis | 1.27 | 0.24 | 0.08 | 0.07 | 0.40 | 5.451E-03 | 8044 | 104805 | 112849 |
| 185 | neoplasms | Cancer of prostate | 0.83 | -0.18 | 0.07 | -0.32 | -0.05 | 5.635E-03 | 18812 | 98420 | 117232 |
| 871.3 | injuries & poisonings | Open wound of foot except toe(s) alone | 1.64 | 0.49 | 0.17 | 0.15 | 0.81 | 5.851E-03 | 1308 | 119137 | 120445 |
| 286.2 | hematopoietic | Encounter for long-term (current) use of anticoagulants | 1.19 | 0.17 | 0.06 | 0.05 | 0.29 | 5.893E-03 | 17393 | 101927 | 119320 |
| 372 | sense organs | Disorders of conjunctiva | 1.36 | 0.31 | 0.11 | 0.09 | 0.51 | 6.058E-03 | 4309 | 110905 | 115214 |
| 513.3 | respiratory | Hypoventilation | 1.71 | 0.54 | 0.18 | 0.16 | 0.87 | 6.226E-03 | 1085 | 118893 | 119978 |
| 333 | neurological | Extrapyramidal disease and abnormal movement disorders | 1.26 | 0.23 | 0.08 | 0.07 | 0.39 | 6.414E-03 | 7519 | 110730 | 118249 |
| 738 | musculoskeletal | Other acquired musculoskeletal deformity | 1.45 | 0.37 | 0.13 | 0.11 | 0.62 | 6.432E-03 | 2277 | 116348 | 118625 |
| 695 | dermatologic | Erythematous conditions | 1.22 | 0.20 | 0.07 | 0.06 | 0.34 | 6.434E-03 | 10526 | 102220 | 112746 |
| 931 | injuries & poisonings | Contact dermatitis and other eczema due to plants [except food] | 2.17 | 0.78 | 0.25 | 0.23 | 1.24 | 6.445E-03 | 413 | 120518 | 120931 |
| 112 | infectious diseases | Candidiasis | 1.45 | 0.37 | 0.13 | 0.11 | 0.61 | 6.582E-03 | 2398 | 115060 | 117458 |

| | | | | | | | | | | | |
|---|---|---|---|---|---|---|---|---|---|---|---|
| 250.42 | endocrine/metabolic | Other abnormal glucose | 1.23 | 0.21 | 0.07 | 0.06 | 0.35 | 6.691E-03 | 9490 | 102428 | 111918 |
| 199 | neoplasms | Neoplasm of uncertain behavior | 1.27 | 0.24 | 0.09 | 0.07 | 0.41 | 6.745E-03 | 6844 | 104945 | 111789 |
| 215 | neoplasms | Other benign neoplasm of connective and other soft tissue | 1.93 | 0.66 | 0.22 | 0.19 | 1.07 | 6.765E-03 | 592 | 119852 | 120444 |
| 10 | infectious diseases | Tuberculosis | 2.15 | 0.76 | 0.25 | 0.23 | 1.23 | 6.828E-03 | 440 | 121492 | 121932 |
| 333.2 | neurological | Myoclonus | 2.21 | 0.80 | 0.26 | 0.24 | 1.28 | 6.935E-03 | 385 | 121692 | 122077 |
| 385 | sense organs | Other disorders of middle ear and mastoid | 1.98 | 0.68 | 0.23 | 0.20 | 1.10 | 6.985E-03 | 577 | 121231 | 121808 |
| 429 | circulatory system | Ill-defined descriptions and complications of heart disease | 1.32 | 0.28 | 0.10 | 0.08 | 0.47 | 7.028E-03 | 5103 | 106503 | 111606 |
| 293.1 | mental disorders | Swelling, mass, or lump in head and neck [Space-occupying lesion, intracranial NOS] | 1.41 | 0.34 | 0.12 | 0.10 | 0.57 | 7.143E-03 | 2876 | 114930 | 117806 |
| 420.2 | circulatory system | Pericarditis | 1.62 | 0.48 | 0.17 | 0.14 | 0.79 | 7.234E-03 | 1327 | 119626 | 120953 |
| 740.3 | musculoskeletal | Osteoarthrosis involving more than one site, but not specified as generalized | 1.53 | 0.43 | 0.15 | 0.12 | 0.71 | 7.327E-03 | 1878 | 117998 | 119876 |
| 613.8 | genitourinary | Other specified disorders of breast | 2.43 | 0.89 | 0.30 | 0.25 | 1.46 | 7.418E-03 | 130 | 122004 | 122134 |
| 841 | injuries & poisonings | Sprains and strains of back and neck | 1.51 | 0.41 | 0.14 | 0.11 | 0.68 | 7.472E-03 | 1734 | 116314 | 118048 |
| 353.1 | neurological | Nerve plexus lesions | 2.53 | 0.93 | 0.30 | 0.27 | 1.48 | 7.490E-03 | 254 | 121930 | 122184 |
| 427.1 | circulatory system | Paroxysmal tachycardia, unspecified | 1.29 | 0.25 | 0.09 | 0.07 | 0.43 | 7.637E-03 | 5900 | 111749 | 117649 |
| 728 | musculoskeletal | Disorders of muscle, ligament, and fascia | 2.19 | 0.79 | 0.26 | 0.22 | 1.27 | 7.664E-03 | 375 | 120980 | 121355 |
| 117.4 | infectious diseases | Aspergillosis | 3.25 | 1.18 | 0.36 | 0.35 | 1.85 | 7.782E-03 | 140 | 122300 | 122440 |
| 742.9 | musculoskeletal | Other derangement of joint | 1.54 | 0.43 | 0.15 | 0.12 | 0.72 | 7.808E-03 | 1480 | 117399 | 118879 |
| 531 | digestive | Peptic ulcer (excl. esophageal) | 1.30 | 0.26 | 0.09 | 0.07 | 0.44 | 7.879E-03 | 5712 | 112901 | 118613 |
| 574.3 | digestive | Cholecystitis without cholelithiasis | 1.49 | 0.40 | 0.14 | 0.11 | 0.66 | 8.016E-03 | 2110 | 118942 | 121052 |
| 426.23 | circulatory system | Second degree AV block | 1.73 | 0.55 | 0.19 | 0.15 | 0.91 | 8.099E-03 | 1084 | 120496 | 121580 |

| | | | | | | | | | | | |
|---|---|---|---|---|---|---|---|---|---|---|---|
| 726.2 | musculoskeletal | Synoviopathy | 1.67 | 0.51 | 0.18 | 0.14 | 0.85 | 8.337E-03 | 993 | 119484 | 120477 |
| 840.1 | injuries & poisonings | Muscle/tendon sprain | 2.98 | 1.09 | 0.35 | 0.31 | 1.73 | 8.372E-03 | 161 | 121696 | 121857 |
| 204.1 | neoplasms | Lymphoid leukemia | 1.63 | 0.49 | 0.17 | 0.13 | 0.81 | 8.431E-03 | 1355 | 120961 | 122316 |
| 496 | respiratory | Chronic airway obstruction | 1.14 | 0.13 | 0.05 | 0.03 | 0.22 | 8.487E-03 | 35402 | 77132 | 112534 |
| 601.11 | genitourinary | Acute prostatitis | 1.92 | 0.65 | 0.22 | 0.18 | 1.07 | 8.688E-03 | 666 | 117101 | 117767 |
| 871.4 | injuries & poisonings | Open wound of toe(s) | 1.87 | 0.62 | 0.22 | 0.17 | 1.02 | 8.697E-03 | 696 | 120205 | 120901 |
| 593 | genitourinary | Hematuria | 1.19 | 0.17 | 0.06 | 0.04 | 0.30 | 8.872E-03 | 15202 | 98762 | 113964 |
| 743.11 | musculoskeletal | Osteoporosis NOS | 1.25 | 0.22 | 0.08 | 0.06 | 0.38 | 9.010E-03 | 7567 | 111864 | 119431 |
| 586 | genitourinary | Other disorders of the kidney and ureters | 1.21 | 0.19 | 0.07 | 0.05 | 0.33 | 9.125E-03 | 11790 | 99827 | 111617 |
| 222 | neoplasms | Benign neoplasm of male genital organs | 0.55 | -0.60 | 0.26 | -1.15 | -0.14 | 9.159E-03 | 1737 | 115045 | 116782 |
| 41.2 | infectious diseases | Streptococcus infection | 1.66 | 0.51 | 0.18 | 0.13 | 0.84 | 9.180E-03 | 1144 | 118968 | 120112 |
| 275 | endocrine/metabolic | Disorders of mineral metabolism | 1.24 | 0.22 | 0.08 | 0.05 | 0.38 | 9.295E-03 | 7675 | 107320 | 114995 |
| 216.1 | neoplasms | Screening for malignant neoplasms of the skin | 1.30 | 0.26 | 0.10 | 0.07 | 0.44 | 9.503E-03 | 5643 | 108785 | 114428 |
| 801 | injuries & poisonings | Fracture of ankle and foot | 1.32 | 0.28 | 0.10 | 0.07 | 0.48 | 9.811E-03 | 3785 | 116463 | 120248 |
| 527 | digestive | Diseases of the salivary glands | 1.39 | 0.33 | 0.12 | 0.08 | 0.56 | 9.822E-03 | 2782 | 115869 | 118651 |
| 681.7 | dermatologic | Cellulitis and abscess of trunk | 1.47 | 0.38 | 0.14 | 0.10 | 0.65 | 9.939E-03 | 1983 | 117447 | 119430 |
| 681.6 | dermatologic | Cellulitis and abscess of foot, toe | 1.69 | 0.53 | 0.19 | 0.13 | 0.88 | 1.004E-02 | 1008 | 120040 | 121048 |
| 727.6 | musculoskeletal | Rupture of tendon, nontraumatic | 1.32 | 0.28 | 0.10 | 0.07 | 0.47 | 1.008E-02 | 4370 | 115067 | 119437 |
| 520 | digestive | Disorders of tooth development | 1.76 | 0.57 | 0.20 | 0.14 | 0.95 | 1.026E-02 | 744 | 119085 | 119829 |
| 172.1 | neoplasms | Melanomas of skin, dx or hx | 1.16 | 0.15 | 0.06 | 0.04 | 0.26 | 1.035E-02 | 19897 | 96112 | 116009 |
| 415.21 | circulatory system | Primary pulmonary hypertension | 1.86 | 0.62 | 0.22 | 0.16 | 1.03 | 1.038E-02 | 700 | 120621 | 121321 |
| 250.41 | endocrine/metabolic | Impaired fasting glucose | 1.19 | 0.17 | 0.07 | 0.04 | 0.30 | 1.041E-02 | 12796 | 103276 | 116072 |
| 733 | musculoskeletal | Other disorders of bone and cartilage | 1.37 | 0.31 | 0.12 | 0.08 | 0.54 | 1.046E-02 | 3016 | 113922 | 116938 |
| 117 | infectious diseases | Mycoses | 1.70 | 0.53 | 0.19 | 0.13 | 0.89 | 1.059E-02 | 1007 | 119994 | 121001 |

| | | | | | | | | | | | |
|---|---|---|---|---|---|---|---|---|---|---|---|
| 270.3 | endocrine/metabolic | Disorders of plasma protein metabolism | 1.46 | 0.38 | 0.14 | 0.09 | 0.64 | 1.060E-02 | 2290 | 118814 | 121104 |
| 710.11 | musculoskeletal | Acute osteomyelitis | 1.60 | 0.47 | 0.17 | 0.12 | 0.79 | 1.060E-02 | 1298 | 120495 | 121793 |
| 362.7 | sense organs | Hereditary retinal dystrophies | 1.75 | 0.56 | 0.20 | 0.14 | 0.94 | 1.064E-02 | 912 | 119900 | 120812 |
| 509.1 | respiratory | Respiratory failure | 1.23 | 0.21 | 0.08 | 0.05 | 0.36 | 1.077E-02 | 8568 | 109160 | 117728 |
| 503 | respiratory | Pulmonary congestion and hypostasis | 1.78 | 0.58 | 0.21 | 0.14 | 0.96 | 1.082E-02 | 787 | 119075 | 119862 |
| 743.1 | musculoskeletal | Osteoporosis | 1.24 | 0.22 | 0.08 | 0.05 | 0.38 | 1.103E-02 | 7667 | 111714 | 119381 |
| 804 | injuries & poisonings | Fracture of hand or wrist | 1.41 | 0.35 | 0.13 | 0.08 | 0.59 | 1.127E-02 | 2249 | 118309 | 120558 |
| 960 | injuries & poisonings | Poisoning by antibiotics | 1.77 | 0.57 | 0.21 | 0.14 | 0.96 | 1.136E-02 | 778 | 119531 | 120309 |
| 965 | injuries & poisonings | Poisoning by analgesics, antipyretics, and antirheumatics | 3.04 | 1.11 | 0.36 | 0.28 | 1.79 | 1.147E-02 | 136 | 121615 | 121751 |
| 555.21 | digestive | Ulcerative colitis (chronic) | 2.01 | 0.70 | 0.25 | 0.17 | 1.16 | 1.177E-02 | 448 | 121471 | 121919 |
| 743.2 | musculoskeletal | Pathologic fracture | 1.45 | 0.37 | 0.14 | 0.08 | 0.64 | 1.213E-02 | 2058 | 117977 | 120035 |
| 535.2 | digestive | Atrophic gastritis | 1.83 | 0.61 | 0.22 | 0.14 | 1.01 | 1.220E-02 | 684 | 119109 | 119793 |
| 478 | respiratory | Throat pain | 1.89 | 0.64 | 0.23 | 0.15 | 1.07 | 1.226E-02 | 535 | 119429 | 119964 |
| 401.21 | circulatory system | Hypertensive heart disease | 1.21 | 0.19 | 0.07 | 0.04 | 0.33 | 1.233E-02 | 10302 | 103799 | 114101 |
| 526.42 | digestive | Arthralgia/ankylosis of temporomandibular joint | 3.04 | 1.11 | 0.37 | 0.26 | 1.80 | 1.240E-02 | 132 | 121776 | 121908 |
| 571.8 | digestive | Liver abscess and sequelae of chronic liver disease | 0.51 | -0.68 | 0.30 | -1.33 | -0.13 | 1.242E-02 | 1047 | 120581 | 121628 |
| 427.8 | circulatory system | Sinoatrial node dysfunction (Bradycardia) | 1.38 | 0.32 | 0.12 | 0.07 | 0.55 | 1.242E-02 | 3281 | 116831 | 120112 |
| 363 | sense organs | Chorioretinal inflammations, scars, and other disorders of choroid | 1.34 | 0.29 | 0.11 | 0.07 | 0.51 | 1.253E-02 | 3844 | 114387 | 118231 |
| 253.7 | endocrine/metabolic | Other disorders of neurohypophysis | 2.36 | 0.86 | 0.30 | 0.20 | 1.41 | 1.257E-02 | 270 | 121855 | 122125 |
| 270 | endocrine/metabolic | Disorders of protein plasma/amino-acid transport and metabolism | 1.38 | 0.32 | 0.12 | 0.07 | 0.55 | 1.262E-02 | 3070 | 116854 | 119924 |
| 695.9 | dermatologic | Unspecified erythematous condition | 2.33 | 0.85 | 0.30 | 0.19 | 1.39 | 1.322E-02 | 306 | 120253 | 120559 |
| 277.7 | endocrine/metabolic | Dysmetabolic syndrome X | 1.64 | 0.50 | 0.19 | 0.11 | 0.85 | 1.336E-02 | 1018 | 120621 | 121639 |

| | | | | | | | | | | | |
|---|---|---|---|---|---|---|---|---|---|---|---|
| 610 | genitourinary | Benign mammary dysplasias | 1.83 | 0.61 | 0.23 | 0.13 | 1.05 | 1.362E-02 | 260 | 2331 | 2591 |
| 550.1 | digestive | Inguinal hernia | 1.21 | 0.19 | 0.07 | 0.04 | 0.33 | 1.379E-02 | 10819 | 108126 | 118945 |
| 513.31 | respiratory | Apnea | 2.15 | 0.77 | 0.27 | 0.17 | 1.27 | 1.379E-02 | 386 | 121032 | 121418 |
| 800.2 | injuries & poisonings | Fracture of unspecified part of femur | 1.70 | 0.53 | 0.20 | 0.11 | 0.90 | 1.383E-02 | 927 | 121028 | 121955 |
| 250.12 | endocrine/metabolic | Type 1 diabetes with renal manifestations | 2.42 | 0.88 | 0.31 | 0.20 | 1.46 | 1.405E-02 | 250 | 122004 | 122254 |
| 287.1 | hematopoietic | Spontaneous ecchymoses | 3.20 | 1.16 | 0.39 | 0.26 | 1.88 | 1.413E-02 | 130 | 121250 | 121380 |
| 724 | musculoskeletal | Other and unspecified disorders of back | 1.36 | 0.31 | 0.12 | 0.06 | 0.54 | 1.428E-02 | 2832 | 114972 | 117804 |
| 357 | neurological | Inflammatory and toxic neuropathy | 1.29 | 0.25 | 0.10 | 0.05 | 0.44 | 1.464E-02 | 4969 | 112057 | 117026 |
| 701.2 | dermatologic | Scar conditions and fibrosis of skin | 1.39 | 0.33 | 0.13 | 0.07 | 0.58 | 1.478E-02 | 2637 | 114262 | 116899 |
| 70.9 | infectious diseases | Hepatitis NOS | 1.83 | 0.61 | 0.23 | 0.13 | 1.03 | 1.479E-02 | 565 | 120981 | 121546 |
| 327.4 | neurological | Insomnia | 1.15 | 0.14 | 0.06 | 0.03 | 0.25 | 1.483E-02 | 18596 | 93357 | 111953 |
| 363.3 | sense organs | Chorioretinal scars | 1.36 | 0.31 | 0.12 | 0.06 | 0.53 | 1.486E-02 | 3330 | 115586 | 118916 |
| 536.8 | digestive | Dyspepsia and other specified disorders of function of stomach | 1.40 | 0.34 | 0.13 | 0.07 | 0.59 | 1.519E-02 | 2490 | 115537 | 118027 |
| 426.8 | circulatory system | Other cardiac conduction disorders | 2.13 | 0.76 | 0.28 | 0.16 | 1.27 | 1.533E-02 | 358 | 120961 | 121319 |
| 292.3 | mental disorders | Memory loss | 1.29 | 0.25 | 0.10 | 0.05 | 0.45 | 1.550E-02 | 5107 | 112584 | 117691 |
| 751 | congenital anomalies | Genitourinary congenital anomalies | 1.39 | 0.33 | 0.13 | 0.06 | 0.57 | 1.552E-02 | 2792 | 115467 | 118259 |
| 949 | injuries & poisonings | Allergies, other | 1.36 | 0.31 | 0.12 | 0.06 | 0.54 | 1.562E-02 | 2761 | 114389 | 117150 |
| 528.7 | digestive | Sialolithiasis | 3.15 | 1.15 | 0.39 | 0.24 | 1.87 | 1.570E-02 | 122 | 122217 | 122339 |
| 261.41 | endocrine/metabolic | Rickets or osteomalacia | 0.04 | -3.16 | 1.79 | -9.68 | -0.38 | 1.578E-02 | 127 | 122279 | 122406 |
| 790 | symptoms | Nonspecific findings on examination of blood | 1.93 | 0.66 | 0.25 | 0.13 | 1.11 | 1.582E-02 | 504 | 120027 | 120531 |
| 362.6 | sense organs | Peripheral retinal degenerations | 1.38 | 0.32 | 0.13 | 0.06 | 0.57 | 1.598E-02 | 2820 | 115117 | 117937 |
| 362.31 | sense organs | Separation of retinal layers | 1.79 | 0.58 | 0.22 | 0.12 | 0.99 | 1.599E-02 | 691 | 120854 | 121545 |
| 292.1 | mental disorders | Aphasia/speech disturbance | 1.32 | 0.28 | 0.11 | 0.05 | 0.49 | 1.605E-02 | 3755 | 115032 | 118787 |

| | | | | | | | | | | | |
|---|---|---|---|---|---|---|---|---|---|---|---|
| 427.2 | circulatory system | Atrial fibrillation and flutter | 1.13 | 0.12 | 0.05 | 0.02 | 0.22 | 1.628E-02 | 31921 | 85847 | 117768 |
| 352 | neurological | Disorders of other cranial nerves | 1.43 | 0.36 | 0.14 | 0.07 | 0.63 | 1.631E-02 | 2061 | 118589 | 120650 |
| 520.1 | digestive | Hereditary disturbances in tooth structure | 2.12 | 0.75 | 0.28 | 0.15 | 1.26 | 1.685E-02 | 348 | 120947 | 121295 |
| 214 | neoplasms | Lipoma | 1.30 | 0.27 | 0.11 | 0.05 | 0.47 | 1.690E-02 | 4044 | 112991 | 117035 |
| 530.12 | digestive | Ulcer of esophagus | 1.69 | 0.52 | 0.20 | 0.10 | 0.90 | 1.699E-02 | 935 | 120444 | 121379 |
| 480.1 | respiratory | Bacterial pneumonia | 1.29 | 0.25 | 0.10 | 0.05 | 0.45 | 1.706E-02 | 4699 | 112638 | 117337 |
| 327.7 | neurological | Sleep related movement disorders | 1.25 | 0.22 | 0.09 | 0.04 | 0.39 | 1.770E-02 | 5939 | 112232 | 118171 |
| 710.1 | musculoskeletal | Osteomyelitis | 1.37 | 0.31 | 0.13 | 0.06 | 0.55 | 1.774E-02 | 2805 | 118598 | 121403 |
| 112.3 | infectious diseases | Candidiasis of skin and nails | 2.34 | 0.85 | 0.31 | 0.16 | 1.43 | 1.781E-02 | 245 | 121209 | 121454 |
| 38.3 | infectious diseases | Bacteremia | 1.37 | 0.31 | 0.13 | 0.06 | 0.55 | 1.814E-02 | 2901 | 117819 | 120720 |
| 711.1 | musculoskeletal | Pyogenic arthritis | 1.80 | 0.59 | 0.23 | 0.11 | 1.01 | 1.823E-02 | 623 | 121286 | 121909 |
| 289.9 | hematopoietic | Abnormality of red blood cells | 2.17 | 0.77 | 0.29 | 0.14 | 1.31 | 1.825E-02 | 316 | 121153 | 121469 |
| 726.3 | musculoskeletal | Bursitis | 1.30 | 0.26 | 0.11 | 0.05 | 0.47 | 1.826E-02 | 3974 | 113491 | 117465 |
| 353.2 | neurological | Nerve root lesions | 1.67 | 0.51 | 0.20 | 0.09 | 0.89 | 1.846E-02 | 767 | 120312 | 121079 |
| 300 | mental disorders | Anxiety, phobic and dissociative disorders | 1.12 | 0.11 | 0.05 | 0.02 | 0.21 | 1.848E-02 | 37899 | 75856 | 113755 |
| 737.3 | musculoskeletal | Kyphoscoliosis and scoliosis | 1.56 | 0.44 | 0.18 | 0.08 | 0.77 | 1.869E-02 | 1096 | 119318 | 120414 |
| 783 | symptoms | Fever of unknown origin | 1.29 | 0.26 | 0.11 | 0.04 | 0.46 | 1.876E-02 | 4345 | 110296 | 114641 |
| 411.3 | circulatory system | Angina pectoris | 1.17 | 0.16 | 0.07 | 0.03 | 0.29 | 1.881E-02 | 13829 | 99166 | 112995 |
| 38.2 | infectious diseases | Gram positive septicemia | 1.97 | 0.68 | 0.26 | 0.12 | 1.16 | 1.914E-02 | 444 | 121275 | 121719 |
| 149.4 | neoplasms | Cancer of larynx | 0.45 | -0.80 | 0.39 | -1.69 | -0.12 | 1.933E-02 | 853 | 121428 | 122281 |
| 375 | sense organs | Disorders of lacrimal system | 1.58 | 0.46 | 0.18 | 0.08 | 0.80 | 1.944E-02 | 1281 | 119393 | 120674 |
| 250.15 | endocrine/metabolic | Diabetes type 1 with peripheral circulatory disorders | 2.99 | 1.09 | 0.39 | 0.20 | 1.80 | 1.953E-02 | 137 | 122197 | 122334 |
| 527.2 | digestive | Sialoadenitis | 1.92 | 0.65 | 0.25 | 0.11 | 1.12 | 1.971E-02 | 464 | 121315 | 121779 |
| 270.38 | endocrine/metabolic | Other specified disorders of plasma protein metabolism | 2.20 | 0.79 | 0.30 | 0.13 | 1.33 | 2.004E-02 | 315 | 121299 | 121614 |
| 599.1 | genitourinary | Urinary obstruction | 1.59 | 0.46 | 0.18 | 0.08 | 0.81 | 2.006E-02 | 1261 | 118509 | 119770 |
| 360 | sense organs | Disorders of the globe | 1.95 | 0.67 | 0.26 | 0.11 | 1.14 | 2.015E-02 | 491 | 121109 | 121600 |
| 204.12 | neoplasms | Lymphoid leukemia, chronic | 1.57 | 0.45 | 0.18 | 0.07 | 0.79 | 2.031E-02 | 1244 | 121137 | 122381 |

| | | | | | | | | | | | |
|---|---|---|---|---|---|---|---|---|---|---|---|
| 411.2 | circulatory system | Myocardial infarction | 1.17 | 0.15 | 0.07 | 0.02 | 0.28 | 2.044E-02 | 14749 | 100690 | 115439 |
| 870.1 | injuries & poisonings | Open wound or laceration of eye or eyelid | 2.11 | 0.75 | 0.28 | 0.12 | 1.27 | 2.053E-02 | 381 | 121250 | 121631 |
| 288.2 | hematopoietic | Elevated white blood cell count | 1.29 | 0.25 | 0.11 | 0.04 | 0.45 | 2.056E-02 | 4243 | 111432 | 115675 |
| 524.3 | digestive | Anomalies of tooth position/malocclusion | 1.71 | 0.54 | 0.21 | 0.09 | 0.93 | 2.065E-02 | 738 | 120293 | 121031 |
| 427.21 | circulatory system | Atrial fibrillation | 1.13 | 0.12 | 0.05 | 0.02 | 0.22 | 2.084E-02 | 30922 | 86805 | 117727 |
| 241 | endocrine/metabolic | Nontoxic nodular goiter | 1.26 | 0.23 | 0.10 | 0.04 | 0.42 | 2.104E-02 | 4941 | 115959 | 120900 |
| 989 | injuries & poisonings | Toxic effect of other substances, chiefly nonmedicinal as to source | 2.10 | 0.74 | 0.28 | 0.12 | 1.27 | 2.108E-02 | 369 | 121316 | 121685 |
| 706.2 | dermatologic | Sebaceous cyst | 1.19 | 0.17 | 0.07 | 0.03 | 0.31 | 2.117E-02 | 10645 | 102124 | 112769 |
| 939.1 | dermatologic | Contact and allergic dermatitis of eyelid | 3.27 | 1.19 | 0.42 | 0.19 | 1.96 | 2.145E-02 | 104 | 122071 | 122175 |
| 781 | symptoms | Symptoms involving nervous and musculoskeletal systems | 1.36 | 0.31 | 0.13 | 0.05 | 0.55 | 2.162E-02 | 2794 | 114127 | 116921 |
| 426.9 | circulatory system | Cardiac pacemaker/device in situ | 1.19 | 0.18 | 0.07 | 0.03 | 0.32 | 2.180E-02 | 11225 | 108988 | 120213 |
| 565.1 | digestive | Anal and rectal polyp | 1.61 | 0.47 | 0.19 | 0.07 | 0.83 | 2.196E-02 | 1003 | 117394 | 118397 |
| 213 | neoplasms | Benign neoplasm of bone and articular cartilage | 2.52 | 0.92 | 0.34 | 0.14 | 1.56 | 2.208E-02 | 187 | 121716 | 121903 |
| 601.1 | genitourinary | Prostatitis | 1.33 | 0.28 | 0.12 | 0.04 | 0.51 | 2.247E-02 | 3587 | 111375 | 114962 |
| 807 | injuries & poisonings | Fracture of ribs | 1.39 | 0.33 | 0.14 | 0.05 | 0.59 | 2.252E-02 | 2302 | 116674 | 118976 |
| 721.2 | musculoskeletal | Spondylosis with myelopathy | 1.46 | 0.38 | 0.16 | 0.05 | 0.68 | 2.315E-02 | 1547 | 119295 | 120842 |
| 440.21 | circulatory system | Atherosclerosis of native arteries of the extremities with ulceration or gangrene | 1.62 | 0.48 | 0.20 | 0.07 | 0.85 | 2.411E-02 | 993 | 120702 | 121695 |
| 972 | injuries & poisonings | Poisoning by agents primarily affecting the cardiovascular system | 2.06 | 0.72 | 0.29 | 0.10 | 1.25 | 2.450E-02 | 367 | 120222 | 120589 |
| 297.1 | mental disorders | Suicidal ideation | 1.36 | 0.31 | 0.13 | 0.04 | 0.56 | 2.458E-02 | 2037 | 118828 | 120865 |
| 278.4 | endocrine/metabolic | Abnormal weight gain | 1.74 | 0.56 | 0.23 | 0.07 | 0.98 | 2.512E-02 | 609 | 119650 | 120259 |
| 521.2 | digestive | Dental abrasion, erosion and attrition | 1.38 | 0.32 | 0.14 | 0.04 | 0.58 | 2.525E-02 | 2115 | 117279 | 119394 |

| | | | | | | | | | | | |
|---|---|---|---|---|---|---|---|---|---|---|---|
| 705 | dermatologic | Disorders of sweat glands | 1.67 | 0.51 | 0.21 | 0.07 | 0.91 | 2.533E-02 | 714 | 120236 | 120950 |
| 579.8 | digestive | Nonspecific abnormal findings in stool contents | 1.35 | 0.30 | 0.13 | 0.04 | 0.55 | 2.535E-02 | 2676 | 112842 | 115518 |
| 261.4 | endocrine/metabolic | Vitamin D deficiency | 1.14 | 0.13 | 0.06 | 0.02 | 0.24 | 2.537E-02 | 19781 | 92846 | 112627 |
| 378.5 | sense organs | Paralytic strabismus | 1.60 | 0.47 | 0.19 | 0.06 | 0.83 | 2.539E-02 | 1014 | 120907 | 121921 |
| 132 | infectious diseases | Infestation (lice, mites) | 1.90 | 0.64 | 0.26 | 0.08 | 1.12 | 2.540E-02 | 463 | 120994 | 121457 |
| 803.3 | injuries & poisonings | Fracture of clavicle or scapula | 1.66 | 0.51 | 0.21 | 0.06 | 0.90 | 2.592E-02 | 761 | 121298 | 122059 |
| 874 | injuries & poisonings | Complication of amputation stump | 1.90 | 0.64 | 0.26 | 0.08 | 1.12 | 2.605E-02 | 454 | 121733 | 122187 |
| 710 | musculoskeletal | Osteomyelitis, periostitis, and other infections involving bone | 1.34 | 0.29 | 0.13 | 0.04 | 0.53 | 2.616E-02 | 2884 | 118380 | 121264 |
| 301.1 | mental disorders | Schizoid personality disorder | 2.04 | 0.71 | 0.29 | 0.08 | 1.25 | 2.751E-02 | 305 | 122036 | 122341 |
| 442.8 | circulatory system | Aneurysm of other specified artery | 1.93 | 0.66 | 0.27 | 0.08 | 1.15 | 2.764E-02 | 415 | 121473 | 121888 |
| 495.11 | respiratory | Chronic obstructive asthma with exacerbation | 2.10 | 0.74 | 0.30 | 0.09 | 1.29 | 2.776E-02 | 327 | 121558 | 121885 |
| 458.1 | circulatory system | Orthostatic hypotension | 1.25 | 0.22 | 0.10 | 0.02 | 0.41 | 2.933E-02 | 5788 | 109634 | 115422 |
| 994 | injuries & poisonings | Sepsis and SIRS | 1.24 | 0.21 | 0.09 | 0.02 | 0.39 | 2.954E-02 | 5698 | 111986 | 117684 |
| 571.81 | digestive | Portal hypertension | 0.44 | -0.83 | 0.43 | -1.81 | -0.07 | 2.967E-02 | 553 | 121445 | 121998 |
| 850 | injuries & poisonings | Hemorrhage or hematoma complicating a procedure | 1.49 | 0.40 | 0.17 | 0.04 | 0.72 | 2.981E-02 | 1394 | 118076 | 119470 |
| 557 | digestive | Intestinal malabsorption (non-celiac) | 2.77 | 1.02 | 0.39 | 0.11 | 1.75 | 2.981E-02 | 117 | 122144 | 122261 |
| 440.9 | circulatory system | Atherosclerosis of aorta | 1.63 | 0.49 | 0.21 | 0.05 | 0.88 | 3.029E-02 | 911 | 117637 | 118548 |
| 80 | infectious diseases | Postoperative infection | 1.42 | 0.35 | 0.15 | 0.03 | 0.64 | 3.093E-02 | 1756 | 118290 | 120046 |
| 130 | infectious diseases | Spirochetal infection | 1.78 | 0.57 | 0.24 | 0.06 | 1.02 | 3.098E-02 | 587 | 119881 | 120468 |
| 592.11 | genitourinary | Acute cystitis | 1.72 | 0.54 | 0.23 | 0.05 | 0.97 | 3.098E-02 | 645 | 119692 | 120337 |
| 477 | respiratory | Epistaxis or throat hemorrhage | 1.30 | 0.26 | 0.12 | 0.02 | 0.49 | 3.102E-02 | 3544 | 115283 | 118827 |
| 366.3 | sense organs | Traumatic cataract | 2.39 | 0.87 | 0.35 | 0.08 | 1.51 | 3.170E-02 | 199 | 121970 | 122169 |
| 276.42 | endocrine/metabolic | Alkalosis | 2.05 | 0.72 | 0.30 | 0.07 | 1.26 | 3.184E-02 | 342 | 120898 | 121240 |

| | | | | | | | | | | | |
|---|---|---|---|---|---|---|---|---|---|---|---|
| 275.5 | endocrine/metabolic | Disorders of calcium/phosphorus metabolism | 1.37 | 0.31 | 0.14 | 0.03 | 0.58 | 3.202E-02 | 2249 | 116607 | 118856 |
| 255 | endocrine/metabolic | Disorders of adrenal glands | 1.46 | 0.38 | 0.17 | 0.03 | 0.69 | 3.233E-02 | 1496 | 119636 | 121132 |
| 317.1 | mental disorders | Alcoholism | 0.85 | -0.17 | 0.08 | -0.33 | -0.01 | 3.293E-02 | 10347 | 107089 | 117436 |
| 457.3 | circulatory system | Encounter for long-term (current) use of aspirin | 1.37 | 0.32 | 0.14 | 0.03 | 0.58 | 3.301E-02 | 2447 | 115110 | 117557 |
| 512.2 | respiratory | Painful respiration | 1.38 | 0.33 | 0.15 | 0.03 | 0.60 | 3.305E-02 | 2002 | 113970 | 115972 |
| 724.8 | musculoskeletal | Other symptoms referable to back | 1.56 | 0.44 | 0.20 | 0.04 | 0.81 | 3.330E-02 | 891 | 119107 | 119998 |
| 729.1 | musculoskeletal | Rheumatism, unspecified and fibrositis | 1.87 | 0.63 | 0.27 | 0.05 | 1.13 | 3.347E-02 | 280 | 122097 | 122377 |
| 130.1 | infectious diseases | Lyme disease | 1.97 | 0.68 | 0.29 | 0.05 | 1.20 | 3.416E-02 | 383 | 121539 | 121922 |
| 255.2 | endocrine/metabolic | Adrenal hypofunction | 1.87 | 0.62 | 0.27 | 0.05 | 1.12 | 3.464E-02 | 439 | 121727 | 122166 |
| 362.21 | sense organs | Macular degeneration, dry | 1.15 | 0.14 | 0.07 | 0.01 | 0.27 | 3.486E-02 | 17756 | 99102 | 116858 |
| 275.3 | endocrine/metabolic | Disorders of magnesium metabolism | 1.31 | 0.27 | 0.12 | 0.02 | 0.50 | 3.496E-02 | 3002 | 115818 | 118820 |
| 540 | digestive | Appendiceal conditions | 1.56 | 0.45 | 0.20 | 0.03 | 0.82 | 3.503E-02 | 887 | 121293 | 122180 |
| 705.3 | dermatologic | Hidradenitis | 2.25 | 0.81 | 0.34 | 0.06 | 1.44 | 3.514E-02 | 173 | 122184 | 122357 |
| 569.2 | digestive | Gastrointestinal complications | 1.76 | 0.56 | 0.24 | 0.04 | 1.02 | 3.528E-02 | 533 | 120531 | 121064 |
| 352.1 | neurological | Trigeminal nerve disorders [CN5] | 1.63 | 0.49 | 0.22 | 0.04 | 0.89 | 3.536E-02 | 788 | 120791 | 121579 |
| 540.1 | digestive | Appendicitis | 1.59 | 0.46 | 0.21 | 0.03 | 0.85 | 3.578E-02 | 815 | 121452 | 122267 |
| 276.13 | endocrine/metabolic | Hyperpotassemia | 1.22 | 0.20 | 0.09 | 0.01 | 0.38 | 3.580E-02 | 6518 | 109114 | 115632 |
| 474.2 | respiratory | Chronic tonsillitis and adenoiditis | 2.24 | 0.81 | 0.34 | 0.05 | 1.43 | 3.599E-02 | 205 | 121902 | 122107 |
| 509.3 | respiratory | Pulmonary insufficiency or respiratory failure following trauma and surgery | 1.69 | 0.53 | 0.23 | 0.03 | 0.96 | 3.657E-02 | 639 | 120596 | 121235 |
| 244.4 | endocrine/metabolic | Hypothyroidism NOS | 1.13 | 0.12 | 0.06 | 0.01 | 0.23 | 3.686E-02 | 19575 | 100110 | 119685 |
| 588 | genitourinary | Disorders resulting from impaired renal function | 1.34 | 0.29 | 0.13 | 0.02 | 0.54 | 3.695E-02 | 2819 | 118036 | 120855 |
| 193 | neoplasms | Thyroid cancer | 1.67 | 0.51 | 0.23 | 0.03 | 0.93 | 3.715E-02 | 657 | 121732 | 122389 |
| 722.8 | musculoskeletal | Postlaminectomy syndrome | 1.46 | 0.38 | 0.17 | 0.02 | 0.70 | 3.736E-02 | 1259 | 120440 | 121699 |

| | | | | | | | | | | | |
|---|---|---|---|---|---|---|---|---|---|---|---|
| 290.1 | mental disorders | Dementias | 1.19 | 0.17 | 0.08 | 0.01 | 0.33 | 3.780E-02 | 10113 | 108974 | 119087 |
| 526 | digestive | Diseases of the jaws | 1.36 | 0.31 | 0.14 | 0.02 | 0.58 | 3.820E-02 | 1930 | 116147 | 118077 |
| 306 | mental disorders | Other mental disorder | 1.20 | 0.19 | 0.09 | 0.01 | 0.35 | 3.857E-02 | 6037 | 107094 | 113131 |
| 244 | endocrine/metabolic | Hypothyroidism | 1.12 | 0.12 | 0.06 | 0.01 | 0.23 | 3.890E-02 | 20228 | 99431 | 119659 |
| 531.4 | digestive | Peptic ulcer, site unspecified | 1.30 | 0.26 | 0.12 | 0.01 | 0.49 | 3.928E-02 | 3384 | 116630 | 120014 |
| 586.2 | genitourinary | Cyst of kidney, acquired | 1.27 | 0.24 | 0.11 | 0.01 | 0.45 | 3.934E-02 | 4147 | 112716 | 116863 |
| 202.2 | neoplasms | Non-Hodgkins lymphoma | 1.36 | 0.31 | 0.14 | 0.02 | 0.58 | 3.967E-02 | 2222 | 119609 | 121831 |
| 136 | infectious diseases | Other infectious and parasitic diseases | 1.51 | 0.41 | 0.19 | 0.02 | 0.76 | 4.023E-02 | 988 | 118688 | 119676 |
| 537.1 | digestive | Lesions of stomach and duodenum | 0.27 | -1.32 | 0.80 | -3.49 | -0.04 | 4.026E-02 | 329 | 121707 | 122036 |
| 751.12 | congenital anomalies | Congenital anomalies of male genital organs | 2.43 | 0.89 | 0.37 | 0.04 | 1.57 | 4.040E-02 | 163 | 119089 | 119252 |
| 269 | endocrine/metabolic | Proteinuria | 1.30 | 0.26 | 0.12 | 0.01 | 0.50 | 4.042E-02 | 3325 | 116193 | 119518 |
| 595 | genitourinary | Hydronephrosis | 1.33 | 0.29 | 0.13 | 0.01 | 0.54 | 4.063E-02 | 2594 | 117706 | 120300 |
| 960.2 | injuries & poisonings | Allergy/adverse effect of penicillin | 1.73 | 0.55 | 0.25 | 0.02 | 1.00 | 4.091E-02 | 548 | 120797 | 121345 |
| 539 | digestive | Bariatric surgery | 1.66 | 0.51 | 0.23 | 0.02 | 0.93 | 4.101E-02 | 525 | 121750 | 122275 |
| 362.29 | sense organs | Macular degeneration (senile) of retina NOS | 1.22 | 0.20 | 0.09 | 0.01 | 0.38 | 4.103E-02 | 7392 | 110202 | 117594 |
| 195 | neoplasms | Cancer, suspected or other | 1.24 | 0.22 | 0.10 | 0.01 | 0.42 | 4.105E-02 | 4493 | 112696 | 117189 |
| 876 | injuries & poisonings | Posttraumatic wound infection not elsewhere classified | 2.13 | 0.76 | 0.33 | 0.03 | 1.35 | 4.130E-02 | 269 | 121317 | 121586 |
| 800 | injuries & poisonings | Fracture of lower limb | 1.25 | 0.22 | 0.11 | 0.01 | 0.42 | 4.159E-02 | 4232 | 116277 | 120509 |
| 224.1 | neoplasms | Benign neoplasm of eye, uveal | 1.18 | 0.17 | 0.08 | 0.01 | 0.32 | 4.176E-02 | 8561 | 110449 | 119010 |
| 345.3 | neurological | Convulsions | 1.31 | 0.27 | 0.13 | 0.01 | 0.51 | 4.177E-02 | 2711 | 117832 | 120543 |
| 241.1 | endocrine/metabolic | Nontoxic uninodular goiter | 1.26 | 0.23 | 0.11 | 0.01 | 0.44 | 4.192E-02 | 3904 | 116900 | 120804 |
| 41.9 | infectious diseases | Infection with drug-resistant microorganisms | 1.91 | 0.65 | 0.29 | 0.02 | 1.17 | 4.221E-02 | 393 | 121269 | 121662 |
| 202 | neoplasms | Cancer of other lymphoid, histiocytic tissue | 1.35 | 0.30 | 0.14 | 0.01 | 0.56 | 4.225E-02 | 2331 | 119332 | 121663 |
| 791 | symptoms | Gangrene | 1.71 | 0.54 | 0.24 | 0.02 | 0.98 | 4.270E-02 | 615 | 121396 | 122011 |

| | | | | | | | | | | | |
|---|---|---|---|---|---|---|---|---|---|---|---|
| 384 | sense organs | Other disorders of tympanic membrane | 1.50 | 0.41 | 0.19 | 0.01 | 0.76 | 4.286E-02 | 1136 | 120308 | 121444 |
| 590 | genitourinary | Pyelonephritis | 1.52 | 0.42 | 0.19 | 0.01 | 0.78 | 4.290E-02 | 928 | 120279 | 121207 |
| 263 | endocrine/metabolic | Other nutritional deficiency | 2.14 | 0.76 | 0.33 | 0.02 | 1.37 | 4.302E-02 | 241 | 121229 | 121470 |
| 802 | injuries & poisonings | Fracture of pelvis | 1.72 | 0.54 | 0.24 | 0.02 | 0.99 | 4.323E-02 | 545 | 121679 | 122224 |
| 345 | neurological | Epilepsy, recurrent seizures, convulsions | 1.25 | 0.23 | 0.11 | 0.01 | 0.43 | 4.371E-02 | 3945 | 116124 | 120069 |
| 535.9 | digestive | Gastritis and duodenitis, NOS | 1.33 | 0.28 | 0.14 | 0.01 | 0.54 | 4.381E-02 | 2525 | 113887 | 116412 |
| 840.3 | injuries & poisonings | Joint/ligament sprain | 1.37 | 0.32 | 0.15 | 0.01 | 0.60 | 4.388E-02 | 1614 | 116544 | 118158 |
| 453 | circulatory system | Chronic venous hypertension | 1.52 | 0.42 | 0.19 | 0.01 | 0.78 | 4.394E-02 | 1049 | 120720 | 121769 |
| 427.12 | circulatory system | Paroxysmal ventricular tachycardia | 1.30 | 0.26 | 0.12 | 0.01 | 0.50 | 4.439E-02 | 3179 | 116448 | 119627 |
| 306.9 | mental disorders | Tension headache | 1.87 | 0.63 | 0.28 | 0.02 | 1.15 | 4.449E-02 | 333 | 121387 | 121720 |
| 173 | neoplasms | Neoplasm of uncertain behavior of skin | 1.12 | 0.11 | 0.06 | 0.00 | 0.22 | 4.539E-02 | 24944 | 79994 | 104938 |
| 458.2 | circulatory system | Iatrogenic hypotension | 1.60 | 0.47 | 0.22 | 0.01 | 0.88 | 4.582E-02 | 804 | 118410 | 119214 |
| 519 | respiratory | Other diseases of respiratory system, not elsewhere classified | 1.21 | 0.19 | 0.09 | 0.00 | 0.36 | 4.586E-02 | 6156 | 108026 | 114182 |
| 594 | genitourinary | Urinary calculus | 1.15 | 0.14 | 0.07 | 0.00 | 0.28 | 4.618E-02 | 11919 | 105059 | 116978 |
| 741.4 | musculoskeletal | Joint effusions | 1.34 | 0.29 | 0.14 | 0.00 | 0.56 | 4.628E-02 | 2177 | 114264 | 116441 |
| 327.71 | neurological | Restless legs syndrome | 1.23 | 0.21 | 0.10 | 0.00 | 0.41 | 4.648E-02 | 4507 | 115994 | 120501 |
| 333.3 | neurological | Tics and choreas | 2.75 | 1.01 | 0.42 | 0.01 | 1.80 | 4.655E-02 | 108 | 122384 | 122492 |
| 313 | mental disorders | Pervasive developmental disorders | 1.43 | 0.36 | 0.17 | 0.00 | 0.69 | 4.706E-02 | 1104 | 120834 | 121938 |
| 964 | injuries & poisonings | Poisoning by agents primarily affecting blood constituents | 1.82 | 0.60 | 0.27 | 0.01 | 1.10 | 4.773E-02 | 468 | 120682 | 121150 |
| 411.41 | circulatory system | Aneurysm and dissection of heart | 2.46 | 0.90 | 0.39 | 0.00 | 1.61 | 4.800E-02 | 164 | 121993 | 122157 |
| 751.2 | congenital anomalies | Congenital anomalies of urinary system | 1.34 | 0.29 | 0.14 | 0.00 | 0.55 | 4.810E-02 | 2421 | 116845 | 119266 |
| 531.2 | digestive | Gastric ulcer | 1.47 | 0.39 | 0.19 | 0.00 | 0.74 | 4.906E-02 | 1156 | 119625 | 120781 |
| 531.3 | digestive | Duodenal ulcer | 1.59 | 0.46 | 0.22 | 0.00 | 0.87 | 4.994E-02 | 794 | 120533 | 121327 |
| 540.11 | digestive | Acute appendicitis | 1.60 | 0.47 | 0.22 | 0.00 | 0.88 | 5.093E-02 | 688 | 121637 | 122325 |

| | | | | | | | | | | | |
|---|---|---|---|---|---|---|---|---|---|---|---|
| 153.2 | neoplasms | Colon cancer | 0.76 | -0.27 | 0.15 | -0.57 | 0.00 | 5.177E-02 | 3817 | 117221 | 121038 |
| 327.31 | neurological | Central/nonobstroctive sleep apnea | 1.38 | 0.32 | 0.16 | 0.00 | 0.62 | 5.200E-02 | 1643 | 119018 | 120661 |
| 509.2 | respiratory | Respiratory insufficiency | 1.65 | 0.50 | 0.24 | -0.01 | 0.94 | 5.214E-02 | 607 | 120543 | 121150 |
| 433.1 | circulatory system | Occlusion and stenosis of precerebral arteries | 1.16 | 0.15 | 0.07 | 0.00 | 0.29 | 5.265E-02 | 11679 | 102316 | 113995 |
| 577 | digestive | Diseases of pancreas | 1.25 | 0.23 | 0.11 | 0.00 | 0.44 | 5.279E-02 | 3802 | 116555 | 120357 |
| 964.1 | injuries & poisonings | Anticoagulants causing adverse effects | 1.84 | 0.61 | 0.28 | -0.01 | 1.13 | 5.358E-02 | 429 | 120834 | 121263 |
| 594.1 | genitourinary | Calculus of kidney | 1.16 | 0.15 | 0.07 | 0.00 | 0.29 | 5.377E-02 | 9859 | 107680 | 117539 |
| 710.19 | musculoskeletal | Unspecified osteomyelitis | 1.36 | 0.30 | 0.15 | -0.01 | 0.59 | 5.413E-02 | 1948 | 119621 | 121569 |
| 224 | neoplasms | Benign neoplasm of eye | 1.17 | 0.16 | 0.08 | 0.00 | 0.31 | 5.440E-02 | 8956 | 109612 | 118568 |
| 742.2 | musculoskeletal | Pathological, developmental or recurrent dislocation | 2.17 | 0.77 | 0.35 | -0.02 | 1.42 | 5.463E-02 | 191 | 122004 | 122195 |
| 214.1 | neoplasms | Lipoma of skin and subcutaneous tissue | 1.35 | 0.30 | 0.15 | -0.01 | 0.58 | 5.509E-02 | 1927 | 117640 | 119567 |
| 711 | musculoskeletal | Arthropathy associated with infections | 1.53 | 0.42 | 0.21 | -0.01 | 0.81 | 5.612E-02 | 865 | 120597 | 121462 |
| 509.8 | respiratory | Dependence on respirator [Ventilator] or supplemental oxygen | 1.26 | 0.23 | 0.12 | -0.01 | 0.45 | 5.699E-02 | 3827 | 115387 | 119214 |
| 575.7 | digestive | Other disorders of gallbladder | 1.51 | 0.41 | 0.20 | -0.01 | 0.79 | 5.704E-02 | 918 | 119925 | 120843 |
| 753.2 | congenital anomalies | Congenital anomalies of posterior segment of eye | 2.22 | 0.80 | 0.36 | -0.03 | 1.46 | 5.730E-02 | 201 | 121855 | 122056 |
| 800.1 | injuries & poisonings | Fracture of neck of femur | 1.39 | 0.33 | 0.17 | -0.01 | 0.64 | 5.909E-02 | 1622 | 120244 | 121866 |
| 253 | endocrine/metabolic | Disorders of the pituitary gland and its hypothalamic control | 1.39 | 0.33 | 0.17 | -0.01 | 0.64 | 5.909E-02 | 1479 | 119691 | 121170 |
| 420.21 | circulatory system | Acute pericarditis | 2.10 | 0.74 | 0.35 | -0.04 | 1.37 | 5.938E-02 | 220 | 122046 | 122266 |
| 619 | genitourinary | Noninflammatory female genital disorders | 1.55 | 0.44 | 0.23 | -0.02 | 0.87 | 5.965E-02 | 317 | 2262 | 2579 |
| 260.1 | endocrine/metabolic | Cachexia | 1.99 | 0.69 | 0.33 | -0.04 | 1.29 | 6.056E-02 | 277 | 121590 | 121867 |
| 429.3 | circulatory system | Symptoms involving cardiovascular system | 1.33 | 0.29 | 0.15 | -0.01 | 0.57 | 6.079E-02 | 2277 | 114476 | 116753 |

| | | | | | | | | | | | |
|---|---|---|---|---|---|---|---|---|---|---|---|
| 961.1 | injuries & poisonings | Poisoning/allergy of sulfonamides | 2.55 | 0.93 | 0.42 | -0.06 | 1.71 | 6.119E-02 | 119 | 122030 | 122149 |
| 338.1 | neurological | Acute pain | 1.19 | 0.18 | 0.09 | -0.01 | 0.35 | 6.196E-02 | 5773 | 106056 | 111829 |
| 597.2 | genitourinary | Urinary complications NEC | 0.29 | -1.22 | 0.80 | -3.39 | 0.05 | 6.218E-02 | 306 | 120801 | 121107 |
| 474 | respiratory | Acute and chronic tonsillitis | 1.84 | 0.61 | 0.30 | -0.04 | 1.16 | 6.220E-02 | 319 | 121436 | 121755 |
| 519.8 | respiratory | Other diseases of respiratory system, NEC | 1.37 | 0.31 | 0.16 | -0.02 | 0.61 | 6.281E-02 | 1699 | 116873 | 118572 |
| 255.21 | endocrine/metabolic | Glucocorticoid deficiency | 1.80 | 0.59 | 0.29 | -0.04 | 1.11 | 6.326E-02 | 388 | 121835 | 122223 |
| 912 | injuries & poisonings | Insect bite | 1.48 | 0.39 | 0.20 | -0.02 | 0.76 | 6.331E-02 | 1044 | 116914 | 117958 |
| 226 | neoplasms | Benign neoplasm of thyroid glands | 2.07 | 0.73 | 0.35 | -0.05 | 1.36 | 6.340E-02 | 226 | 121905 | 122131 |
| 706.1 | dermatologic | Acne | 1.50 | 0.40 | 0.21 | -0.02 | 0.79 | 6.344E-02 | 819 | 119603 | 120422 |
| 555 | digestive | Inflammatory bowel disease and other gastroenteritis and colitis | 1.33 | 0.28 | 0.15 | -0.02 | 0.56 | 6.397E-02 | 2052 | 119223 | 121275 |
| 592 | genitourinary | Cystitis and urethritis | 1.33 | 0.28 | 0.15 | -0.02 | 0.56 | 6.402E-02 | 2040 | 116180 | 118220 |
| 394.7 | circulatory system | Disease of tricuspid valve | 1.62 | 0.49 | 0.24 | -0.03 | 0.93 | 6.439E-02 | 674 | 119223 | 119897 |
| 189.12 | neoplasms | Malignant neoplasm of renal pelvis | 0.43 | -0.86 | 0.53 | -2.13 | 0.05 | 6.439E-02 | 486 | 121654 | 122140 |
| 362.4 | sense organs | Retinal vascular changes and abnomalities | 1.20 | 0.18 | 0.10 | -0.01 | 0.37 | 6.502E-02 | 6197 | 112204 | 118401 |
| 227 | neoplasms | Benign neoplasm of other endocrine glands and related structures | 1.38 | 0.32 | 0.17 | -0.02 | 0.63 | 6.521E-02 | 1512 | 120022 | 121534 |
| 480.2 | respiratory | Viral pneumonia | 0.38 | -0.98 | 0.62 | -2.54 | 0.06 | 6.840E-02 | 365 | 121321 | 121686 |
| 610.1 | genitourinary | Cystic mastopathy | 1.75 | 0.56 | 0.29 | -0.05 | 1.11 | 6.854E-02 | 146 | 2523 | 2669 |
| 751.1 | congenital anomalies | Congenital anomalies of genital organs | 1.90 | 0.64 | 0.32 | -0.06 | 1.22 | 6.968E-02 | 292 | 121205 | 121497 |
| 145.1 | neoplasms | Cancer of lip | 2.44 | 0.89 | 0.42 | -0.10 | 1.65 | 7.044E-02 | 141 | 122203 | 122344 |
| 276.14 | endocrine/metabolic | Hypopotassemia | 1.18 | 0.16 | 0.09 | -0.01 | 0.33 | 7.066E-02 | 6779 | 108215 | 114994 |
| 375.2 | sense organs | Epiphora | 1.48 | 0.40 | 0.21 | -0.04 | 0.78 | 7.069E-02 | 1054 | 120006 | 121060 |
| 446 | circulatory system | Polyarteritis nodosa and allied conditions | 1.55 | 0.44 | 0.22 | -0.04 | 0.85 | 7.077E-02 | 775 | 121142 | 121917 |
| 430.3 | circulatory system | Subdural hemorrhage | 1.58 | 0.46 | 0.24 | -0.04 | 0.89 | 7.112E-02 | 732 | 121388 | 122120 |

| | | | | | | | | | | | |
|---|---|---|---|---|---|---|---|---|---|---|---|
| 579.2 | digestive | Splenomegaly | 1.50 | 0.41 | 0.21 | -0.04 | 0.80 | 7.207E-02 | 841 | 120532 | 121373 |
| 344 | neurological | Other paralytic syndromes | 1.40 | 0.33 | 0.18 | -0.03 | 0.66 | 7.211E-02 | 1340 | 119791 | 121131 |
| 686 | dermatologic | Other local infections of skin and subcutaneous tissue | 1.39 | 0.33 | 0.18 | -0.03 | 0.66 | 7.245E-02 | 1386 | 116219 | 117605 |
| 771 | symptoms | Musculoskeletal symptoms referable to limbs | 1.76 | 0.57 | 0.29 | -0.06 | 1.09 | 7.258E-02 | 393 | 120462 | 120855 |
| 972.6 | injuries & poisonings | Antihypertensive agents causing adverse effects | 2.24 | 0.81 | 0.39 | -0.09 | 1.52 | 7.317E-02 | 168 | 121117 | 121285 |
| 426.22 | circulatory system | Mobitz II AV block | 2.22 | 0.80 | 0.39 | -0.10 | 1.50 | 7.456E-02 | 197 | 122125 | 122322 |
| 722.7 | musculoskeletal | Intervertebral disc disorder with myelopathy | 1.50 | 0.40 | 0.21 | -0.04 | 0.80 | 7.467E-02 | 849 | 120481 | 121330 |
| 396 | circulatory system | Abnormal heart sounds | 1.20 | 0.18 | 0.10 | -0.02 | 0.38 | 7.484E-02 | 5288 | 110016 | 115304 |
| 612 | genitourinary | Breast conditions, congenital or relating to hormones | 1.40 | 0.33 | 0.18 | -0.04 | 0.67 | 7.732E-02 | 1323 | 119676 | 120999 |
| 592.12 | genitourinary | Chronic cystitis | 1.84 | 0.61 | 0.31 | -0.08 | 1.18 | 7.757E-02 | 325 | 121343 | 121668 |
| 687.3 | dermatologic | Changes in skin texture | 2.37 | 0.86 | 0.42 | -0.12 | 1.62 | 7.920E-02 | 144 | 121288 | 121432 |
| 53.1 | infectious diseases | Herpes zoster with nervous system complications | 1.46 | 0.38 | 0.21 | -0.05 | 0.76 | 8.081E-02 | 1012 | 120680 | 121692 |
| 227.1 | neoplasms | Benign neoplasm of adrenal gland | 1.49 | 0.40 | 0.22 | -0.05 | 0.80 | 8.121E-02 | 822 | 120995 | 121817 |
| 723.1 | musculoskeletal | Torticollis | 2.16 | 0.77 | 0.39 | -0.12 | 1.48 | 8.187E-02 | 150 | 121845 | 121995 |
| 246.7 | endocrine/metabolic | Abnormal results of function study of thyroid | 1.40 | 0.34 | 0.18 | -0.05 | 0.68 | 8.235E-02 | 1164 | 119089 | 120253 |
| 202.24 | neoplasms | Large cell lymphoma | 2.59 | 0.95 | 0.46 | -0.16 | 1.79 | 8.242E-02 | 103 | 122410 | 122513 |
| 596.1 | genitourinary | Bladder neck obstruction | 1.29 | 0.25 | 0.14 | -0.03 | 0.52 | 8.350E-02 | 2732 | 116723 | 119455 |
| 389.2 | sense organs | Conductive hearing loss | 1.31 | 0.27 | 0.15 | -0.04 | 0.56 | 8.445E-02 | 1996 | 117371 | 119367 |
| 569 | digestive | Other disorders of intestine | 1.31 | 0.27 | 0.15 | -0.04 | 0.56 | 8.458E-02 | 1995 | 115421 | 117416 |
| 272.9 | endocrine/metabolic | Unspecified disorder of lipoid metabolism | 1.57 | 0.45 | 0.24 | -0.07 | 0.89 | 8.493E-02 | 696 | 119435 | 120131 |
| 578.8 | digestive | Hemorrhage of rectum and anus | 1.22 | 0.20 | 0.11 | -0.03 | 0.42 | 8.501E-02 | 3898 | 112566 | 116464 |
| 285.21 | hematopoietic | Anemia in chronic kidney disease | 1.24 | 0.22 | 0.12 | -0.03 | 0.45 | 8.505E-02 | 3759 | 116195 | 119954 |
| 190 | neoplasms | Cancer of eye | 1.86 | 0.62 | 0.32 | -0.10 | 1.21 | 8.564E-02 | 331 | 121763 | 122094 |

| | | | | | | | | | | | |
|---|---|---|---|---|---|---|---|---|---|---|---|
| 681.2 | dermatologic | Cellulitis and abscess of face/neck | 1.49 | 0.40 | 0.22 | -0.06 | 0.80 | 8.576E-02 | 785 | 119954 | 120739 |
| 443.1 | circulatory system | Raynaud's syndrome | 1.59 | 0.47 | 0.25 | -0.07 | 0.93 | 8.608E-02 | 533 | 121621 | 122154 |
| 528.12 | digestive | Oral aphthae | 2.16 | 0.77 | 0.39 | -0.13 | 1.49 | 8.661E-02 | 156 | 121646 | 121802 |
| 735.3 | musculoskeletal | Hallux valgus (Bunion) | 1.19 | 0.18 | 0.10 | -0.03 | 0.37 | 8.680E-02 | 4857 | 113351 | 118208 |
| 149 | neoplasms | Cancer of larynx, pharynx, nasal cavities | 0.72 | -0.33 | 0.21 | -0.76 | 0.05 | 8.768E-02 | 1874 | 119947 | 121821 |
| 735.22 | musculoskeletal | Claw toe (acquired) | 2.13 | 0.76 | 0.39 | -0.14 | 1.46 | 8.892E-02 | 180 | 122121 | 122301 |
| 938.1 | injuries & poisonings | Acute dermatitis due to solar radiation | 2.01 | 0.70 | 0.36 | -0.13 | 1.36 | 8.948E-02 | 219 | 121267 | 121486 |
| 279.7 | endocrine/metabolic | Other immunological findings | 1.72 | 0.54 | 0.29 | -0.09 | 1.08 | 8.963E-02 | 352 | 121471 | 121823 |
| 575.8 | digestive | Other disorders of biliary tract | 1.61 | 0.47 | 0.26 | -0.08 | 0.95 | 8.991E-02 | 540 | 120924 | 121464 |
| 578.9 | digestive | Hemorrhage of gastrointestinal tract | 1.20 | 0.18 | 0.10 | -0.03 | 0.38 | 9.248E-02 | 5163 | 112494 | 117657 |
| 378.2 | sense organs | Nystagmus and other irregular eye movements | 1.94 | 0.66 | 0.35 | -0.13 | 1.30 | 9.281E-02 | 224 | 121949 | 122173 |
| 872 | injuries & poisonings | Traumatic amputation | 1.40 | 0.34 | 0.19 | -0.06 | 0.70 | 9.298E-02 | 1157 | 120699 | 121856 |
| 350.1 | neurological | Abnormal involuntary movements | 1.17 | 0.16 | 0.09 | -0.03 | 0.33 | 9.350E-02 | 6310 | 110205 | 116515 |
| 593.2 | genitourinary | Microscopic hematuria | 1.17 | 0.15 | 0.09 | -0.03 | 0.33 | 9.352E-02 | 6529 | 110728 | 117257 |
| 288.3 | hematopoietic | Eosinophilia | 1.78 | 0.58 | 0.31 | -0.11 | 1.14 | 9.377E-02 | 338 | 121749 | 122087 |
| 496.2 | respiratory | Chronic bronchitis | 1.14 | 0.13 | 0.08 | -0.02 | 0.28 | 9.414E-02 | 9723 | 106136 | 115859 |
| 751.21 | congenital anomalies | Cystic kidney disease | 1.30 | 0.26 | 0.15 | -0.05 | 0.54 | 9.460E-02 | 2217 | 117359 | 119576 |
| 327.6 | neurological | Circadian rhythm sleep disorder | 2.02 | 0.70 | 0.37 | -0.15 | 1.39 | 9.642E-02 | 182 | 121836 | 122018 |
| 618 | genitourinary | Genital prolapse | 1.52 | 0.42 | 0.24 | -0.08 | 0.87 | 9.706E-02 | 292 | 2461 | 2753 |
| 345.1 | neurological | Epilepsy | 1.28 | 0.25 | 0.14 | -0.05 | 0.52 | 9.711E-02 | 2118 | 119169 | 121287 |
| 588.1 | genitourinary | Renal osteodystrophy | 1.55 | 0.44 | 0.25 | -0.09 | 0.90 | 9.918E-02 | 666 | 121294 | 121960 |
| 612.2 | genitourinary | Hypertrophy of breast (Gynecomastia) | 1.37 | 0.32 | 0.18 | -0.06 | 0.66 | 9.955E-02 | 1315 | 119709 | 121024 |
| 529 | digestive | Diseases and other conditions of the tongue | 1.58 | 0.45 | 0.26 | -0.10 | 0.93 | 1.024E-01 | 539 | 120737 | 121276 |
| 358 | neurological | Myoneural disorders | 1.63 | 0.49 | 0.27 | -0.11 | 0.99 | 1.027E-01 | 518 | 121863 | 122381 |

| | | | | | | | | | | | |
|---|---|---|---|---|---|---|---|---|---|---|---|
| 380 | sense organs | Disorders of external ear | 1.41 | 0.35 | 0.20 | -0.08 | 0.72 | 1.032E-01 | 1090 | 118454 | 119544 |
| 426.92 | circulatory system | Cardiac defibrillator in situ | 1.22 | 0.20 | 0.12 | -0.04 | 0.42 | 1.032E-01 | 4035 | 117242 | 121277 |
| 280.2 | hematopoietic | Iron deficiency anemia secondary to blood loss (chronic) | 1.31 | 0.27 | 0.16 | -0.06 | 0.57 | 1.033E-01 | 1906 | 117629 | 119535 |
| 771.2 | symptoms | Cramp of limb | 1.33 | 0.29 | 0.17 | -0.06 | 0.61 | 1.035E-01 | 1615 | 117497 | 119112 |
| 537 | digestive | Other disorders of stomach and duodenum | 1.30 | 0.26 | 0.16 | -0.06 | 0.56 | 1.040E-01 | 1909 | 116264 | 118173 |
| 580.14 | genitourinary | Chronic glomerulonephritis, NOS | 2.41 | 0.88 | 0.46 | -0.23 | 1.71 | 1.043E-01 | 120 | 122280 | 122400 |
| 281.1 | hematopoietic | Megaloblastic anemia | 1.19 | 0.17 | 0.10 | -0.04 | 0.37 | 1.055E-01 | 4960 | 113948 | 118908 |
| 187.8 | neoplasms | Neoplasm of uncertain behavior of male genital organs | 1.94 | 0.66 | 0.36 | -0.16 | 1.32 | 1.055E-01 | 251 | 118850 | 119101 |
| 496.21 | respiratory | Obstructive chronic bronchitis | 1.14 | 0.13 | 0.08 | -0.03 | 0.29 | 1.058E-01 | 8635 | 108272 | 116907 |
| 289.4 | hematopoietic | Lymphadenitis | 1.22 | 0.20 | 0.12 | -0.04 | 0.43 | 1.066E-01 | 3305 | 114195 | 117500 |
| 153 | neoplasms | Colorectal cancer | 0.81 | -0.21 | 0.13 | -0.49 | 0.04 | 1.068E-01 | 4216 | 116726 | 120942 |
| 601.3 | genitourinary | Orchitis and epididymitis | 1.32 | 0.28 | 0.17 | -0.06 | 0.59 | 1.070E-01 | 1706 | 115828 | 117534 |
| 389.4 | sense organs | Tinnitus | 1.09 | 0.09 | 0.05 | -0.02 | 0.19 | 1.072E-01 | 23394 | 81510 | 104904 |
| 701.3 | dermatologic | Circumscribed scleroderma | 1.81 | 0.59 | 0.33 | -0.15 | 1.21 | 1.073E-01 | 204 | 122178 | 122382 |
| 370.3 | sense organs | Keratoconjunctivitis | 1.29 | 0.26 | 0.15 | -0.06 | 0.54 | 1.078E-01 | 1950 | 116671 | 118621 |
| 427.41 | circulatory system | Ventricular fibrillation and flutter | 1.65 | 0.50 | 0.29 | -0.12 | 1.03 | 1.080E-01 | 429 | 121483 | 121912 |
| 347 | neurological | Cataplexy and narcolepsy | 1.94 | 0.66 | 0.37 | -0.17 | 1.33 | 1.088E-01 | 206 | 122239 | 122445 |
| 346.1 | neurological | Nonspecific abnormal findings on radiological and other examination of skull and head | 1.73 | 0.55 | 0.31 | -0.14 | 1.12 | 1.099E-01 | 340 | 120247 | 120587 |
| 277.4 | endocrine/metabolic | Disorders of bilirubin excretion | 1.48 | 0.39 | 0.23 | -0.10 | 0.82 | 1.102E-01 | 750 | 120518 | 121268 |
| 426.24 | circulatory system | Atrioventricular block, complete | 1.24 | 0.21 | 0.13 | -0.05 | 0.46 | 1.107E-01 | 3244 | 118381 | 121625 |
| 580.31 | genitourinary | Nephritis and nephropathy in diseases classified elsewhere | 1.48 | 0.39 | 0.23 | -0.10 | 0.81 | 1.108E-01 | 832 | 120742 | 121574 |
| 426.25 | circulatory system | Other heart block | 1.46 | 0.38 | 0.22 | -0.09 | 0.79 | 1.113E-01 | 918 | 120480 | 121398 |

| | | | | | | | | | | | |
|---|---|---|---|---|---|---|---|---|---|---|---|
| 724.9 | musculoskeletal | Other unspecified back disorders | 1.38 | 0.32 | 0.19 | -0.08 | 0.68 | 1.114E-01 | 1082 | 119886 | 120968 |
| 457.2 | circulatory system | Encounter for long-term (current) use of antiplatelets/antithrombotics | 1.46 | 0.38 | 0.22 | -0.10 | 0.79 | 1.119E-01 | 866 | 120068 | 120934 |
| 836 | injuries & poisonings | Traumatic arthropathy | 1.30 | 0.26 | 0.16 | -0.06 | 0.55 | 1.120E-01 | 1686 | 118544 | 120230 |
| 429.2 | circulatory system | Abnormal function study of cardiovascular system | 1.30 | 0.26 | 0.16 | -0.06 | 0.56 | 1.122E-01 | 1840 | 115748 | 117588 |
| 805 | injuries & poisonings | Fracture of vertebral column without mention of spinal cord injury | 1.25 | 0.22 | 0.14 | -0.06 | 0.48 | 1.135E-01 | 2608 | 117984 | 120592 |
| 350.3 | neurological | Lack of coordination | 1.30 | 0.26 | 0.16 | -0.07 | 0.56 | 1.138E-01 | 1861 | 117919 | 119780 |
| 560.2 | digestive | Impaction of intestine | 1.63 | 0.49 | 0.29 | -0.13 | 1.01 | 1.147E-01 | 455 | 120948 | 121403 |
| 737.1 | musculoskeletal | Kyphosis (acquired) | 1.63 | 0.49 | 0.29 | -0.13 | 1.01 | 1.163E-01 | 430 | 121294 | 121724 |
| 555.1 | digestive | Regional enteritis | 1.45 | 0.37 | 0.22 | -0.10 | 0.78 | 1.166E-01 | 771 | 121427 | 122198 |
| 550.4 | digestive | Umbilical hernia | 1.20 | 0.18 | 0.11 | -0.05 | 0.39 | 1.183E-01 | 4049 | 116253 | 120302 |
| 994.2 | injuries & poisonings | Sepsis | 1.17 | 0.16 | 0.10 | -0.04 | 0.35 | 1.191E-01 | 5283 | 112588 | 117871 |
| 723 | musculoskeletal | Other disorders of cervical region | 1.48 | 0.39 | 0.24 | -0.11 | 0.83 | 1.196E-01 | 646 | 120574 | 121220 |
| 345.12 | neurological | Partial epilepsy | 1.42 | 0.35 | 0.21 | -0.10 | 0.74 | 1.204E-01 | 881 | 121053 | 121934 |
| 395.6 | circulatory system | Heart valve replaced | 1.20 | 0.19 | 0.12 | -0.05 | 0.41 | 1.206E-01 | 4140 | 117314 | 121454 |
| 285.22 | hematopoietic | Anemia in neoplastic disease | 1.48 | 0.39 | 0.24 | -0.11 | 0.82 | 1.212E-01 | 742 | 120786 | 121528 |
| 627.3 | genitourinary | Postmenopausal atrophic vaginitis | 1.43 | 0.36 | 0.23 | -0.10 | 0.79 | 1.217E-01 | 319 | 2260 | 2579 |
| 317 | mental disorders | Alcohol-related disorders | 0.90 | -0.10 | 0.07 | -0.24 | 0.03 | 1.219E-01 | 14942 | 101828 | 116770 |
| 275.6 | endocrine/metabolic | Hypercalcemia | 1.28 | 0.24 | 0.15 | -0.07 | 0.53 | 1.238E-01 | 1896 | 119007 | 120903 |
| 446.9 | circulatory system | Arteritis NOS | 1.87 | 0.63 | 0.36 | -0.20 | 1.29 | 1.238E-01 | 229 | 122016 | 122245 |
| 242 | endocrine/metabolic | Thyrotoxicosis with or without goiter | 1.30 | 0.26 | 0.16 | -0.08 | 0.57 | 1.254E-01 | 1545 | 119611 | 121156 |
| 601.12 | genitourinary | Chronic prostatitis | 1.33 | 0.29 | 0.18 | -0.09 | 0.62 | 1.259E-01 | 1547 | 116010 | 117557 |
| 705.1 | dermatologic | Dyshidrosis | 1.64 | 0.49 | 0.30 | -0.16 | 1.04 | 1.268E-01 | 400 | 121393 | 121793 |
| 592.1 | genitourinary | Cystitis | 1.28 | 0.25 | 0.16 | -0.07 | 0.54 | 1.277E-01 | 1850 | 116807 | 118657 |
| 525.2 | digestive | Atrophy of edentulous alveolar ridge | 1.36 | 0.31 | 0.19 | -0.09 | 0.67 | 1.283E-01 | 1064 | 119960 | 121024 |

| | | | | | | | | | | | |
|---|---|---|---|---|---|---|---|---|---|---|---|
| 274.1 | endocrine/metabolic | Gout | 0.90 | -0.10 | 0.07 | -0.24 | 0.03 | 1.293E-01 | 15776 | 102925 | 118701 |
| 260.6 | endocrine/metabolic | Anorexia | 1.46 | 0.38 | 0.24 | -0.12 | 0.81 | 1.298E-01 | 788 | 119593 | 120381 |
| 284 | hematopoietic | Aplastic anemia | 1.28 | 0.25 | 0.16 | -0.08 | 0.54 | 1.300E-01 | 1951 | 119143 | 121094 |
| 427.4 | circulatory system | Cardiac arrest and ventricular fibrillation | 1.40 | 0.34 | 0.21 | -0.11 | 0.73 | 1.311E-01 | 932 | 120172 | 121104 |
| 447.7 | circulatory system | Aortic ectasia | 1.57 | 0.45 | 0.27 | -0.15 | 0.95 | 1.312E-01 | 504 | 120600 | 121104 |
| 531.1 | digestive | Hemorrhage from gastrointestinal ulcer | 1.44 | 0.37 | 0.23 | -0.12 | 0.79 | 1.320E-01 | 815 | 120632 | 121447 |
| 333.1 | neurological | Essential tremor | 1.17 | 0.16 | 0.10 | -0.05 | 0.35 | 1.333E-01 | 4921 | 115166 | 120087 |
| 443.8 | circulatory system | Other specified peripheral vascular diseases | 1.39 | 0.33 | 0.21 | -0.11 | 0.72 | 1.341E-01 | 1055 | 119667 | 120722 |
| 285.1 | hematopoietic | Acute posthemorrhagic anemia | 1.25 | 0.22 | 0.14 | -0.07 | 0.50 | 1.346E-01 | 2268 | 115601 | 117869 |
| 740.2 | musculoskeletal | Osteoarthrosis, generalized | 1.15 | 0.14 | 0.09 | -0.04 | 0.32 | 1.347E-01 | 6326 | 110292 | 116618 |
| 300.9 | mental disorders | Posttraumatic stress disorder | 0.92 | -0.09 | 0.06 | -0.20 | 0.03 | 1.351E-01 | 20044 | 99163 | 119207 |
| 963.1 | injuries & poisonings | Antineoplastic and immunosuppressive drugs causing adverse effects | 1.45 | 0.37 | 0.24 | -0.13 | 0.81 | 1.351E-01 | 646 | 120704 | 121350 |
| 818.1 | injuries & poisonings | Subdural hemorrhage (injury) | 1.92 | 0.65 | 0.39 | -0.24 | 1.35 | 1.358E-01 | 211 | 122176 | 122387 |
| 527.7 | digestive | Disturbance of salivary secretion | 1.26 | 0.23 | 0.15 | -0.08 | 0.52 | 1.366E-01 | 1942 | 117682 | 119624 |
| 242.3 | endocrine/metabolic | Exophthalmos | 1.83 | 0.60 | 0.36 | -0.22 | 1.26 | 1.366E-01 | 244 | 121975 | 122219 |
| 574.2 | digestive | Calculus of bile duct | 1.35 | 0.30 | 0.19 | -0.10 | 0.65 | 1.369E-01 | 1273 | 120223 | 121496 |
| 496.3 | respiratory | Bronchiectasis | 1.45 | 0.37 | 0.24 | -0.13 | 0.81 | 1.383E-01 | 777 | 120874 | 121651 |
| 384.4 | sense organs | Perforation of tympanic membrane | 1.39 | 0.33 | 0.21 | -0.11 | 0.72 | 1.386E-01 | 986 | 120754 | 121740 |
| 495.2 | respiratory | Asthma with exacerbation | 1.37 | 0.31 | 0.20 | -0.11 | 0.69 | 1.395E-01 | 906 | 120485 | 121391 |
| 401.3 | circulatory system | Other hypertensive complications | 1.28 | 0.25 | 0.16 | -0.09 | 0.56 | 1.418E-01 | 1719 | 117917 | 119636 |
| 769 | symptoms | Nonallopathic lesions NEC | 1.21 | 0.19 | 0.13 | -0.07 | 0.43 | 1.423E-01 | 2703 | 119006 | 121709 |
| 575.2 | digestive | Obstruction of bile duct | 1.64 | 0.50 | 0.31 | -0.19 | 1.06 | 1.424E-01 | 389 | 121705 | 122094 |
| 613.5 | genitourinary | Mastodynia | 1.50 | 0.41 | 0.26 | -0.15 | 0.89 | 1.430E-01 | 411 | 121130 | 121541 |
| 430.1 | circulatory system | Subarachnoid hemorrhage | 0.42 | -0.87 | 0.66 | -2.53 | 0.26 | 1.442E-01 | 224 | 122123 | 122347 |
| 172.11 | neoplasms | Melanomas of skin | 1.18 | 0.16 | 0.11 | -0.06 | 0.38 | 1.462E-01 | 4423 | 116196 | 120619 |

| | | | | | | | | | | | |
|---|---|---|---|---|---|---|---|---|---|---|---|
| 389.3 | sense organs | Degenerative and vascular disorders of ear | 1.41 | 0.34 | 0.22 | -0.13 | 0.76 | 1.472E-01 | 915 | 119939 | 120854 |
| 359 | neurological | Muscular dystrophies and other myopathies | 1.51 | 0.41 | 0.27 | -0.16 | 0.91 | 1.490E-01 | 505 | 121254 | 121759 |
| 705.8 | dermatologic | Hyperhidrosis | 1.46 | 0.38 | 0.25 | -0.15 | 0.84 | 1.493E-01 | 595 | 120119 | 120714 |
| 281.12 | hematopoietic | Other vitamin B12 deficiency anemia | 1.18 | 0.16 | 0.11 | -0.06 | 0.37 | 1.526E-01 | 4398 | 114976 | 119374 |
| 334.1 | neurological | Spinocerebellar disease | 1.86 | 0.62 | 0.39 | -0.27 | 1.32 | 1.537E-01 | 215 | 122101 | 122316 |
| 426.31 | circulatory system | Right bundle branch block | 1.21 | 0.19 | 0.13 | -0.07 | 0.44 | 1.544E-01 | 3178 | 115484 | 118662 |
| 586.4 | genitourinary | Stricture/obstruction of ureter | 1.32 | 0.28 | 0.19 | -0.11 | 0.63 | 1.548E-01 | 1339 | 119239 | 120578 |
| 535.6 | digestive | Duodenitis | 1.48 | 0.39 | 0.26 | -0.16 | 0.86 | 1.561E-01 | 633 | 119792 | 120425 |
| 585.34 | genitourinary | Chronic Kidney Disease, Stage IV | 1.18 | 0.17 | 0.12 | -0.07 | 0.39 | 1.570E-01 | 4273 | 116729 | 121002 |
| 395.4 | circulatory system | Nonrheumatic pulmonary valve disorders | 0.22 | -1.50 | 1.35 | -6.33 | 0.43 | 1.583E-01 | 135 | 121473 | 121608 |
| 767 | symptoms | Cervicocranial/Cervicobrachial syndrome | 2.03 | 0.71 | 0.44 | -0.33 | 1.52 | 1.586E-01 | 123 | 122312 | 122435 |
| 430 | circulatory system | Intracranial hemorrhage | 1.30 | 0.26 | 0.18 | -0.11 | 0.59 | 1.590E-01 | 1513 | 120095 | 121608 |
| 275.51 | endocrine/metabolic | Hypocalcemia | 1.45 | 0.37 | 0.25 | -0.16 | 0.84 | 1.591E-01 | 640 | 120629 | 121269 |
| 724.2 | musculoskeletal | Disorders of coccyx | 1.97 | 0.68 | 0.43 | -0.32 | 1.46 | 1.597E-01 | 142 | 122087 | 122229 |
| 731.1 | musculoskeletal | Osteitis deformans [Paget's disease of bone] | 0.38 | -0.96 | 0.80 | -3.13 | 0.31 | 1.611E-01 | 249 | 122211 | 122460 |
| 385.3 | sense organs | Cholesteatoma | 1.54 | 0.43 | 0.29 | -0.19 | 0.96 | 1.611E-01 | 438 | 121822 | 122260 |
| 962.1 | injuries & poisonings | Adrenal cortical steroids causing adverse effects in therapeutic use | 2.10 | 0.74 | 0.46 | -0.36 | 1.57 | 1.613E-01 | 126 | 122000 | 122126 |
| 960.1 | injuries & poisonings | Adverse effects of antibacterials (not penicillins) | 2.11 | 0.74 | 0.46 | -0.36 | 1.58 | 1.615E-01 | 124 | 121733 | 121857 |
| 253.2 | endocrine/metabolic | Pituitary hypofunction | 1.76 | 0.57 | 0.36 | -0.26 | 1.23 | 1.616E-01 | 241 | 122221 | 122462 |
| 270.32 | endocrine/metabolic | Paraproteinemia | 1.27 | 0.24 | 0.17 | -0.10 | 0.55 | 1.624E-01 | 1850 | 120216 | 122066 |
| 732 | musculoskeletal | Osteochondropathies | 1.83 | 0.60 | 0.39 | -0.28 | 1.33 | 1.634E-01 | 156 | 122023 | 122179 |
| 270.35 | endocrine/metabolic | Macroglobulinemia | 2.09 | 0.74 | 0.46 | -0.37 | 1.56 | 1.644E-01 | 138 | 122405 | 122543 |
| 303.3 | mental disorders | Psychogenic disorder | 1.79 | 0.58 | 0.38 | -0.28 | 1.28 | 1.653E-01 | 182 | 121785 | 121967 |

| | | | | | | | | | | | |
|---|---|---|---|---|---|---|---|---|---|---|---|
| 481 | respiratory | Influenza | 1.27 | 0.24 | 0.17 | -0.10 | 0.55 | 1.660E-01 | 1650 | 116755 | 118405 |
| 530.3 | digestive | Stricture and stenosis of esophagus | 1.26 | 0.23 | 0.16 | -0.10 | 0.53 | 1.661E-01 | 1889 | 118463 | 120352 |
| 473.4 | respiratory | Voice disturbance | 1.15 | 0.14 | 0.10 | -0.06 | 0.33 | 1.674E-01 | 5627 | 113120 | 118747 |
| 292.5 | mental disorders | Transient alteration of awareness | 1.61 | 0.47 | 0.32 | -0.22 | 1.06 | 1.675E-01 | 323 | 121019 | 121342 |
| 530.13 | digestive | Barrett's esophagus | 1.16 | 0.15 | 0.11 | -0.06 | 0.35 | 1.680E-01 | 4625 | 116641 | 121266 |
| 528.6 | digestive | Leukoplakia of oral mucosa | 1.56 | 0.44 | 0.30 | -0.21 | 0.99 | 1.684E-01 | 397 | 121448 | 121845 |
| 360.2 | sense organs | Progressive myopia | 1.82 | 0.60 | 0.39 | -0.30 | 1.31 | 1.689E-01 | 195 | 121886 | 122081 |
| 441.2 | circulatory system | Chronic vascular insufficiency of intestine | 2.07 | 0.73 | 0.46 | -0.38 | 1.57 | 1.702E-01 | 116 | 122224 | 122340 |
| 555.2 | digestive | Ulcerative colitis | 1.29 | 0.25 | 0.18 | -0.12 | 0.58 | 1.713E-01 | 1458 | 120003 | 121461 |
| 259.3 | endocrine/metabolic | Delay in sexual development and puberty NEC | 1.58 | 0.46 | 0.31 | -0.22 | 1.02 | 1.713E-01 | 419 | 121959 | 122378 |
| 696.42 | dermatologic | Psoriatic arthropathy | 1.39 | 0.33 | 0.23 | -0.15 | 0.75 | 1.719E-01 | 779 | 121578 | 122357 |
| 733.9 | musculoskeletal | Chondromalacia | 2.01 | 0.70 | 0.45 | -0.37 | 1.53 | 1.728E-01 | 104 | 122036 | 122140 |
| 274 | endocrine/metabolic | Gout and other crystal arthropathies | 0.91 | -0.09 | 0.07 | -0.23 | 0.04 | 1.755E-01 | 16660 | 101285 | 117945 |
| 963 | injuries & poisonings | Poisoning by primarily systemic agents | 1.40 | 0.34 | 0.23 | -0.16 | 0.77 | 1.755E-01 | 687 | 120551 | 121238 |
| 480.3 | respiratory | Pneumonia due to fungus (mycoses) | 1.52 | 0.42 | 0.29 | -0.21 | 0.95 | 1.763E-01 | 444 | 120799 | 121243 |
| 276.6 | endocrine/metabolic | Fluid overload | 1.26 | 0.23 | 0.17 | -0.11 | 0.55 | 1.764E-01 | 1754 | 117761 | 119515 |
| 695.8 | dermatologic | Other specified erythematous conditions | 1.25 | 0.23 | 0.16 | -0.11 | 0.53 | 1.779E-01 | 1787 | 117314 | 119101 |
| 557.1 | digestive | Celiac disease | 1.59 | 0.47 | 0.32 | -0.24 | 1.05 | 1.784E-01 | 318 | 122146 | 122464 |
| 276.11 | endocrine/metabolic | Hyperosmolality and/or hypernatremia | 0.61 | -0.49 | 0.39 | -1.38 | 0.20 | 1.791E-01 | 611 | 120546 | 121157 |
| 473 | respiratory | Diseases of the larynx and vocal cords | 1.13 | 0.12 | 0.09 | -0.06 | 0.30 | 1.842E-01 | 6617 | 111578 | 118195 |
| 426.4 | circulatory system | Anomalous atrioventricular excitation | 2.02 | 0.70 | 0.46 | -0.41 | 1.55 | 1.843E-01 | 114 | 122389 | 122503 |
| 686.2 | dermatologic | Impetigo | 0.40 | -0.92 | 0.79 | -3.09 | 0.36 | 1.843E-01 | 220 | 121270 | 121490 |
| 150 | neoplasms | Cancer of esophagus | 1.42 | 0.35 | 0.25 | -0.18 | 0.81 | 1.872E-01 | 700 | 121504 | 122204 |

| | | | | | | | | | | | |
|---|---|---|---|---|---|---|---|---|---|---|---|
| 980 | infectious diseases | Encounter for long-term (current) use of antibiotics | 1.36 | 0.30 | 0.22 | -0.16 | 0.71 | 1.877E-01 | 909 | 120504 | 121413 |
| 172.3 | neoplasms | Carcinoma in situ of skin | 1.22 | 0.20 | 0.14 | -0.10 | 0.47 | 1.890E-01 | 2657 | 116001 | 118658 |
| 229 | neoplasms | Benign neoplasm of unspecified sites | 0.40 | -0.91 | 0.80 | -3.08 | 0.37 | 1.895E-01 | 219 | 120887 | 121106 |
| 505 | respiratory | Other pulmonary inflamation or edema | 1.49 | 0.40 | 0.29 | -0.22 | 0.92 | 1.908E-01 | 485 | 120625 | 121110 |
| 53 | infectious diseases | Herpes zoster | 1.16 | 0.15 | 0.11 | -0.08 | 0.36 | 1.918E-01 | 4542 | 111602 | 116144 |
| 739 | musculoskeletal | Contracture of joint | 0.67 | -0.40 | 0.33 | -1.12 | 0.19 | 1.925E-01 | 758 | 120448 | 121206 |
| 585.32 | genitourinary | End stage renal disease | 1.24 | 0.22 | 0.16 | -0.12 | 0.52 | 1.929E-01 | 1997 | 119670 | 121667 |
| 701.4 | dermatologic | Keloid scar | 1.41 | 0.34 | 0.25 | -0.19 | 0.81 | 1.930E-01 | 656 | 120375 | 121031 |
| 686.3 | dermatologic | Pilonidal cyst | 1.77 | 0.57 | 0.40 | -0.34 | 1.30 | 1.936E-01 | 186 | 122086 | 122272 |
| 191.11 | neoplasms | Cancer of brain | 0.54 | -0.62 | 0.53 | -1.90 | 0.28 | 1.974E-01 | 358 | 121893 | 122251 |
| 529.1 | digestive | Glossitis | 0.25 | -1.39 | 1.33 | -6.23 | 0.54 | 1.991E-01 | 109 | 122091 | 122200 |
| 735.23 | musculoskeletal | Hallux rigidus | 1.21 | 0.19 | 0.15 | -0.11 | 0.47 | 1.994E-01 | 2141 | 118786 | 120927 |
| 627.22 | genitourinary | Need for Hormone replacement therapy (postmenopausal) | 1.52 | 0.42 | 0.31 | -0.25 | 1.00 | 2.004E-01 | 160 | 2588 | 2748 |
| 573 | digestive | Other disorders of liver | 1.22 | 0.20 | 0.15 | -0.11 | 0.49 | 2.006E-01 | 2041 | 116810 | 118851 |
| 301.2 | mental disorders | Antisocial/borderline personality disorder | 1.41 | 0.35 | 0.26 | -0.20 | 0.83 | 2.009E-01 | 386 | 121837 | 122223 |
| 427.61 | circulatory system | Supraventricular premature beats | 1.34 | 0.29 | 0.22 | -0.17 | 0.70 | 2.011E-01 | 939 | 118457 | 119396 |
| 738.4 | musculoskeletal | Acquired spondylolisthesis | 1.26 | 0.23 | 0.18 | -0.13 | 0.56 | 2.012E-01 | 1356 | 118714 | 120070 |
| 575 | digestive | Other biliary tract disease | 1.18 | 0.17 | 0.13 | -0.09 | 0.41 | 2.016E-01 | 2998 | 115762 | 118760 |
| 331 | neurological | Other cerebral degenerations | 1.36 | 0.31 | 0.23 | -0.18 | 0.73 | 2.018E-01 | 901 | 119097 | 119998 |
| 276.4 | endocrine/metabolic | Acid-base balance disorder | 1.18 | 0.17 | 0.13 | -0.09 | 0.41 | 2.031E-01 | 3212 | 113943 | 117155 |
| 442 | circulatory system | Other aneurysm | 1.10 | 0.10 | 0.08 | -0.05 | 0.25 | 2.040E-01 | 10854 | 107096 | 117950 |
| 194 | neoplasms | Cancer of other endocrine glands | 1.50 | 0.40 | 0.30 | -0.25 | 0.95 | 2.057E-01 | 422 | 121712 | 122134 |
| 573.4 | digestive | Acute and subacute necrosis of liver | 1.94 | 0.66 | 0.46 | -0.45 | 1.50 | 2.060E-01 | 127 | 121990 | 122117 |
| 530.9 | digestive | Heartburn | 1.31 | 0.27 | 0.20 | -0.16 | 0.65 | 2.081E-01 | 969 | 118227 | 119196 |
| 471 | respiratory | Nasal polyps | 1.28 | 0.24 | 0.19 | -0.15 | 0.59 | 2.093E-01 | 1352 | 120255 | 121607 |

| | | | | | | | | | | | |
|---|---|---|---|---|---|---|---|---|---|---|---|
| 472 | respiratory | Chronic pharyngitis and nasopharyngitis | 1.13 | 0.12 | 0.09 | -0.07 | 0.30 | 2.095E-01 | 6540 | 108467 | 115007 |
| 743.12 | musculoskeletal | Senile osteoporosis | 1.73 | 0.55 | 0.40 | -0.37 | 1.29 | 2.107E-01 | 120 | 122314 | 122434 |
| 225.1 | neoplasms | Benign neoplasm of brain, cranial nerves, meninges | 1.33 | 0.28 | 0.22 | -0.17 | 0.68 | 2.113E-01 | 925 | 121135 | 122060 |
| 729.3 | musculoskeletal | Panniculitis | 1.95 | 0.67 | 0.47 | -0.46 | 1.53 | 2.118E-01 | 106 | 122232 | 122338 |
| 295.2 | mental disorders | Paranoid disorders | 1.44 | 0.36 | 0.28 | -0.23 | 0.87 | 2.127E-01 | 464 | 121658 | 122122 |
| 452 | circulatory system | Other venous embolism and thrombosis | 1.11 | 0.10 | 0.08 | -0.06 | 0.26 | 2.136E-01 | 8624 | 109601 | 118225 |
| 394.2 | circulatory system | Mitral valve disease | 1.51 | 0.41 | 0.31 | -0.27 | 0.98 | 2.139E-01 | 422 | 121447 | 121869 |
| 526.4 | digestive | Temporomandibular joint disorders | 1.29 | 0.26 | 0.20 | -0.16 | 0.63 | 2.182E-01 | 959 | 119439 | 120398 |
| 172.21 | neoplasms | Basal cell carcinoma (new) | 1.09 | 0.09 | 0.07 | -0.05 | 0.22 | 2.185E-01 | 14106 | 101957 | 116063 |
| 496.1 | respiratory | Emphysema | 1.15 | 0.14 | 0.11 | -0.09 | 0.35 | 2.199E-01 | 4322 | 113593 | 117915 |
| 279.11 | endocrine/metabolic | Deficiency of humoral immunity | 0.50 | -0.70 | 0.62 | -2.27 | 0.37 | 2.215E-01 | 231 | 122183 | 122414 |
| 556.11 | digestive | Angiodysplasia of intestine (without mention of hemorrhage) | 0.50 | -0.69 | 0.62 | -2.25 | 0.35 | 2.220E-01 | 300 | 121424 | 121724 |
| 291 | mental disorders | Other specified nonpsychotic and/or transient mental disorders | 1.24 | 0.22 | 0.17 | -0.14 | 0.54 | 2.243E-01 | 1522 | 117278 | 118800 |
| 752 | congenital anomalies | Nervous system congenital anomalies | 1.76 | 0.57 | 0.42 | -0.42 | 1.33 | 2.273E-01 | 166 | 122061 | 122227 |
| 241.2 | endocrine/metabolic | Nontoxic multinodular goiter | 1.21 | 0.19 | 0.15 | -0.12 | 0.48 | 2.281E-01 | 1919 | 119603 | 121522 |
| 292.11 | mental disorders | Aphasia | 1.33 | 0.28 | 0.22 | -0.19 | 0.70 | 2.301E-01 | 895 | 120607 | 121502 |
| 516 | respiratory | Abnormal sputum | 1.22 | 0.20 | 0.16 | -0.13 | 0.50 | 2.324E-01 | 2034 | 117829 | 119863 |
| 857 | injuries & poisonings | Mechanical complication of unspecified genitourinary device, implant, and graft | 1.30 | 0.26 | 0.21 | -0.18 | 0.65 | 2.327E-01 | 1151 | 119593 | 120744 |
| 800.4 | injuries & poisonings | Fracture of patella | 1.40 | 0.34 | 0.27 | -0.24 | 0.83 | 2.327E-01 | 504 | 121767 | 122271 |
| 694.1 | dermatologic | Vitiligo | 1.47 | 0.38 | 0.30 | -0.28 | 0.94 | 2.334E-01 | 407 | 121943 | 122350 |
| 195.1 | neoplasms | Malignant neoplasm, other | 1.19 | 0.17 | 0.14 | -0.12 | 0.44 | 2.338E-01 | 2504 | 116103 | 118607 |
| 71.1 | infectious diseases | HIV infection, symptomatic | 1.44 | 0.37 | 0.29 | -0.27 | 0.90 | 2.352E-01 | 421 | 122077 | 122498 |

| | | | | | | | | | | | |
|---|---|---|---|---|---|---|---|---|---|---|---|
| 240 | endocrine/metabolic | Simple and unspecified goiter | 1.42 | 0.35 | 0.28 | -0.25 | 0.86 | 2.376E-01 | 488 | 121445 | 121933 |
| 295 | mental disorders | Schizophrenia and other psychotic disorders | 1.17 | 0.15 | 0.13 | -0.11 | 0.40 | 2.376E-01 | 2728 | 117038 | 119766 |
| 202.22 | neoplasms | Reticulosarcoma | 1.43 | 0.36 | 0.29 | -0.26 | 0.88 | 2.383E-01 | 484 | 121934 | 122418 |
| 579 | digestive | Other symptoms involving abdomen and pelvis | 1.21 | 0.19 | 0.16 | -0.13 | 0.48 | 2.388E-01 | 1868 | 116860 | 118728 |
| 781.2 | symptoms | Abnormal posture | 1.65 | 0.50 | 0.39 | -0.40 | 1.21 | 2.423E-01 | 213 | 122091 | 122304 |
| 592.2 | genitourinary | Urethritis and urethral syndrome | 1.86 | 0.62 | 0.47 | -0.52 | 1.49 | 2.431E-01 | 117 | 121848 | 121965 |
| 740.11 | musculoskeletal | Osteoarthrosis, localized, primary | 1.06 | 0.06 | 0.05 | -0.04 | 0.17 | 2.432E-01 | 23930 | 86061 | 109991 |
| 500 | respiratory | Lung disease due to external agents | 1.26 | 0.23 | 0.19 | -0.17 | 0.59 | 2.438E-01 | 1398 | 119844 | 121242 |
| 71 | infectious diseases | Human immunodeficiency virus [HIV] disease | 1.43 | 0.36 | 0.29 | -0.27 | 0.90 | 2.439E-01 | 427 | 122033 | 122460 |
| 271 | endocrine/metabolic | Disorders of carbohydrate transport and metabolism | 1.15 | 0.14 | 0.12 | -0.10 | 0.36 | 2.440E-01 | 3567 | 115351 | 118918 |
| 817 | injuries & poisonings | Concussion | 1.44 | 0.36 | 0.29 | -0.27 | 0.90 | 2.452E-01 | 390 | 120857 | 121247 |
| 502 | respiratory | Postinflammatory pulmonary fibrosis | 1.19 | 0.18 | 0.15 | -0.13 | 0.46 | 2.466E-01 | 2427 | 117383 | 119810 |
| 743.4 | musculoskeletal | Stress fracture | 1.50 | 0.40 | 0.33 | -0.32 | 1.00 | 2.497E-01 | 357 | 121442 | 121799 |
| 480.5 | respiratory | Bronchopneumonia and lung abscess | 1.45 | 0.37 | 0.30 | -0.29 | 0.92 | 2.500E-01 | 407 | 120835 | 121242 |
| 733.2 | musculoskeletal | Cyst of bone | 1.74 | 0.55 | 0.43 | -0.47 | 1.35 | 2.508E-01 | 140 | 121709 | 121849 |
| 709 | dermatologic | Diffuse diseases of connective tissue | 1.36 | 0.31 | 0.25 | -0.24 | 0.78 | 2.509E-01 | 545 | 120955 | 121500 |
| 433.6 | circulatory system | Acute, but ill-defined cerebrovascular disease | 1.14 | 0.13 | 0.11 | -0.09 | 0.34 | 2.511E-01 | 4840 | 115457 | 120297 |
| 289 | hematopoietic | Other diseases of blood and blood-forming organs | 1.24 | 0.22 | 0.18 | -0.16 | 0.56 | 2.514E-01 | 1450 | 118231 | 119681 |
| 425.11 | circulatory system | Hypertrophic obstructive cardiomyopathy | 1.57 | 0.45 | 0.36 | -0.36 | 1.11 | 2.521E-01 | 241 | 122160 | 122401 |
| 204 | neoplasms | Leukemia | 1.18 | 0.17 | 0.14 | -0.13 | 0.44 | 2.525E-01 | 2616 | 119353 | 121969 |
| 174.11 | neoplasms | Malignant neoplasm of female breast | 1.30 | 0.27 | 0.23 | -0.20 | 0.70 | 2.533E-01 | 339 | 2515 | 2854 |

| | | | | | | | | | | | |
|---|---|---|---|---|---|---|---|---|---|---|---|
| 110.13 | infectious diseases | Dermatophytosis of the body | 1.19 | 0.18 | 0.15 | -0.13 | 0.46 | 2.544E-01 | 2308 | 114613 | 116921 |
| 440.2 | circulatory system | Atherosclerosis of the extremities | 1.13 | 0.12 | 0.10 | -0.09 | 0.31 | 2.557E-01 | 5502 | 112063 | 117565 |
| 433.11 | circulatory system | Occlusion of cerebral arteries, with cerebral infarction | 0.72 | -0.33 | 0.31 | -1.01 | 0.22 | 2.563E-01 | 899 | 120377 | 121276 |
| 429.9 | circulatory system | Cardiac complications, not elsewhere classified | 1.52 | 0.42 | 0.34 | -0.35 | 1.04 | 2.581E-01 | 350 | 120028 | 120378 |
| 227.2 | neoplasms | Benign neoplasm of parathyroid gland | 1.69 | 0.52 | 0.42 | -0.46 | 1.29 | 2.593E-01 | 172 | 122241 | 122413 |
| 736.4 | musculoskeletal | Genu valgum or varum (acquired) | 1.52 | 0.42 | 0.35 | -0.35 | 1.05 | 2.595E-01 | 285 | 121688 | 121973 |
| 430.2 | circulatory system | Intracerebral hemorrhage | 1.41 | 0.34 | 0.29 | -0.28 | 0.86 | 2.597E-01 | 503 | 121580 | 122083 |
| 327.41 | neurological | Organic or persistent insomnia | 1.26 | 0.23 | 0.20 | -0.18 | 0.60 | 2.613E-01 | 1057 | 119964 | 121021 |
| 217.1 | neoplasms | Nevus, non-neoplastic | 1.14 | 0.13 | 0.11 | -0.10 | 0.34 | 2.631E-01 | 4119 | 109188 | 113307 |
| 585.2 | genitourinary | Renal failure NOS | 1.19 | 0.18 | 0.15 | -0.14 | 0.46 | 2.653E-01 | 2328 | 117277 | 119605 |
| 418.1 | circulatory system | Precordial pain | 1.34 | 0.30 | 0.25 | -0.25 | 0.76 | 2.657E-01 | 605 | 119702 | 120307 |
| 772 | symptoms | Symptoms of the muscles | 1.40 | 0.34 | 0.29 | -0.29 | 0.86 | 2.664E-01 | 478 | 120654 | 121132 |
| 588.2 | genitourinary | Secondary hyperparathyroidism (of renal origin) | 1.20 | 0.18 | 0.16 | -0.15 | 0.48 | 2.673E-01 | 2188 | 119210 | 121398 |
| 736.2 | musculoskeletal | Acquired deformities of finger | 0.62 | -0.48 | 0.47 | -1.58 | 0.33 | 2.694E-01 | 382 | 121631 | 122013 |
| 172.22 | neoplasms | Squamous cell carcinoma | 1.10 | 0.09 | 0.08 | -0.07 | 0.26 | 2.710E-01 | 9145 | 107846 | 116991 |
| 967 | injuries & poisonings | Adverse effects of sedatives or other central nervous system depressants and anesthetics | 1.77 | 0.57 | 0.47 | -0.55 | 1.43 | 2.717E-01 | 113 | 121760 | 121873 |
| 450 | circulatory system | Noninfectious disorders of lymphatic channels | 1.20 | 0.19 | 0.16 | -0.15 | 0.49 | 2.722E-01 | 1710 | 119310 | 121020 |
| 433.8 | circulatory system | Late effects of cerebrovascular disease | 1.12 | 0.11 | 0.10 | -0.09 | 0.31 | 2.729E-01 | 5492 | 112955 | 118447 |
| 197 | neoplasms | Chemotherapy | 1.14 | 0.13 | 0.12 | -0.11 | 0.36 | 2.735E-01 | 3642 | 117100 | 120742 |
| 334 | neurological | Degenerative disease of the spinal cord | 1.27 | 0.24 | 0.21 | -0.20 | 0.63 | 2.744E-01 | 985 | 120107 | 121092 |
| 613.9 | genitourinary | Breast disorder NOS | 1.56 | 0.45 | 0.38 | -0.42 | 1.16 | 2.754E-01 | 126 | 122019 | 122145 |

| | | | | | | | | | | | |
|---|---|---|---|---|---|---|---|---|---|---|---|
| 747.2 | congenital anomalies | Congenital anomalies of peripheral vascular system | 1.46 | 0.38 | 0.33 | -0.34 | 0.97 | 2.761E-01 | 378 | 121458 | 121836 |
| 830 | injuries & poisonings | Dislocation | 1.15 | 0.14 | 0.12 | -0.12 | 0.37 | 2.772E-01 | 3018 | 116257 | 119275 |
| 279 | endocrine/metabolic | Disorders involving the immune mechanism | 1.38 | 0.32 | 0.28 | -0.29 | 0.84 | 2.801E-01 | 454 | 121570 | 122024 |
| 289.1 | hematopoietic | Myelofibrosis | 1.87 | 0.63 | 0.52 | -0.65 | 1.55 | 2.818E-01 | 114 | 122445 | 122559 |
| 282 | hematopoietic | Hereditary hemolytic anemias | 1.53 | 0.43 | 0.37 | -0.41 | 1.10 | 2.825E-01 | 292 | 121816 | 122108 |
| 580.1 | genitourinary | Glomerulonephritis | 1.49 | 0.40 | 0.34 | -0.37 | 1.02 | 2.830E-01 | 333 | 121828 | 122161 |
| 370.31 | sense organs | Keratoconjunctivitis sicca | 1.22 | 0.20 | 0.18 | -0.17 | 0.53 | 2.830E-01 | 1455 | 117947 | 119402 |
| 291.4 | mental disorders | Specific nonpsychotic mental disorders due to brain damage | 1.34 | 0.29 | 0.26 | -0.27 | 0.77 | 2.835E-01 | 597 | 120926 | 121523 |
| 225 | neoplasms | Benign neoplasm of brain and other parts of nervous system | 1.27 | 0.24 | 0.21 | -0.21 | 0.64 | 2.839E-01 | 966 | 121048 | 122014 |
| 290.11 | mental disorders | Alzheimer's disease | 0.83 | -0.18 | 0.17 | -0.54 | 0.14 | 2.840E-01 | 2638 | 118619 | 121257 |
| 245.21 | endocrine/metabolic | Chronic lymphocytic thyroiditis | 1.53 | 0.42 | 0.37 | -0.40 | 1.10 | 2.847E-01 | 194 | 122230 | 122424 |
| 366.2 | sense organs | Senile cataract | 1.06 | 0.06 | 0.06 | -0.05 | 0.18 | 2.896E-01 | 91358 | 20000 | 111358 |
| 530.14 | digestive | Reflux esophagitis | 1.13 | 0.12 | 0.11 | -0.11 | 0.33 | 2.920E-01 | 4065 | 113456 | 117521 |
| 695.4 | dermatologic | Lupus (localized and systemic) | 0.63 | -0.47 | 0.47 | -1.58 | 0.36 | 2.931E-01 | 306 | 122118 | 122424 |
| 275.53 | endocrine/metabolic | Disorders of phosphorus metabolism | 1.29 | 0.25 | 0.23 | -0.24 | 0.68 | 2.935E-01 | 829 | 119970 | 120799 |
| 727.7 | musculoskeletal | Contracture of tendon (sheath) | 1.72 | 0.54 | 0.46 | -0.57 | 1.37 | 2.937E-01 | 145 | 122026 | 122171 |
| 528.5 | digestive | Diseases of lips | 1.47 | 0.38 | 0.34 | -0.38 | 1.00 | 2.943E-01 | 353 | 121182 | 121535 |
| 613.7 | genitourinary | Other signs and symptoms in breast | 1.44 | 0.37 | 0.33 | -0.36 | 0.99 | 2.948E-01 | 161 | 121878 | 122039 |
| 526.41 | digestive | Temporomandibular joint disorder, unspecified | 1.27 | 0.24 | 0.22 | -0.23 | 0.66 | 2.954E-01 | 752 | 120120 | 120872 |
| 716.2 | musculoskeletal | Unspecified monoarthritis | 0.71 | -0.34 | 0.34 | -1.11 | 0.27 | 2.965E-01 | 645 | 120553 | 121198 |
| 327.5 | neurological | Parasomnia | 1.17 | 0.16 | 0.15 | -0.15 | 0.44 | 2.971E-01 | 2102 | 117405 | 119507 |
| 752.2 | congenital anomalies | Other specified congenital anomalies of nervous system | 1.82 | 0.60 | 0.52 | -0.68 | 1.52 | 2.992E-01 | 116 | 122238 | 122354 |

| | | | | | | | | | | | |
|---|---|---|---|---|---|---|---|---|---|---|---|
| 286.12 | hematopoietic | Congenital deficiency of other clotting factors (including factor VII) | 0.48 | -0.74 | 0.78 | -2.92 | 0.55 | 2.994E-01 | 150 | 122288 | 122438 |
| 560.3 | digestive | Peritoneal or intestinal adhesions | 0.56 | -0.59 | 0.62 | -2.15 | 0.46 | 3.020E-01 | 249 | 121971 | 122220 |
| 701.5 | dermatologic | Abnormal granulation tissue | 0.48 | -0.73 | 0.79 | -2.91 | 0.55 | 3.027E-01 | 182 | 121461 | 121643 |
| 217 | neoplasms | Vascular hamartomas and non-neoplastic nevi | 1.12 | 0.12 | 0.11 | -0.11 | 0.33 | 3.049E-01 | 4265 | 108880 | 113145 |
| 377.1 | sense organs | Optic atrophy | 1.19 | 0.17 | 0.16 | -0.17 | 0.48 | 3.049E-01 | 2041 | 117976 | 120017 |
| 609.2 | genitourinary | Abnormal spermatozoa | 0.66 | -0.41 | 0.43 | -1.39 | 0.33 | 3.060E-01 | 489 | 118514 | 119003 |
| 313.1 | mental disorders | Attention deficit hyperactivity disorder | 1.24 | 0.21 | 0.20 | -0.21 | 0.59 | 3.077E-01 | 870 | 121438 | 122308 |
| 519.2 | respiratory | Respiratory complications | 1.68 | 0.52 | 0.46 | -0.59 | 1.35 | 3.088E-01 | 155 | 121503 | 121658 |
| 302 | mental disorders | Sexual and gender identity disorders | 1.24 | 0.21 | 0.20 | -0.21 | 0.59 | 3.094E-01 | 1117 | 119703 | 120820 |
| 133 | infectious diseases | Arthropod-borne diseases | 0.47 | -0.75 | 0.80 | -2.95 | 0.59 | 3.097E-01 | 132 | 122210 | 122342 |
| 174 | neoplasms | Breast cancer | 1.24 | 0.21 | 0.21 | -0.21 | 0.60 | 3.106E-01 | 549 | 121739 | 122288 |
| 153.3 | neoplasms | Malignant neoplasm of rectum, rectosigmoid junction, and anus | 1.24 | 0.22 | 0.21 | -0.22 | 0.60 | 3.117E-01 | 1126 | 120854 | 121980 |
| 835 | injuries & poisonings | Internal derangement of knee | 1.13 | 0.12 | 0.12 | -0.12 | 0.34 | 3.127E-01 | 3490 | 114888 | 118378 |
| 283 | hematopoietic | Acquired hemolytic anemias | 1.53 | 0.43 | 0.39 | -0.48 | 1.14 | 3.143E-01 | 220 | 122062 | 122282 |
| 271.3 | endocrine/metabolic | Intestinal disaccharidase deficiencies and disaccharide malabsorption | 1.13 | 0.12 | 0.12 | -0.12 | 0.35 | 3.159E-01 | 3420 | 115641 | 119061 |
| 589 | genitourinary | Abnormal results of function study of kidney | 0.71 | -0.35 | 0.37 | -1.17 | 0.30 | 3.168E-01 | 618 | 119550 | 120168 |
| 573.9 | digestive | Abnormal serum enzyme levels | 1.21 | 0.19 | 0.18 | -0.19 | 0.53 | 3.169E-01 | 1436 | 117505 | 118941 |
| 149.3 | neoplasms | Cancer of hypopharynx | 0.48 | -0.73 | 0.79 | -2.91 | 0.60 | 3.199E-01 | 139 | 122349 | 122488 |
| 174.1 | neoplasms | Breast cancer [female] | 1.26 | 0.23 | 0.23 | -0.23 | 0.66 | 3.205E-01 | 354 | 2505 | 2859 |
| 245 | endocrine/metabolic | Thyroiditis | 1.36 | 0.30 | 0.29 | -0.33 | 0.84 | 3.211E-01 | 378 | 121802 | 122180 |
| 772.4 | symptoms | Rhabdomyolysis | 1.37 | 0.31 | 0.30 | -0.34 | 0.86 | 3.224E-01 | 460 | 121524 | 121984 |

| | | | | | | | | | | | |
|---|---|---|---|---|---|---|---|---|---|---|---|
| 792 | symptoms | Abnormal Papanicolaou smear of cervix and cervical HPV | 1.37 | 0.32 | 0.31 | -0.35 | 0.91 | 3.230E-01 | 143 | 2642 | 2785 |
| 381.9 | sense organs | Otorrhea | 1.43 | 0.36 | 0.34 | -0.41 | 0.98 | 3.241E-01 | 350 | 121345 | 121695 |
| 295.3 | mental disorders | Psychosis | 1.18 | 0.17 | 0.17 | -0.18 | 0.48 | 3.243E-01 | 1605 | 118603 | 120208 |
| 568.1 | digestive | Peritoneal adhesions (postoperative) (postinfection) | 1.34 | 0.30 | 0.29 | -0.33 | 0.82 | 3.254E-01 | 511 | 120912 | 121423 |
| 198.6 | neoplasms | Secondary malignancy of bone | 1.18 | 0.16 | 0.16 | -0.17 | 0.47 | 3.260E-01 | 2022 | 119860 | 121882 |
| 681.3 | dermatologic | Cellulitis and abscess of arm/hand | 1.18 | 0.16 | 0.16 | -0.17 | 0.47 | 3.264E-01 | 1790 | 117389 | 119179 |
| 586.3 | genitourinary | Vascular disorders of kidney/hypertrophy | 1.75 | 0.56 | 0.52 | -0.72 | 1.48 | 3.317E-01 | 118 | 122188 | 122306 |
| 790.1 | symptoms | Elevated sedimentation rate | 0.50 | -0.69 | 0.79 | -2.86 | 0.59 | 3.330E-01 | 178 | 121940 | 122118 |
| 200 | neoplasms | Myeloproliferative disease | 1.16 | 0.15 | 0.15 | -0.16 | 0.44 | 3.330E-01 | 2226 | 119004 | 121230 |
| 358.1 | neurological | Myasthenia gravis | 1.37 | 0.32 | 0.31 | -0.36 | 0.88 | 3.335E-01 | 472 | 121992 | 122464 |
| 276.41 | endocrine/metabolic | Acidosis | 1.15 | 0.14 | 0.14 | -0.15 | 0.40 | 3.341E-01 | 2680 | 115336 | 118016 |
| 312 | mental disorders | Conduct disorders | 1.26 | 0.23 | 0.23 | -0.26 | 0.66 | 3.343E-01 | 742 | 121120 | 121862 |
| 349 | neurological | Other and unspecified disorders of the nervous system | 0.79 | -0.24 | 0.26 | -0.79 | 0.23 | 3.343E-01 | 1106 | 119500 | 120606 |
| 451 | circulatory system | Phlebitis and thrombophlebitis | 1.22 | 0.20 | 0.20 | -0.22 | 0.58 | 3.351E-01 | 1176 | 119281 | 120457 |
| 526.5 | digestive | Inflammatory conditions of jaw | 1.58 | 0.46 | 0.44 | -0.57 | 1.26 | 3.355E-01 | 158 | 121851 | 122009 |
| 279.1 | endocrine/metabolic | Immunity deficiency | 0.65 | -0.44 | 0.48 | -1.56 | 0.41 | 3.373E-01 | 304 | 121915 | 122219 |
| 260.2 | endocrine/metabolic | severe protein-calorie malnutrition | 0.75 | -0.28 | 0.31 | -0.96 | 0.28 | 3.429E-01 | 758 | 120466 | 121224 |
| 334.2 | neurological | Anterior horn cell disease | 0.58 | -0.54 | 0.62 | -2.11 | 0.50 | 3.436E-01 | 250 | 122107 | 122357 |
| 731 | musculoskeletal | Osteitis deformans and osteopathies associated with other disorders classified elsewhere | 1.36 | 0.31 | 0.31 | -0.37 | 0.87 | 3.439E-01 | 460 | 121637 | 122097 |
| 704.1 | dermatologic | Alopecia | 1.36 | 0.31 | 0.31 | -0.37 | 0.88 | 3.447E-01 | 312 | 121495 | 121807 |

| | | | | | | | | | | | |
|---|---|---|---|---|---|---|---|---|---|---|---|
| 441 | circulatory system | Vascular insufficiency of intestine | 1.31 | 0.27 | 0.28 | -0.32 | 0.78 | 3.452E-01 | 551 | 121341 | 121892 |
| 740.12 | musculoskeletal | Osteoarthrosis, localized, secondary | 1.14 | 0.13 | 0.14 | -0.15 | 0.39 | 3.455E-01 | 2575 | 116168 | 118743 |
| 394.1 | circulatory system | Mitral valve stenosis and aortic valve stenosis | 1.27 | 0.24 | 0.24 | -0.28 | 0.68 | 3.470E-01 | 882 | 119450 | 120332 |
| 41.8 | infectious diseases | H. pylori | 1.26 | 0.23 | 0.24 | -0.27 | 0.67 | 3.477E-01 | 811 | 120447 | 121258 |
| 577.2 | digestive | Chronic pancreatitis | 1.29 | 0.25 | 0.26 | -0.30 | 0.72 | 3.494E-01 | 672 | 121192 | 121864 |
| 686.1 | dermatologic | Carbuncle and furuncle | 1.24 | 0.21 | 0.22 | -0.25 | 0.62 | 3.501E-01 | 912 | 118823 | 119735 |
| 8.52 | infectious diseases | Intestinal infection due to C. difficile | 1.18 | 0.17 | 0.17 | -0.19 | 0.49 | 3.506E-01 | 1659 | 119593 | 121252 |
| 158 | neoplasms | Neoplasm of unspecified nature of digestive system | 0.76 | -0.28 | 0.31 | -0.96 | 0.28 | 3.523E-01 | 786 | 119979 | 120765 |
| 362.27 | sense organs | Drusen (degenerative) of retina | 1.11 | 0.11 | 0.11 | -0.12 | 0.32 | 3.530E-01 | 4551 | 111529 | 116080 |
| 202.21 | neoplasms | Nodular lymphoma | 1.36 | 0.30 | 0.31 | -0.38 | 0.87 | 3.533E-01 | 439 | 121966 | 122405 |
| 189.2 | neoplasms | Cancer of bladder | 0.90 | -0.11 | 0.12 | -0.35 | 0.12 | 3.550E-01 | 5372 | 115903 | 121275 |
| 427.5 | circulatory system | Arrhythmia (cardiac) NOS | 0.93 | -0.07 | 0.08 | -0.22 | 0.08 | 3.580E-01 | 11799 | 98292 | 110091 |
| 386.1 | sense organs | Meniere's disease | 1.28 | 0.25 | 0.26 | -0.31 | 0.72 | 3.595E-01 | 636 | 121552 | 122188 |
| 580.4 | genitourinary | Renal sclerosis, NOS | 1.59 | 0.46 | 0.46 | -0.65 | 1.29 | 3.600E-01 | 154 | 121976 | 122130 |
| 754 | congenital anomalies | Congenital musculoskeletal deformities of spine | 1.30 | 0.26 | 0.28 | -0.34 | 0.77 | 3.626E-01 | 521 | 120950 | 121471 |
| 362.22 | sense organs | Macular degeneration, wet | 1.11 | 0.10 | 0.11 | -0.12 | 0.31 | 3.630E-01 | 5398 | 115277 | 120675 |
| 559 | digestive | Ileostomy status | 1.30 | 0.26 | 0.27 | -0.33 | 0.76 | 3.639E-01 | 597 | 121762 | 122359 |
| 627.1 | genitourinary | Postmenopausal bleeding | 1.30 | 0.26 | 0.28 | -0.34 | 0.80 | 3.658E-01 | 190 | 2612 | 2802 |
| 342 | neurological | Hemiplegia | 1.27 | 0.24 | 0.26 | -0.31 | 0.72 | 3.687E-01 | 665 | 120742 | 121407 |
| 819 | injuries & poisonings | Skull and face fracture and other intercranial injury | 1.14 | 0.13 | 0.15 | -0.16 | 0.41 | 3.699E-01 | 2167 | 117709 | 119876 |
| 345.11 | neurological | Generalized convulsive epilepsy | 1.46 | 0.38 | 0.40 | -0.54 | 1.10 | 3.749E-01 | 216 | 122056 | 122272 |
| 286.81 | hematopoietic | Primary hypercoagulable state | 1.27 | 0.24 | 0.26 | -0.32 | 0.72 | 3.763E-01 | 584 | 121545 | 122129 |
| 315 | mental disorders | Develomental delays and disorders | 1.30 | 0.26 | 0.29 | -0.36 | 0.79 | 3.768E-01 | 531 | 120606 | 121137 |

| | | | | | | | | | | | |
|---|---|---|---|---|---|---|---|---|---|---|---|
| 577.3 | digestive | Cyst and pseudocyst of pancreas | 1.23 | 0.20 | 0.22 | -0.27 | 0.62 | 3.777E-01 | 985 | 120928 | 121913 |
| 145 | neoplasms | Cancer of mouth | 1.17 | 0.16 | 0.18 | -0.21 | 0.49 | 3.785E-01 | 1595 | 120007 | 121602 |
| 277.51 | endocrine/metabolic | Lipoprotein disorders | 1.38 | 0.32 | 0.35 | -0.46 | 0.96 | 3.810E-01 | 295 | 121685 | 121980 |
| 250.21 | endocrine/metabolic | Type 2 diabetes with ketoacidosis | 1.48 | 0.39 | 0.42 | -0.59 | 1.16 | 3.837E-01 | 202 | 122103 | 122305 |
| 352.2 | neurological | Facial nerve disorders [CN7] | 1.19 | 0.18 | 0.20 | -0.24 | 0.55 | 3.852E-01 | 1169 | 120549 | 121718 |
| 278.3 | endocrine/metabolic | Localized adiposity | 1.54 | 0.43 | 0.47 | -0.68 | 1.28 | 3.853E-01 | 125 | 122148 | 122273 |
| 618.1 | genitourinary | Prolapse of vaginal walls | 1.27 | 0.24 | 0.27 | -0.33 | 0.76 | 3.871E-01 | 239 | 2528 | 2767 |
| 528.1 | digestive | Stomatitis and mucositis | 1.31 | 0.27 | 0.30 | -0.38 | 0.81 | 3.900E-01 | 465 | 120500 | 120965 |
| 500.1 | respiratory | Extrinsic allergic alveolitis | 1.62 | 0.48 | 0.52 | -0.81 | 1.43 | 3.935E-01 | 105 | 122328 | 122433 |
| 189.21 | neoplasms | Malignant neoplasm of bladder | 0.90 | -0.10 | 0.12 | -0.35 | 0.13 | 3.938E-01 | 4957 | 116580 | 121537 |
| 245.2 | endocrine/metabolic | Chronic thyroiditis | 1.39 | 0.33 | 0.37 | -0.49 | 1.00 | 3.962E-01 | 213 | 122174 | 122387 |
| 910 | injuries & poisonings | Superficial injury, infected | 1.61 | 0.48 | 0.52 | -0.80 | 1.40 | 3.996E-01 | 125 | 121555 | 121680 |
| 286.8 | hematopoietic | Hypercoagulable state | 1.25 | 0.22 | 0.25 | -0.32 | 0.69 | 4.015E-01 | 639 | 121386 | 122025 |
| 414 | circulatory system | Other forms of chronic heart disease | 0.91 | -0.10 | 0.12 | -0.34 | 0.13 | 4.028E-01 | 5201 | 110708 | 115909 |
| 253.4 | endocrine/metabolic | Anterior pituitary disorders | 1.26 | 0.23 | 0.27 | -0.34 | 0.72 | 4.029E-01 | 609 | 121357 | 121966 |
| 593.1 | genitourinary | Gross hematuria | 1.09 | 0.08 | 0.10 | -0.12 | 0.27 | 4.038E-01 | 6143 | 112252 | 118395 |
| 513.8 | respiratory | Disorders of diaphragm | 1.27 | 0.24 | 0.27 | -0.36 | 0.74 | 4.057E-01 | 632 | 120909 | 121541 |
| 550.6 | digestive | Incisional hernia | 0.85 | -0.16 | 0.20 | -0.58 | 0.21 | 4.065E-01 | 1650 | 119905 | 121555 |
| 747.12 | congenital anomalies | Valvular heart disease/ heart chambers | 1.36 | 0.30 | 0.35 | -0.48 | 0.94 | 4.078E-01 | 325 | 121619 | 121944 |
| 70.4 | infectious diseases | Chronic hepatitis | 1.33 | 0.29 | 0.33 | -0.45 | 0.90 | 4.093E-01 | 299 | 121861 | 122160 |
| 709.2 | dermatologic | Sicca syndrome | 1.33 | 0.29 | 0.34 | -0.46 | 0.90 | 4.145E-01 | 293 | 121508 | 121801 |
| 198.2 | neoplasms | Secondary malignancy of respiratory organs | 1.20 | 0.18 | 0.22 | -0.28 | 0.59 | 4.146E-01 | 1057 | 120737 | 121794 |
| 242.1 | endocrine/metabolic | Graves' disease | 1.30 | 0.26 | 0.31 | -0.41 | 0.82 | 4.150E-01 | 380 | 121966 | 122346 |
| 346.2 | neurological | Nonspecific abnormal results of function study of brain and central nervous system | 0.56 | -0.58 | 0.78 | -2.76 | 0.71 | 4.168E-01 | 135 | 121664 | 121799 |
| 716.1 | musculoskeletal | Unspecified polyarthropathy or polyarthritis | 0.70 | -0.36 | 0.47 | -1.46 | 0.46 | 4.175E-01 | 324 | 121817 | 122141 |

| | | | | | | | | | | | |
|---|---|---|---|---|---|---|---|---|---|---|---|
| 286.7 | hematopoietic | Other and unspecified coagulation defects | 1.15 | 0.14 | 0.17 | -0.21 | 0.46 | 4.181E-01 | 1959 | 118777 | 120736 |
| 871.2 | injuries & poisonings | Open wound of finger(s) | 1.17 | 0.16 | 0.19 | -0.24 | 0.52 | 4.182E-01 | 1239 | 118029 | 119268 |
| 389 | sense organs | Hearing loss | 1.04 | 0.04 | 0.05 | -0.06 | 0.14 | 4.194E-01 | 80615 | 32710 | 113325 |
| 564.9 | digestive | Personal history of diseases of digestive system | 1.24 | 0.21 | 0.26 | -0.34 | 0.69 | 4.211E-01 | 732 | 119044 | 119776 |
| 284.1 | hematopoietic | Pancytopenia | 1.16 | 0.14 | 0.18 | -0.22 | 0.47 | 4.226E-01 | 1688 | 119576 | 121264 |
| 331.1 | neurological | Hydrocephalus | 1.36 | 0.31 | 0.36 | -0.51 | 0.96 | 4.227E-01 | 347 | 121955 | 122302 |
| 727.5 | musculoskeletal | Rupture of synovium | 1.24 | 0.21 | 0.26 | -0.34 | 0.69 | 4.250E-01 | 670 | 120526 | 121196 |
| 803 | injuries & poisonings | Fracture of upper limb | 1.09 | 0.09 | 0.11 | -0.14 | 0.30 | 4.331E-01 | 3849 | 117236 | 121085 |
| 446.5 | circulatory system | Giant cell arteritis | 1.30 | 0.27 | 0.33 | -0.45 | 0.85 | 4.332E-01 | 438 | 121868 | 122306 |
| 189 | neoplasms | Cancer of urinary organs (incl. kidney and bladder) | 0.93 | -0.08 | 0.10 | -0.28 | 0.11 | 4.373E-01 | 7425 | 113348 | 120773 |
| 282.8 | hematopoietic | Other hemoglobinopathies | 1.42 | 0.35 | 0.43 | -0.65 | 1.12 | 4.396E-01 | 244 | 122029 | 122273 |
| 444 | circulatory system | Arterial embolism and thrombosis | 0.84 | -0.17 | 0.23 | -0.66 | 0.25 | 4.406E-01 | 1286 | 119449 | 120735 |
| 614.52 | genitourinary | Vaginitis and vulvovaginitis | 1.27 | 0.24 | 0.31 | -0.41 | 0.81 | 4.421E-01 | 169 | 2448 | 2617 |
| 958 | injuries & poisonings | Certain early complications of trauma or procedure | 1.37 | 0.32 | 0.39 | -0.58 | 1.02 | 4.422E-01 | 253 | 121656 | 121909 |
| 290.16 | mental disorders | Vascular dementia | 1.13 | 0.12 | 0.16 | -0.20 | 0.42 | 4.432E-01 | 2392 | 118527 | 120919 |
| 756 | congenital anomalies | Other congenital musculoskeletal anomalies | 1.28 | 0.25 | 0.32 | -0.45 | 0.83 | 4.484E-01 | 351 | 120875 | 121226 |
| 870.2 | injuries & poisonings | Open wound of ear | 1.52 | 0.42 | 0.52 | -0.86 | 1.34 | 4.495E-01 | 134 | 121972 | 122106 |
| 165 | neoplasms | Cancer within the respiratory system | 0.92 | -0.08 | 0.11 | -0.32 | 0.13 | 4.520E-01 | 5256 | 115732 | 120988 |
| 442.11 | circulatory system | Abdominal aortic aneurysm | 1.07 | 0.07 | 0.09 | -0.12 | 0.25 | 4.535E-01 | 7610 | 111679 | 119289 |
| 530.6 | digestive | Diverticulum of esophagus, acquired | 1.36 | 0.30 | 0.39 | -0.59 | 1.00 | 4.553E-01 | 300 | 122020 | 122320 |
| 198.4 | neoplasms | Secondary malignant neoplasm of liver | 0.82 | -0.20 | 0.27 | -0.79 | 0.30 | 4.556E-01 | 968 | 121145 | 122113 |
| 149.9 | neoplasms | Cancer of of nasal cavities | 0.59 | -0.53 | 0.77 | -2.70 | 0.74 | 4.559E-01 | 149 | 122215 | 122364 |
| 8.5 | infectious diseases | Bacterial enteritis | 1.13 | 0.13 | 0.17 | -0.22 | 0.44 | 4.568E-01 | 1868 | 118908 | 120776 |
| 210 | neoplasms | Benign neoplasm of lip, oral cavity, and pharynx | 1.18 | 0.16 | 0.22 | -0.29 | 0.57 | 4.581E-01 | 979 | 119932 | 120911 |

| | | | | | | | | | | | |
|---|---|---|---|---|---|---|---|---|---|---|---|
| 442.1 | circulatory system | Aortic aneurysm | 1.06 | 0.06 | 0.08 | -0.10 | 0.22 | 4.589E-01 | 9813 | 108822 | 118635 |
| 500.2 | respiratory | Pneumoconiosis | 1.19 | 0.17 | 0.23 | -0.31 | 0.59 | 4.605E-01 | 1086 | 120762 | 121848 |
| 187 | neoplasms | Cancer of other male genital organs | 1.22 | 0.20 | 0.27 | -0.37 | 0.69 | 4.611E-01 | 699 | 117834 | 118533 |
| 348.8 | neurological | Encephalopathy, not elsewhere classified | 0.88 | -0.13 | 0.18 | -0.50 | 0.20 | 4.619E-01 | 2146 | 117669 | 119815 |
| 433.2 | circulatory system | Occlusion of cerebral arteries | 1.07 | 0.06 | 0.09 | -0.11 | 0.23 | 4.628E-01 | 8037 | 110489 | 118526 |
| 433.21 | circulatory system | Cerebral artery occlusion, with cerebral infarction | 1.07 | 0.07 | 0.09 | -0.11 | 0.23 | 4.633E-01 | 7866 | 110816 | 118682 |
| 772.1 | symptoms | Muscular wasting and disuse atrophy | 1.25 | 0.23 | 0.30 | -0.43 | 0.77 | 4.675E-01 | 472 | 121087 | 121559 |
| 359.2 | neurological | Myopathy | 1.26 | 0.23 | 0.31 | -0.45 | 0.80 | 4.730E-01 | 452 | 121365 | 121817 |
| 211 | neoplasms | Benign neoplasm of other parts of digestive system | 0.85 | -0.16 | 0.24 | -0.66 | 0.27 | 4.777E-01 | 1206 | 118380 | 119586 |
| 290.2 | mental disorders | Delirium due to conditions classified elsewhere | 1.14 | 0.13 | 0.18 | -0.24 | 0.46 | 4.785E-01 | 1695 | 118638 | 120333 |
| 459.7 | circulatory system | Blood vessel replaced | 0.83 | -0.19 | 0.27 | -0.78 | 0.31 | 4.788E-01 | 960 | 120073 | 121033 |
| 750.1 | congenital anomalies | Upper gastrointestinal congenital anomalies | 1.28 | 0.25 | 0.34 | -0.52 | 0.87 | 4.815E-01 | 383 | 121045 | 121428 |
| 695.2 | dermatologic | Bullous dermatoses | 1.28 | 0.25 | 0.34 | -0.52 | 0.87 | 4.829E-01 | 397 | 121747 | 122144 |
| 295.1 | mental disorders | Schizophrenia | 1.14 | 0.13 | 0.19 | -0.25 | 0.48 | 4.836E-01 | 1157 | 120804 | 121961 |
| 317.11 | mental disorders | Alcoholic liver damage | 0.88 | -0.13 | 0.19 | -0.53 | 0.23 | 4.866E-01 | 1563 | 119990 | 121553 |
| 199.4 | neoplasms | Neurofibromatosis | 1.46 | 0.38 | 0.52 | -0.91 | 1.31 | 4.881E-01 | 120 | 122257 | 122377 |
| 516.1 | respiratory | Hemoptysis | 1.13 | 0.12 | 0.17 | -0.23 | 0.44 | 4.887E-01 | 1917 | 118610 | 120527 |
| 733.6 | musculoskeletal | Costochondritis | 1.40 | 0.33 | 0.47 | -0.79 | 1.19 | 4.906E-01 | 144 | 121715 | 121859 |
| 858 | injuries & poisonings | Complication of internal orthopedic device | 1.12 | 0.11 | 0.16 | -0.21 | 0.40 | 4.907E-01 | 1982 | 119057 | 121039 |
| 227.3 | neoplasms | Benign neoplasm of pituitary gland and craniopharyngeal duct (pouch) | 1.23 | 0.21 | 0.30 | -0.44 | 0.75 | 4.934E-01 | 511 | 121960 | 122471 |
| 187.2 | neoplasms | Malignant neoplasm of testis | 0.70 | -0.36 | 0.55 | -1.70 | 0.60 | 4.956E-01 | 217 | 119256 | 119473 |
| 288.11 | hematopoietic | Neutropenia | 1.15 | 0.14 | 0.20 | -0.27 | 0.50 | 4.956E-01 | 1273 | 120167 | 121440 |
| 794 | symptoms | Abnormal results of other function studies (bladder, | 1.27 | 0.24 | 0.34 | -0.52 | 0.86 | 4.958E-01 | 378 | 119506 | 119884 |

| | | | | | | | | | | | |
|---|---|---|---|---|---|---|---|---|---|---|---|
| | | pancreas, placenta, spleen, etc) | | | | | | | | | |
| 318 | mental disorders | Tobacco use disorder | 1.03 | 0.03 | 0.05 | -0.06 | 0.13 | 4.961E-01 | 36140 | 70858 | 106998 |
| 246.2 | endocrine/metabolic | Thyroid cyst | 0.61 | -0.49 | 0.78 | -2.66 | 0.80 | 4.970E-01 | 136 | 122219 | 122355 |
| 695.42 | dermatologic | Systemic lupus erythematosus | 0.71 | -0.34 | 0.53 | -1.63 | 0.59 | 4.997E-01 | 202 | 122266 | 122468 |
| 594.8 | genitourinary | Renal colic | 1.30 | 0.26 | 0.38 | -0.59 | 0.95 | 5.031E-01 | 259 | 121255 | 121514 |
| 441.1 | circulatory system | Acute vascular insufficiency of intestine | 1.34 | 0.29 | 0.42 | -0.69 | 1.05 | 5.039E-01 | 220 | 122016 | 122236 |
| 327.72 | neurological | Sleep related leg cramps | 1.23 | 0.20 | 0.30 | -0.44 | 0.74 | 5.046E-01 | 548 | 120665 | 121213 |
| 289.5 | hematopoietic | Diseases of spleen | 1.24 | 0.21 | 0.31 | -0.47 | 0.78 | 5.053E-01 | 453 | 121510 | 121963 |
| 715.2 | musculoskeletal | Ankylosing spondylitis | 1.27 | 0.24 | 0.34 | -0.53 | 0.86 | 5.058E-01 | 372 | 121983 | 122355 |
| 159.2 | neoplasms | Malignant neoplasm of small intestine, including duodenum | 0.68 | -0.38 | 0.62 | -1.95 | 0.66 | 5.065E-01 | 217 | 122243 | 122460 |
| 853 | injuries & poisonings | Complication of colostomy or enterostomy | 1.34 | 0.29 | 0.42 | -0.69 | 1.04 | 5.071E-01 | 240 | 122087 | 122327 |
| 246 | endocrine/metabolic | Other disorders of thyroid | 1.14 | 0.13 | 0.20 | -0.28 | 0.50 | 5.097E-01 | 1167 | 119448 | 120615 |
| 860 | neoplasms | Bone marrow or stem cell transplant | 1.43 | 0.36 | 0.52 | -0.93 | 1.29 | 5.107E-01 | 121 | 122401 | 122522 |
| 333.4 | neurological | Torsion dystonia | 1.19 | 0.18 | 0.26 | -0.39 | 0.66 | 5.132E-01 | 612 | 121333 | 121945 |
| 712 | musculoskeletal | Infective connective tissue disorders | 0.67 | -0.40 | 0.66 | -2.11 | 0.75 | 5.144E-01 | 142 | 122244 | 122386 |
| 562.2 | digestive | Diverticulitis | 1.08 | 0.08 | 0.12 | -0.17 | 0.31 | 5.148E-01 | 3591 | 114771 | 118362 |
| 38.1 | infectious diseases | Gram negative septicemia | 1.17 | 0.16 | 0.24 | -0.34 | 0.59 | 5.153E-01 | 925 | 120099 | 121024 |
| 70.2 | infectious diseases | Viral hepatitis B | 0.80 | -0.22 | 0.35 | -0.99 | 0.41 | 5.158E-01 | 465 | 121692 | 122157 |
| 324 | neurological | Other CNS infection and poliomyelitis | 1.28 | 0.25 | 0.37 | -0.58 | 0.90 | 5.166E-01 | 327 | 121951 | 122278 |
| 797 | symptoms | Shock | 1.19 | 0.18 | 0.27 | -0.40 | 0.66 | 5.194E-01 | 695 | 120662 | 121357 |
| 577.1 | digestive | Acute pancreatitis | 1.11 | 0.10 | 0.16 | -0.22 | 0.40 | 5.208E-01 | 2019 | 119444 | 121463 |
| 389.1 | sense organs | Sensorineural hearing loss | 1.03 | 0.03 | 0.05 | -0.06 | 0.13 | 5.209E-01 | 72799 | 37267 | 110066 |
| 969 | injuries & poisonings | Poisoning by psychotropic agents | 1.29 | 0.26 | 0.39 | -0.63 | 0.97 | 5.218E-01 | 218 | 121671 | 121889 |
| 157 | neoplasms | Pancreatic cancer | 1.20 | 0.19 | 0.28 | -0.43 | 0.70 | 5.229E-01 | 647 | 121603 | 122250 |

| | | | | | | | | | | | |
|---|---|---|---|---|---|---|---|---|---|---|---|
| 271.9 | endocrine/metabolic | Other disorders of carbohydrate transport and metabolism | 1.41 | 0.34 | 0.52 | -0.94 | 1.26 | 5.234E-01 | 151 | 122242 | 122393 |
| 444.2 | circulatory system | Embolism and thrombosis of abdominal aorta | 1.42 | 0.35 | 0.53 | -0.95 | 1.29 | 5.266E-01 | 125 | 122142 | 122267 |
| 743.21 | musculoskeletal | Pathologic fracture of vertebrae | 1.22 | 0.20 | 0.31 | -0.48 | 0.76 | 5.296E-01 | 496 | 120935 | 121431 |
| 603.1 | genitourinary | Hydrocele | 1.11 | 0.10 | 0.16 | -0.24 | 0.41 | 5.365E-01 | 2119 | 115681 | 117800 |
| 262 | endocrine/metabolic | Mineral deficiency NEC | 1.18 | 0.16 | 0.26 | -0.39 | 0.64 | 5.373E-01 | 715 | 120574 | 121289 |
| 291.8 | mental disorders | Alteration of consciousness | 1.17 | 0.16 | 0.25 | -0.38 | 0.62 | 5.393E-01 | 758 | 119089 | 119847 |
| 170.2 | neoplasms | Cancer of connective tissue | 1.23 | 0.20 | 0.32 | -0.51 | 0.79 | 5.396E-01 | 469 | 121565 | 122034 |
| 381.3 | sense organs | Mastoiditis & related conditions | 1.23 | 0.20 | 0.33 | -0.52 | 0.79 | 5.399E-01 | 452 | 121565 | 122017 |
| 818 | injuries & poisonings | Intracranial hemorrhage (injury) | 0.85 | -0.16 | 0.26 | -0.73 | 0.33 | 5.415E-01 | 971 | 121022 | 121993 |
| 355 | neurological | Complex regional/central pain syndrome | 1.21 | 0.19 | 0.31 | -0.48 | 0.76 | 5.424E-01 | 400 | 121238 | 121638 |
| 519.9 | respiratory | Symptoms involving respiratory system and other chest symptoms | 1.10 | 0.10 | 0.16 | -0.24 | 0.40 | 5.473E-01 | 2054 | 116725 | 118779 |
| 498 | respiratory | Acute bronchospasm | 1.33 | 0.29 | 0.46 | -0.82 | 1.11 | 5.489E-01 | 191 | 121212 | 121403 |
| 604.3 | genitourinary | Peyronie's disease | 0.83 | -0.19 | 0.33 | -0.91 | 0.40 | 5.516E-01 | 639 | 118590 | 119229 |
| 287.32 | hematopoietic | Secondary thrombocytopenia | 1.17 | 0.16 | 0.26 | -0.40 | 0.63 | 5.524E-01 | 738 | 120649 | 121387 |
| 520.2 | digestive | Disturbances in tooth eruption | 1.33 | 0.28 | 0.47 | -0.83 | 1.14 | 5.543E-01 | 148 | 121074 | 121222 |
| 174.2 | neoplasms | Breast cancer [male] | 1.44 | 0.36 | 0.61 | -1.20 | 1.42 | 5.545E-01 | 110 | 119504 | 119614 |
| 316.1 | mental disorders | Polyneuropathy due to drugs | 0.66 | -0.42 | 0.78 | -2.59 | 0.86 | 5.563E-01 | 135 | 122216 | 122351 |
| 946 | injuries & poisonings | Anaphylactic shock NOS | 0.81 | -0.21 | 0.37 | -1.03 | 0.45 | 5.571E-01 | 493 | 121396 | 121889 |
| 750.2 | congenital anomalies | Lower gastrointestinal congenital anomalies | 0.77 | -0.26 | 0.47 | -1.36 | 0.56 | 5.590E-01 | 324 | 121114 | 121438 |
| 513.32 | respiratory | Orthopnea | 1.29 | 0.25 | 0.42 | -0.73 | 1.00 | 5.613E-01 | 252 | 121414 | 121666 |
| 223 | neoplasms | Benign neoplasm of kidney and other urinary organs | 1.19 | 0.17 | 0.30 | -0.47 | 0.71 | 5.633E-01 | 590 | 120715 | 121305 |
| 297.2 | mental disorders | Suicide or self-inflicted injury | 1.19 | 0.17 | 0.30 | -0.47 | 0.72 | 5.655E-01 | 429 | 121622 | 122051 |
| 990 | injuries & poisonings | Effects radiation NOS | 0.91 | -0.09 | 0.16 | -0.42 | 0.21 | 5.698E-01 | 2595 | 117225 | 119820 |

| | | | | | | | | | | | |
|---|---|---|---|---|---|---|---|---|---|---|---|
| 727.2 | musculoskeletal | Bursitis disorders | 1.10 | 0.09 | 0.16 | -0.24 | 0.39 | 5.712E-01 | 1990 | 116566 | 118556 |
| 585.4 | genitourinary | Chronic kidney disease, Stage I or II | 1.08 | 0.07 | 0.13 | -0.19 | 0.32 | 5.781E-01 | 3412 | 115464 | 118876 |
| 751.22 | congenital anomalies | Other specified congenital anomalies of kidney | 1.34 | 0.29 | 0.52 | -0.99 | 1.21 | 5.798E-01 | 149 | 122208 | 122357 |
| 334.21 | neurological | Amyotrophic Lateral Sclerosis | 0.73 | -0.32 | 0.62 | -1.88 | 0.73 | 5.811E-01 | 194 | 122297 | 122491 |
| 994.1 | injuries & poisonings | Systemic inflammatory response syndrome (SIRS) | 1.23 | 0.20 | 0.37 | -0.62 | 0.86 | 5.830E-01 | 336 | 121068 | 121404 |
| 292.4 | mental disorders | Altered mental status | 1.06 | 0.06 | 0.11 | -0.16 | 0.27 | 5.874E-01 | 4949 | 111669 | 116618 |
| 155.1 | neoplasms | Malignant neoplasm of liver, primary | 0.86 | -0.15 | 0.29 | -0.78 | 0.38 | 5.924E-01 | 700 | 121559 | 122259 |
| 574.11 | digestive | Cholelithiasis with acute cholecystitis | 0.83 | -0.19 | 0.36 | -1.01 | 0.46 | 5.930E-01 | 516 | 121160 | 121676 |
| 274.2 | endocrine/metabolic | Crystal arthropathies | 1.12 | 0.11 | 0.21 | -0.32 | 0.49 | 5.936E-01 | 1295 | 119729 | 121024 |
| 189.11 | neoplasms | Malignant neoplasm of kidney, except pelvis | 0.91 | -0.09 | 0.18 | -0.46 | 0.24 | 5.940E-01 | 2137 | 119751 | 121888 |
| 603 | genitourinary | Other disorders of testis | 1.08 | 0.08 | 0.15 | -0.23 | 0.36 | 5.944E-01 | 2658 | 114625 | 117283 |
| 170.1 | neoplasms | Bone cancer | 1.28 | 0.25 | 0.46 | -0.85 | 1.07 | 5.969E-01 | 208 | 121978 | 122186 |
| 459.1 | circulatory system | Hemorrhage NOS | 1.32 | 0.28 | 0.52 | -1.00 | 1.19 | 5.974E-01 | 162 | 121206 | 121368 |
| 749 | congenital anomalies | Congenital anomalies of face and neck | 1.31 | 0.27 | 0.52 | -1.01 | 1.19 | 5.997E-01 | 159 | 121822 | 121981 |
| 374.1 | sense organs | Ectropion or entropion | 0.94 | -0.06 | 0.12 | -0.32 | 0.17 | 6.006E-01 | 4822 | 114036 | 118858 |
| 601.4 | genitourinary | Balanoposthitis | 1.14 | 0.13 | 0.25 | -0.40 | 0.59 | 6.016E-01 | 933 | 117399 | 118332 |
| 733.8 | musculoskeletal | Malunion and nonunion of fracture | 1.17 | 0.15 | 0.29 | -0.48 | 0.70 | 6.029E-01 | 481 | 121383 | 121864 |
| 264 | endocrine/metabolic | Lack of normal physiological development | 0.70 | -0.36 | 0.78 | -2.53 | 0.93 | 6.040E-01 | 126 | 122098 | 122224 |
| 602.3 | genitourinary | Dysplasia of prostate | 0.81 | -0.21 | 0.42 | -1.18 | 0.54 | 6.060E-01 | 434 | 118453 | 118887 |
| 528.3 | digestive | Cellulitis and abscess of oral soft tissues | 1.25 | 0.23 | 0.44 | -0.79 | 1.01 | 6.061E-01 | 202 | 121395 | 121597 |
| 535.8 | digestive | Other specified gastritis | 1.14 | 0.13 | 0.26 | -0.42 | 0.60 | 6.075E-01 | 769 | 118601 | 119370 |
| 599.6 | genitourinary | Oliguria and anuria | 1.34 | 0.29 | 0.60 | -1.28 | 1.35 | 6.083E-01 | 106 | 121970 | 122076 |
| 526.9 | digestive | Jaw disease NOS | 1.30 | 0.26 | 0.52 | -1.02 | 1.18 | 6.090E-01 | 144 | 121772 | 121916 |
| 750 | congenital anomalies | Digestive congenital anomalies | 1.15 | 0.14 | 0.27 | -0.43 | 0.62 | 6.097E-01 | 725 | 119585 | 120310 |

| | | | | | | | | | | | |
|---|---|---|---|---|---|---|---|---|---|---|---|
| 573.3 | digestive | Hepatomegaly | 0.86 | -0.16 | 0.31 | -0.84 | 0.42 | 6.098E-01 | 593 | 120306 | 120899 |
| 427.42 | circulatory system | Cardiac arrest | 1.16 | 0.15 | 0.29 | -0.47 | 0.67 | 6.100E-01 | 578 | 120995 | 121573 |
| 270.2 | endocrine/metabolic | Disorders of amino-acid metabolism | 0.87 | -0.14 | 0.29 | -0.77 | 0.38 | 6.108E-01 | 712 | 120725 | 121437 |
| 694.3 | dermatologic | Vascular disorders of skin | 0.70 | -0.35 | 0.78 | -2.53 | 0.93 | 6.115E-01 | 121 | 122184 | 122305 |
| 277.5 | endocrine/metabolic | Other disorders of lipoid metabolism | 1.09 | 0.09 | 0.17 | -0.26 | 0.40 | 6.138E-01 | 1725 | 118183 | 119908 |
| 560 | digestive | Intestinal obstruction without mention of hernia | 1.07 | 0.06 | 0.13 | -0.19 | 0.31 | 6.148E-01 | 3535 | 115214 | 118749 |
| 376 | sense organs | Disorders of the orbit | 1.33 | 0.28 | 0.61 | -1.28 | 1.34 | 6.173E-01 | 115 | 122156 | 122271 |
| 440.1 | circulatory system | Atherosclerosis of renal artery | 1.18 | 0.16 | 0.33 | -0.55 | 0.75 | 6.179E-01 | 496 | 121511 | 122007 |
| 290.12 | mental disorders | Dementia with cerebral degenerations | 0.87 | -0.14 | 0.28 | -0.75 | 0.38 | 6.194E-01 | 898 | 120946 | 121844 |
| 204.22 | neoplasms | Myeloid leukemia, chronic | 1.23 | 0.21 | 0.42 | -0.77 | 0.96 | 6.206E-01 | 251 | 122267 | 122518 |
| 803.1 | injuries & poisonings | Fracture of humerus | 0.91 | -0.10 | 0.20 | -0.53 | 0.28 | 6.207E-01 | 1348 | 120724 | 122072 |
| 244.2 | endocrine/metabolic | Acquired hypothyroidism | 1.09 | 0.09 | 0.18 | -0.28 | 0.42 | 6.213E-01 | 1556 | 119252 | 120808 |
| 348.9 | neurological | Other conditions of brain, NOS | 1.14 | 0.13 | 0.27 | -0.44 | 0.63 | 6.214E-01 | 655 | 119924 | 120579 |
| 695.22 | dermatologic | Pemphigus and pemphigoid | 1.25 | 0.23 | 0.46 | -0.87 | 1.04 | 6.220E-01 | 231 | 122228 | 122459 |
| 465.4 | respiratory | Acute laryngitis and tracheitis | 1.26 | 0.23 | 0.47 | -0.89 | 1.07 | 6.223E-01 | 162 | 121418 | 121580 |
| 335 | neurological | Multiple sclerosis | 1.15 | 0.14 | 0.29 | -0.49 | 0.68 | 6.281E-01 | 441 | 121890 | 122331 |
| 603.2 | genitourinary | Spermatocele | 0.86 | -0.15 | 0.33 | -0.87 | 0.43 | 6.301E-01 | 648 | 118034 | 118682 |
| 854 | injuries & poisonings | Complications of cardiac/vascular device, implant, and graft | 1.08 | 0.08 | 0.16 | -0.25 | 0.37 | 6.302E-01 | 2232 | 117482 | 119714 |
| 747.13 | congenital anomalies | Congenital anomalies of great vessels | 1.27 | 0.24 | 0.52 | -1.04 | 1.16 | 6.303E-01 | 149 | 122063 | 122212 |
| 264.2 | endocrine/metabolic | Failure to thrive (childhood) | 0.73 | -0.32 | 0.78 | -2.49 | 0.97 | 6.338E-01 | 124 | 122107 | 122231 |
| 747.11 | congenital anomalies | Cardiac shunt/ heart septal defect | 1.19 | 0.17 | 0.37 | -0.65 | 0.83 | 6.350E-01 | 346 | 121789 | 122135 |
| 348.2 | neurological | Cerebral edema and compression of brain | 1.23 | 0.21 | 0.45 | -0.85 | 1.03 | 6.358E-01 | 174 | 121965 | 122139 |
| 871.1 | injuries & poisonings | Open wound of hand except finger(s) | 1.13 | 0.12 | 0.26 | -0.44 | 0.61 | 6.368E-01 | 709 | 119391 | 120100 |

| | | | | | | | | | | | |
|---|---|---|---|---|---|---|---|---|---|---|---|
| 346 | neurological | Abnormal findings on study of brain and/or nervous system | 1.24 | 0.21 | 0.47 | -0.89 | 1.04 | 6.370E-01 | 190 | 121190 | 121380 |
| 759 | congenital anomalies | Other and unspecified congenital anomalies | 1.27 | 0.24 | 0.53 | -1.07 | 1.19 | 6.382E-01 | 124 | 122114 | 122238 |
| 149.5 | neoplasms | Hx of malignant neoplasm of oral cavity and pharynx | 1.20 | 0.18 | 0.39 | -0.71 | 0.89 | 6.392E-01 | 299 | 122009 | 122308 |
| 425.8 | circulatory system | Other cardiomyopathy | 1.24 | 0.22 | 0.48 | -0.91 | 1.07 | 6.396E-01 | 154 | 122246 | 122400 |
| 427.11 | circulatory system | Paroxysmal supraventricular tachycardia | 1.07 | 0.07 | 0.14 | -0.23 | 0.34 | 6.401E-01 | 2578 | 117064 | 119642 |
| 741.2 | musculoskeletal | Stiffness of joint | 0.91 | -0.10 | 0.21 | -0.55 | 0.30 | 6.415E-01 | 1179 | 119206 | 120385 |
| 614.5 | genitourinary | Inflammatory disease of cervix, vagina, and vulva | 0.87 | -0.13 | 0.31 | -0.79 | 0.44 | 6.452E-01 | 215 | 2368 | 2583 |
| 291.1 | mental disorders | Transient mental disorders due to conditions classified elsewhere | 1.29 | 0.26 | 0.61 | -1.32 | 1.33 | 6.464E-01 | 101 | 122153 | 122254 |
| 253.3 | endocrine/metabolic | Diabetes insipidus | 0.74 | -0.30 | 0.78 | -2.47 | 0.99 | 6.471E-01 | 113 | 122325 | 122438 |
| 252.2 | endocrine/metabolic | Hypoparathyroidism | 1.25 | 0.22 | 0.53 | -1.07 | 1.16 | 6.505E-01 | 128 | 122301 | 122429 |
| 442.2 | circulatory system | Aneurysm of iliac artery | 0.89 | -0.12 | 0.27 | -0.71 | 0.38 | 6.528E-01 | 954 | 120890 | 121844 |
| 312.3 | mental disorders | Impulse control disorder | 1.13 | 0.12 | 0.27 | -0.45 | 0.61 | 6.535E-01 | 609 | 121666 | 122275 |
| 994.21 | injuries & poisonings | Septic shock | 0.89 | -0.12 | 0.27 | -0.71 | 0.38 | 6.562E-01 | 862 | 120490 | 121352 |
| 741.1 | musculoskeletal | Ankylosis of joint | 1.24 | 0.22 | 0.53 | -1.07 | 1.15 | 6.610E-01 | 133 | 122095 | 122228 |
| 747 | congenital anomalies | Cardiac and circulatory congenital anomalies | 1.09 | 0.08 | 0.19 | -0.31 | 0.44 | 6.611E-01 | 1482 | 118402 | 119884 |
| 656 | pregnancy complications | Other perinatal conditions of fetus or newborn | 0.75 | -0.29 | 0.78 | -2.47 | 0.99 | 6.614E-01 | 115 | 122065 | 122180 |
| 473.3 | respiratory | Paralysis/spasm of vocal cords or larynx | 0.86 | -0.15 | 0.37 | -0.97 | 0.50 | 6.624E-01 | 495 | 121677 | 122172 |
| 501 | respiratory | Pneumonitis due to inhalation of food or vomitus | 0.91 | -0.09 | 0.22 | -0.57 | 0.32 | 6.678E-01 | 1370 | 118841 | 120211 |
| 151 | neoplasms | Cancer of stomach | 1.16 | 0.15 | 0.34 | -0.62 | 0.76 | 6.688E-01 | 437 | 121843 | 122280 |
| 952 | injuries & poisonings | Spinal cord injury without evidence of spinal bone injury | 1.15 | 0.14 | 0.33 | -0.60 | 0.75 | 6.708E-01 | 388 | 121748 | 122136 |
| 411.9 | circulatory system | Other acute and subacute forms of ischemic heart disease | 1.14 | 0.13 | 0.31 | -0.55 | 0.69 | 6.717E-01 | 535 | 120056 | 120591 |

| | | | | | | | | | | | |
|---|---|---|---|---|---|---|---|---|---|---|---|
| 275.1 | hematopoietic | Disorders of iron metabolism | 0.89 | -0.11 | 0.28 | -0.71 | 0.39 | 6.723E-01 | 768 | 121198 | 121966 |
| 448 | circulatory system | Disease of capillaries | 0.82 | -0.20 | 0.53 | -1.48 | 0.71 | 6.735E-01 | 241 | 121453 | 121694 |
| 567 | digestive | Peritonitis and retroperitoneal infections | 1.10 | 0.09 | 0.23 | -0.38 | 0.51 | 6.752E-01 | 969 | 120270 | 121239 |
| 444.1 | circulatory system | Arterial embolism and thrombosis of lower extremity artery | 0.89 | -0.12 | 0.31 | -0.80 | 0.44 | 6.839E-01 | 681 | 120926 | 121607 |
| 415.11 | circulatory system | Pulmonary embolism and infarction, acute | 1.05 | 0.05 | 0.12 | -0.20 | 0.28 | 6.849E-01 | 3641 | 117794 | 121435 |
| 300.8 | mental disorders | Acute reaction to stress | 1.11 | 0.10 | 0.26 | -0.44 | 0.58 | 6.860E-01 | 625 | 120267 | 120892 |
| 733.4 | musculoskeletal | Aseptic necrosis of bone | 0.89 | -0.11 | 0.29 | -0.75 | 0.42 | 6.865E-01 | 625 | 121386 | 122011 |
| 170 | neoplasms | Cancer of bone and connective tissue | 1.12 | 0.11 | 0.28 | -0.50 | 0.63 | 6.872E-01 | 665 | 121024 | 121689 |
| 575.9 | digestive | Nonspecific abnormal findings on radiological and other examination of biliary tract | 0.90 | -0.11 | 0.28 | -0.71 | 0.40 | 6.893E-01 | 721 | 120190 | 120911 |
| 363.4 | sense organs | Choroidal degenerations | 0.79 | -0.24 | 0.78 | -2.41 | 1.05 | 6.902E-01 | 123 | 122067 | 122190 |
| 714 | musculoskeletal | Rheumatoid arthritis and other inflammatory polyarthropathies | 1.05 | 0.04 | 0.11 | -0.18 | 0.26 | 6.918E-01 | 4331 | 115892 | 120223 |
| 286.5 | hematopoietic | Hemorrhagic disorder due to intrinsic circulating anticoagulants | 1.17 | 0.16 | 0.42 | -0.82 | 0.90 | 6.977E-01 | 302 | 121927 | 122229 |
| 198.1 | neoplasms | Secondary malignancy of lymph nodes | 1.06 | 0.06 | 0.15 | -0.25 | 0.34 | 6.983E-01 | 2467 | 118867 | 121334 |
| 425.2 | circulatory system | Secondary/extrinsic cardiomyopathies | 0.88 | -0.13 | 0.34 | -0.89 | 0.49 | 6.985E-01 | 575 | 120810 | 121385 |
| 695.41 | dermatologic | Cutaneous lupus erythematosus | 0.83 | -0.18 | 0.53 | -1.47 | 0.76 | 7.007E-01 | 181 | 122325 | 122506 |
| 782.6 | symptoms | Pallor and flushing | 0.83 | -0.18 | 0.53 | -1.48 | 0.75 | 7.008E-01 | 202 | 121603 | 121805 |
| 604.1 | genitourinary | Redundant prepuce and phimosis/BXO | 1.08 | 0.08 | 0.20 | -0.34 | 0.45 | 7.015E-01 | 1533 | 117320 | 118853 |
| 292.12 | mental disorders | Symbolic dysfunction | 0.82 | -0.20 | 0.61 | -1.77 | 0.86 | 7.029E-01 | 157 | 122154 | 122311 |
| 415.1 | circulatory system | Acute pulmonary heart disease | 1.05 | 0.05 | 0.12 | -0.20 | 0.28 | 7.062E-01 | 3696 | 117647 | 121343 |

| | | | | | | | | | | | |
|---|---|---|---|---|---|---|---|---|---|---|---|
| 189.1 | neoplasms | Cancer of kidney and renal pelvis | 0.94 | -0.06 | 0.17 | -0.41 | 0.25 | 7.072E-01 | 2348 | 119462 | 121810 |
| 756.5 | congenital anomalies | Congenital osteodystrophies | 1.17 | 0.16 | 0.47 | -0.96 | 1.00 | 7.075E-01 | 166 | 121730 | 121896 |
| 260 | endocrine/metabolic | Protein-calorie malnutrition | 1.04 | 0.04 | 0.11 | -0.17 | 0.24 | 7.084E-01 | 5445 | 111793 | 117238 |
| 578.1 | digestive | Hematemesis | 1.12 | 0.11 | 0.31 | -0.57 | 0.68 | 7.094E-01 | 501 | 121217 | 121718 |
| 255.12 | endocrine/metabolic | Hyperaldosteronism | 1.18 | 0.16 | 0.46 | -0.94 | 0.98 | 7.108E-01 | 231 | 122241 | 122472 |
| 332 | neurological | Parkinson's disease | 1.05 | 0.05 | 0.13 | -0.22 | 0.29 | 7.122E-01 | 3697 | 117934 | 121631 |
| 386.3 | sense organs | Labyrinthitis | 1.17 | 0.16 | 0.47 | -0.94 | 0.98 | 7.123E-01 | 228 | 121403 | 121631 |
| 274.11 | endocrine/metabolic | Gouty arthropathy | 1.05 | 0.05 | 0.14 | -0.24 | 0.32 | 7.149E-01 | 2959 | 116626 | 119585 |
| 803.21 | injuries & poisonings | Colles' fracture | 1.16 | 0.15 | 0.47 | -0.96 | 0.99 | 7.162E-01 | 167 | 122251 | 122418 |
| 797.1 | symptoms | Cardiogenic shock | 1.14 | 0.13 | 0.39 | -0.76 | 0.83 | 7.169E-01 | 332 | 121657 | 121989 |
| 585.31 | genitourinary | Renal dialysis | 1.09 | 0.08 | 0.23 | -0.40 | 0.50 | 7.191E-01 | 1075 | 121115 | 122190 |
| 480.12 | respiratory | Pseudomonal pneumonia | 1.19 | 0.18 | 0.61 | -1.39 | 1.23 | 7.204E-01 | 127 | 122230 | 122357 |
| 293 | mental disorders | Symptoms involving head and neck | 1.19 | 0.17 | 0.61 | -1.40 | 1.23 | 7.214E-01 | 119 | 121457 | 121576 |
| 697 | dermatologic | Sarcoidosis | 1.13 | 0.12 | 0.36 | -0.67 | 0.77 | 7.217E-01 | 320 | 122144 | 122464 |
| 331.9 | neurological | Cerebral degeneration, unspecified | 1.13 | 0.12 | 0.36 | -0.70 | 0.77 | 7.266E-01 | 414 | 120139 | 120553 |
| 191.1 | neoplasms | Cancer of brain and nervous system | 0.88 | -0.13 | 0.39 | -1.02 | 0.57 | 7.275E-01 | 413 | 121719 | 122132 |
| 159.3 | neoplasms | Malignant neoplasm of gallbladder and extrahepatic bile ducts | 1.18 | 0.16 | 0.52 | -1.11 | 1.07 | 7.281E-01 | 180 | 122310 | 122490 |
| 724.1 | musculoskeletal | Disorders of sacrum | 0.91 | -0.09 | 0.28 | -0.69 | 0.42 | 7.305E-01 | 668 | 120784 | 121452 |
| 560.1 | digestive | Paralytic ileus | 0.92 | -0.09 | 0.26 | -0.66 | 0.40 | 7.307E-01 | 914 | 119611 | 120525 |
| 433.5 | circulatory system | Cerebral aneurysm | 1.10 | 0.10 | 0.30 | -0.55 | 0.64 | 7.342E-01 | 519 | 121782 | 122301 |
| 572 | digestive | Ascites (non malignant) | 1.08 | 0.07 | 0.22 | -0.39 | 0.48 | 7.344E-01 | 1074 | 120311 | 121385 |
| 556 | digestive | Ulceration of the lower GI tract | 1.11 | 0.10 | 0.31 | -0.58 | 0.66 | 7.346E-01 | 565 | 120374 | 120939 |
| 204.2 | neoplasms | Myeloid leukemia | 0.89 | -0.12 | 0.36 | -0.94 | 0.54 | 7.356E-01 | 486 | 121928 | 122414 |
| 204.4 | neoplasms | Multiple myeloma | 0.90 | -0.10 | 0.32 | -0.82 | 0.48 | 7.360E-01 | 657 | 121666 | 122323 |
| 286.1 | hematopoietic | Congenital coagulation defects | 0.87 | -0.14 | 0.47 | -1.25 | 0.71 | 7.416E-01 | 227 | 122154 | 122381 |

| | | | | | | | | | | | |
|---|---|---|---|---|---|---|---|---|---|---|---|
| 550.5 | digestive | Ventral hernia | 0.95 | -0.05 | 0.15 | -0.35 | 0.23 | 7.417E-01 | 2951 | 116813 | 119764 |
| 305.2 | mental disorders | Eating disorder | 0.89 | -0.12 | 0.44 | -1.15 | 0.70 | 7.418E-01 | 167 | 122225 | 122392 |
| 302.1 | mental disorders | Decreased libido | 1.11 | 0.10 | 0.33 | -0.63 | 0.71 | 7.423E-01 | 406 | 121207 | 121613 |
| 165.1 | neoplasms | Cancer of bronchus; lung | 0.96 | -0.04 | 0.12 | -0.27 | 0.18 | 7.467E-01 | 4899 | 116330 | 121229 |
| 253.1 | endocrine/metabolic | Pituitary hyperfunction | 1.15 | 0.14 | 0.52 | -1.14 | 1.05 | 7.513E-01 | 169 | 122285 | 122454 |
| 580.2 | genitourinary | Nephrotic syndrome without mention of glomerulonephritis | 0.88 | -0.13 | 0.47 | -1.23 | 0.69 | 7.542E-01 | 282 | 121899 | 122181 |
| 155 | neoplasms | Cancer of liver and intrahepatic bile duct | 1.08 | 0.07 | 0.24 | -0.44 | 0.51 | 7.552E-01 | 875 | 121287 | 122162 |
| 145.2 | neoplasms | Cancer of tongue | 1.09 | 0.09 | 0.29 | -0.53 | 0.61 | 7.555E-01 | 654 | 121653 | 122307 |
| 361.2 | sense organs | Retinoschisis and retinal cysts | 1.09 | 0.09 | 0.28 | -0.53 | 0.60 | 7.570E-01 | 660 | 121526 | 122186 |
| 702.4 | dermatologic | Degenerative skin disorders | 0.91 | -0.10 | 0.78 | -2.27 | 1.19 | 7.577E-01 | 107 | 122203 | 122310 |
| 696 | dermatologic | Psoriasis and related disorders | 1.03 | 0.03 | 0.11 | -0.19 | 0.24 | 7.587E-01 | 4983 | 115188 | 120171 |
| 428.2 | circulatory system | Heart failure NOS | 1.05 | 0.05 | 0.18 | -0.31 | 0.38 | 7.593E-01 | 1946 | 117596 | 119542 |
| 189.4 | neoplasms | Malignant neoplasm of other urinary organs | 0.89 | -0.12 | 0.42 | -1.09 | 0.63 | 7.616E-01 | 395 | 121773 | 122168 |
| 270.33 | endocrine/metabolic | Amyloidosis | 1.14 | 0.13 | 0.52 | -1.14 | 1.04 | 7.651E-01 | 205 | 122247 | 122452 |
| 573.5 | digestive | Jaundice (not of newborn) | 0.92 | -0.09 | 0.31 | -0.76 | 0.47 | 7.693E-01 | 669 | 120874 | 121543 |
| 117.1 | infectious diseases | Histoplasmosis | 1.10 | 0.10 | 0.37 | -0.72 | 0.75 | 7.709E-01 | 408 | 121938 | 122346 |
| 386.21 | sense organs | Central origin vertigo | 1.11 | 0.10 | 0.43 | -0.89 | 0.86 | 7.748E-01 | 252 | 121482 | 121734 |
| 242.2 | endocrine/metabolic | Toxic multinodular goiter | 1.12 | 0.11 | 0.61 | -1.46 | 1.17 | 7.748E-01 | 123 | 122328 | 122451 |
| 442.3 | circulatory system | Aneurysm of artery of lower extremity | 1.09 | 0.09 | 0.32 | -0.63 | 0.67 | 7.749E-01 | 523 | 121585 | 122108 |
| 201 | neoplasms | Hodgkin's disease | 0.90 | -0.10 | 0.44 | -1.11 | 0.67 | 7.780E-01 | 283 | 122030 | 122313 |
| 159 | neoplasms | Malignant neoplasm of other and ill-defined sites within the digestive organs and peritoneum | 0.92 | -0.08 | 0.30 | -0.72 | 0.46 | 7.788E-01 | 717 | 121161 | 121878 |
| 110.2 | infectious diseases | Dermatomycoses | 0.92 | -0.09 | 0.33 | -0.81 | 0.51 | 7.790E-01 | 544 | 120335 | 120879 |
| 187.1 | neoplasms | Malignant neoplasm of unspecified male genital organ | 1.11 | 0.11 | 0.52 | -1.17 | 1.02 | 7.799E-01 | 190 | 119095 | 119285 |

| | | | | | | | | | | | |
|---|---|---|---|---|---|---|---|---|---|---|---|
| 580.12 | genitourinary | Non-proliferative glomerulonephritis | 0.88 | -0.12 | 0.61 | -1.69 | 0.93 | 7.817E-01 | 154 | 122310 | 122464 |
| 287.4 | hematopoietic | Qualitative platelet defects | 1.10 | 0.09 | 0.61 | -1.48 | 1.15 | 7.825E-01 | 129 | 122208 | 122337 |
| 535.1 | digestive | Acute gastritis | 0.92 | -0.09 | 0.37 | -0.92 | 0.57 | 7.851E-01 | 425 | 120138 | 120563 |
| 962.3 | injuries & poisonings | Hormones and synthetic substitutes causing adverse effects in therapeutic use | 1.11 | 0.10 | 0.52 | -1.18 | 1.03 | 7.861E-01 | 164 | 121518 | 121682 |
| 550.3 | digestive | Femoral hernia | 1.03 | 0.03 | 0.61 | -1.54 | 1.09 | 7.874E-01 | 116 | 122258 | 122374 |
| 706.3 | dermatologic | Seborrhea | 0.91 | -0.09 | 0.42 | -1.07 | 0.65 | 7.964E-01 | 371 | 121067 | 121438 |
| 977 | injuries & poisonings | Personal history of allergy to medicinal agents | 1.06 | 0.06 | 0.61 | -1.51 | 1.11 | 7.966E-01 | 132 | 121790 | 121922 |
| 747.1 | congenital anomalies | Cardiac congenital anomalies | 0.94 | -0.06 | 0.24 | -0.56 | 0.38 | 7.967E-01 | 1068 | 119507 | 120575 |
| 696.41 | dermatologic | Psoriasis vulgaris | 0.97 | -0.03 | 0.12 | -0.26 | 0.19 | 7.988E-01 | 4670 | 115774 | 120444 |
| 451.2 | circulatory system | Phlebitis and thrombophlebitis of lower extremities | 0.93 | -0.07 | 0.31 | -0.75 | 0.49 | 8.001E-01 | 648 | 120753 | 121401 |
| 198.3 | neoplasms | Secondary malignant neoplasm of digestive systems | 1.09 | 0.09 | 0.42 | -0.89 | 0.84 | 8.014E-01 | 290 | 121900 | 122190 |
| 504 | respiratory | Other alveolar and parietoalveolar pneumonopathy | 1.04 | 0.04 | 0.18 | -0.33 | 0.38 | 8.015E-01 | 1878 | 119280 | 121158 |
| 259 | endocrine/metabolic | Other endocrine disorders | 0.91 | -0.10 | 0.47 | -1.20 | 0.72 | 8.018E-01 | 273 | 121378 | 121651 |
| 255.1 | endocrine/metabolic | Adrenal hyperfunction | 0.91 | -0.09 | 0.47 | -1.19 | 0.72 | 8.021E-01 | 289 | 122099 | 122388 |
| 772.6 | symptoms | Facial weakness | 0.92 | -0.09 | 0.43 | -1.07 | 0.67 | 8.040E-01 | 306 | 121191 | 121497 |
| 289.8 | hematopoietic | Polycythemia vera, secondary | 0.95 | -0.06 | 0.24 | -0.56 | 0.38 | 8.042E-01 | 985 | 120845 | 121830 |
| 587 | genitourinary | Kidney replaced by transpant | 0.91 | -0.09 | 0.47 | -1.19 | 0.73 | 8.047E-01 | 256 | 122227 | 122483 |
| 394.3 | circulatory system | Aortic valve disease | 1.05 | 0.05 | 0.61 | -1.52 | 1.10 | 8.052E-01 | 144 | 122083 | 122227 |
| 480.11 | respiratory | Pneumococcal pneumonia | 0.94 | -0.06 | 0.27 | -0.66 | 0.44 | 8.059E-01 | 805 | 119698 | 120503 |
| 721.8 | musculoskeletal | Other allied disorders of spine | 1.08 | 0.08 | 0.37 | -0.75 | 0.74 | 8.065E-01 | 376 | 120418 | 120794 |
| 741.5 | musculoskeletal | Hemarthrosis | 1.06 | 0.06 | 0.61 | -1.51 | 1.11 | 8.075E-01 | 133 | 122156 | 122289 |
| 204.21 | neoplasms | Myeloid leukemia, acute | 0.91 | -0.09 | 0.53 | -1.37 | 0.82 | 8.096E-01 | 223 | 122258 | 122481 |
| 323 | neurological | Encephalitis | 0.92 | -0.08 | 0.61 | -1.65 | 0.97 | 8.101E-01 | 149 | 122220 | 122369 |

| | | | | | | | | | | | |
|---|---|---|---|---|---|---|---|---|---|---|---|
| 506 | respiratory | Empyema and pneumothorax | 0.95 | -0.05 | 0.21 | -0.50 | 0.35 | 8.141E-01 | 1338 | 119817 | 121155 |
| 383 | sense organs | Otosclerosis | 1.08 | 0.08 | 0.52 | -1.20 | 0.99 | 8.146E-01 | 176 | 122202 | 122378 |
| 333.8 | neurological | Other degenerative diseases of the basal ganglia | 0.99 | -0.01 | 0.61 | -1.58 | 1.03 | 8.169E-01 | 161 | 122136 | 122297 |
| 592.13 | genitourinary | Chronic interstitial cystitis | 0.99 | -0.01 | 0.54 | -1.32 | 0.95 | 8.184E-01 | 125 | 122281 | 122406 |
| 780 | symptoms | Hypothermia/Chills | 0.92 | -0.08 | 0.61 | -1.65 | 0.97 | 8.188E-01 | 153 | 121387 | 121540 |
| 527.8 | digestive | Other specified diseases of the salivary glands | 1.02 | 0.02 | 0.62 | -1.59 | 1.11 | 8.195E-01 | 123 | 122093 | 122216 |
| 425.12 | circulatory system | Other hypertrophic cardiomyopathy | 1.07 | 0.07 | 0.52 | -1.17 | 0.99 | 8.203E-01 | 152 | 122219 | 122371 |
| 368.91 | sense organs | Psychophysical visual disturbances | 0.92 | -0.08 | 0.47 | -1.18 | 0.73 | 8.205E-01 | 284 | 121654 | 121938 |
| 619.2 | genitourinary | Disorders of uterus, NEC | 1.05 | 0.04 | 0.32 | -0.64 | 0.64 | 8.211E-01 | 181 | 2584 | 2765 |
| 803.2 | injuries & poisonings | Fracture of radius and ulna | 0.96 | -0.04 | 0.16 | -0.37 | 0.27 | 8.218E-01 | 1866 | 120020 | 121886 |
| 274.21 | endocrine/metabolic | Chondrocalcinosis | 1.05 | 0.05 | 0.24 | -0.45 | 0.48 | 8.240E-01 | 1047 | 120183 | 121230 |
| 602 | genitourinary | Other disorders of prostate | 1.03 | 0.03 | 0.14 | -0.26 | 0.30 | 8.241E-01 | 3219 | 112093 | 115312 |
| 512.1 | respiratory | Wheezing | 1.05 | 0.04 | 0.21 | -0.40 | 0.43 | 8.246E-01 | 1245 | 117150 | 118395 |
| 574.12 | digestive | Cholelithiasis with other cholecystitis | 0.95 | -0.06 | 0.27 | -0.65 | 0.44 | 8.250E-01 | 792 | 120056 | 120848 |
| 473.1 | respiratory | Chronic laryngitis | 0.93 | -0.07 | 0.47 | -1.17 | 0.75 | 8.264E-01 | 285 | 121372 | 121657 |
| 300.13 | mental disorders | Phobia | 1.06 | 0.06 | 0.37 | -0.77 | 0.73 | 8.266E-01 | 350 | 121609 | 121959 |
| 536.7 | digestive | Complications of gastrostomy, colostomy and enterostomy | 0.93 | -0.07 | 0.43 | -1.05 | 0.69 | 8.269E-01 | 292 | 121952 | 122244 |
| 281.13 | hematopoietic | Folate-deficiency anemia | 1.07 | 0.07 | 0.43 | -0.93 | 0.83 | 8.295E-01 | 246 | 121944 | 122190 |
| 750.22 | congenital anomalies | Congenital anomaly of gallbladder, bile ducts, liver, pancreas | 1.06 | 0.06 | 0.47 | -1.04 | 0.88 | 8.301E-01 | 230 | 121859 | 122089 |
| 504.1 | respiratory | Idiopathic fibrosing alveolitis | 1.05 | 0.05 | 0.24 | -0.47 | 0.49 | 8.343E-01 | 1011 | 120724 | 121735 |
| 316 | mental disorders | Substance addiction and disorders | 0.98 | -0.02 | 0.09 | -0.20 | 0.15 | 8.364E-01 | 6271 | 111948 | 118219 |
| 601.8 | genitourinary | Other inflammatory disorders of male genital organs | 1.05 | 0.05 | 0.44 | -0.95 | 0.83 | 8.391E-01 | 257 | 118724 | 118981 |
| 145.3 | neoplasms | Cancer of major salivary glands | 1.06 | 0.06 | 0.39 | -0.83 | 0.76 | 8.399E-01 | 361 | 122005 | 122366 |

| | | | | | | | | | | | |
|---|---|---|---|---|---|---|---|---|---|---|---|
| 388 | sense organs | Other disorders of ear | 1.05 | 0.04 | 0.24 | -0.47 | 0.49 | 8.405E-01 | 1055 | 118571 | 119626 |
| 573.2 | digestive | Liver replaced by transplant | 0.95 | -0.05 | 0.48 | -1.18 | 0.81 | 8.406E-01 | 199 | 122338 | 122537 |
| 209 | neoplasms | Neuroendocrine tumors | 1.03 | 0.03 | 0.52 | -1.25 | 0.94 | 8.435E-01 | 188 | 122255 | 122443 |
| 750.11 | congenital anomalies | Esophageal atresia/tracheoesophageal fistula | 1.00 | 0.00 | 0.52 | -1.27 | 0.91 | 8.453E-01 | 202 | 121810 | 122012 |
| 796 | symptoms | Elevated prostate specific antigen [PSA] | 0.99 | -0.01 | 0.06 | -0.13 | 0.10 | 8.471E-01 | 22497 | 91093 | 113590 |
| 31 | infectious diseases | Diseases due to other mycobacteria | 0.95 | -0.05 | 0.42 | -1.03 | 0.70 | 8.472E-01 | 318 | 122061 | 122379 |
| 614 | genitourinary | Inflammatory diseases of female pelvic organs | 1.04 | 0.04 | 0.29 | -0.56 | 0.58 | 8.490E-01 | 229 | 2341 | 2570 |
| 575.1 | digestive | Cholangitis | 0.95 | -0.05 | 0.36 | -0.87 | 0.60 | 8.500E-01 | 477 | 121741 | 122218 |
| 573.6 | digestive | Nonspecific elevation of levels of transaminase or lactic acid dehydrogenase [LDH] | 0.96 | -0.04 | 0.21 | -0.48 | 0.36 | 8.527E-01 | 1189 | 118413 | 119602 |
| 275.11 | hematopoietic | Hereditary hemochromatosis | 0.98 | -0.02 | 0.43 | -1.00 | 0.74 | 8.548E-01 | 299 | 122221 | 122520 |
| 743.13 | musculoskeletal | Other specified osteoporosis | 0.98 | -0.02 | 0.47 | -1.13 | 0.80 | 8.554E-01 | 220 | 121993 | 122213 |
| 514.2 | respiratory | Solitary pulmonary nodule | 0.98 | -0.02 | 0.08 | -0.19 | 0.15 | 8.554E-01 | 8713 | 107438 | 116151 |
| 556.1 | digestive | Ulceration of intestine | 0.95 | -0.05 | 0.36 | -0.87 | 0.60 | 8.561E-01 | 465 | 120651 | 121116 |
| 290.13 | mental disorders | Senile dementia | 1.04 | 0.04 | 0.39 | -0.85 | 0.73 | 8.575E-01 | 402 | 121793 | 122195 |
| 913 | injuries & poisonings | Toxic effect of venom | 0.96 | -0.04 | 0.47 | -1.15 | 0.78 | 8.582E-01 | 243 | 121098 | 121341 |
| 800.3 | injuries & poisonings | Fracture of tibia and fibula | 0.97 | -0.03 | 0.21 | -0.47 | 0.36 | 8.593E-01 | 1160 | 120483 | 121643 |
| 320 | neurological | Meningitis | 1.03 | 0.03 | 0.47 | -1.07 | 0.85 | 8.603E-01 | 236 | 122067 | 122303 |
| 609 | genitourinary | Male infertility and abnormal spermatozoa | 1.05 | 0.04 | 0.33 | -0.68 | 0.63 | 8.618E-01 | 515 | 118385 | 118900 |
| 475.9 | respiratory | Postnasal drip | 0.96 | -0.05 | 0.33 | -0.76 | 0.54 | 8.620E-01 | 584 | 120002 | 120586 |
| 429.1 | circulatory system | Heart transplant/surgery | 1.04 | 0.04 | 0.24 | -0.48 | 0.48 | 8.659E-01 | 1037 | 119533 | 120570 |
| 414.2 | circulatory system | ASCVD | 0.97 | -0.03 | 0.19 | -0.43 | 0.33 | 8.673E-01 | 1804 | 119058 | 120862 |
| 292.6 | mental disorders | Hallucinations | 1.03 | 0.03 | 0.42 | -0.96 | 0.78 | 8.692E-01 | 282 | 121543 | 121825 |
| 252 | endocrine/metabolic | Disorders of parathyroid gland | 0.97 | -0.03 | 0.17 | -0.37 | 0.29 | 8.698E-01 | 2074 | 119281 | 121355 |
| 686.4 | dermatologic | Pyogenic granuloma | 1.01 | 0.01 | 0.47 | -1.09 | 0.82 | 8.700E-01 | 264 | 121537 | 121801 |

| | | | | | | | | | | | |
|---|---|---|---|---|---|---|---|---|---|---|---|
| 754.2 | congenital anomalies | Spondylolisthesis, congenital | 1.04 | 0.03 | 0.37 | -0.79 | 0.69 | 8.763E-01 | 373 | 121567 | 121940 |
| 798.1 | symptoms | Chronic fatigue syndrome | 1.03 | 0.03 | 0.23 | -0.45 | 0.45 | 8.766E-01 | 936 | 119314 | 120250 |
| 801.1 | injuries & poisonings | Fracture of foot | 1.03 | 0.03 | 0.19 | -0.36 | 0.38 | 8.770E-01 | 1397 | 120489 | 121886 |
| 149.1 | neoplasms | Cancer of oropharynx | 0.97 | -0.03 | 0.28 | -0.63 | 0.47 | 8.770E-01 | 704 | 121599 | 122303 |
| 530.2 | digestive | Esophageal bleeding (varices/hemorrhage) | 1.03 | 0.03 | 0.22 | -0.44 | 0.44 | 8.780E-01 | 979 | 120205 | 121184 |
| 514.1 | respiratory | Abnormal results of function study of pulmonary system | 0.99 | -0.01 | 0.47 | -1.11 | 0.81 | 8.794E-01 | 261 | 120995 | 121256 |
| 717 | musculoskeletal | Polymyalgia Rheumatica | 1.03 | 0.03 | 0.21 | -0.41 | 0.41 | 8.805E-01 | 1387 | 120855 | 122242 |
| 389.5 | sense organs | Disorders of acoustic nerve | 0.97 | -0.03 | 0.39 | -0.92 | 0.67 | 8.820E-01 | 360 | 121413 | 121773 |
| 198.5 | neoplasms | Secondary malignancy of brain/spine | 1.03 | 0.03 | 0.31 | -0.65 | 0.59 | 8.835E-01 | 567 | 121744 | 122311 |
| 348 | neurological | Other conditions of brain | 0.98 | -0.02 | 0.16 | -0.34 | 0.27 | 8.894E-01 | 2493 | 116799 | 119292 |
| 281.11 | hematopoietic | Pernicious anemia | 0.98 | -0.03 | 0.34 | -0.79 | 0.59 | 8.900E-01 | 494 | 121640 | 122134 |
| 374.2 | sense organs | Lagophthalmos | 0.98 | -0.02 | 0.36 | -0.84 | 0.63 | 8.904E-01 | 456 | 121389 | 121845 |
| 568 | digestive | Other disorders of peritoneum | 0.97 | -0.03 | 0.29 | -0.65 | 0.49 | 8.906E-01 | 709 | 120381 | 121090 |
| 212 | neoplasms | Benign neoplasm of respiratory and intrathoracic organs | 1.01 | 0.01 | 0.36 | -0.81 | 0.66 | 8.953E-01 | 423 | 121356 | 121779 |
| 433.12 | circulatory system | Cerebral atherosclerosis | 1.02 | 0.02 | 0.33 | -0.69 | 0.61 | 8.966E-01 | 573 | 120773 | 121346 |
| 599.8 | genitourinary | Other symptoms involving urinary system | 0.98 | -0.02 | 0.14 | -0.30 | 0.25 | 8.986E-01 | 3342 | 112859 | 116201 |
| 350.6 | neurological | Disturbances of sensation of smell and taste | 0.99 | -0.01 | 0.33 | -0.73 | 0.58 | 9.091E-01 | 522 | 121343 | 121865 |
| 571.51 | digestive | Cirrhosis of liver without mention of alcohol | 0.99 | -0.01 | 0.13 | -0.28 | 0.24 | 9.108E-01 | 2985 | 118580 | 121565 |
| 54 | infectious diseases | Herpes simplex | 0.99 | -0.01 | 0.16 | -0.34 | 0.28 | 9.125E-01 | 1991 | 118141 | 120132 |
| 452.2 | circulatory system | Deep vein thrombosis [DVT] | 1.01 | 0.01 | 0.11 | -0.22 | 0.22 | 9.146E-01 | 4735 | 115344 | 120079 |
| 714.1 | musculoskeletal | Rheumatoid arthritis | 0.99 | -0.01 | 0.13 | -0.28 | 0.23 | 9.149E-01 | 3374 | 117756 | 121130 |
| 200.1 | neoplasms | Polycythemia vera | 1.01 | 0.01 | 0.30 | -0.64 | 0.55 | 9.156E-01 | 639 | 121575 | 122214 |
| 191 | neoplasms | Malignant and unknown neoplasms of brain and nervous system | 1.01 | 0.01 | 0.30 | -0.64 | 0.55 | 9.158E-01 | 621 | 121307 | 121928 |

| | | | | | | | | | | | |
|---|---|---|---|---|---|---|---|---|---|---|---|
| 530.5 | digestive | Disorders of esophageal motility | 0.99 | -0.01 | 0.27 | -0.60 | 0.49 | 9.203E-01 | 759 | 120742 | 121501 |
| 440.22 | circulatory system | Atherosclerosis of native arteries of the extremities with intermittent claudication | 1.01 | 0.01 | 0.13 | -0.25 | 0.26 | 9.217E-01 | 3661 | 115529 | 119190 |
| 560.4 | digestive | Other intestinal obstruction | 0.99 | -0.01 | 0.16 | -0.35 | 0.29 | 9.236E-01 | 2254 | 118638 | 120892 |
| 260.3 | endocrine/metabolic | Adult failure to thrive | 0.99 | -0.01 | 0.18 | -0.40 | 0.33 | 9.244E-01 | 1881 | 118608 | 120489 |
| 604 | genitourinary | Disorders of penis | 1.01 | 0.01 | 0.15 | -0.30 | 0.29 | 9.268E-01 | 2795 | 114490 | 117285 |
| 196 | neoplasms | Radiotherapy | 0.99 | -0.01 | 0.16 | -0.35 | 0.29 | 9.268E-01 | 2278 | 119689 | 121967 |
| 288.1 | hematopoietic | Decreased white blood cell count | 1.01 | 0.01 | 0.16 | -0.32 | 0.32 | 9.312E-01 | 2095 | 118373 | 120468 |
| 252.1 | endocrine/metabolic | Hyperparathyroidism | 0.99 | -0.01 | 0.17 | -0.37 | 0.31 | 9.343E-01 | 1937 | 119537 | 121474 |
| 695.3 | dermatologic | Rosacea | 0.99 | -0.01 | 0.11 | -0.23 | 0.20 | 9.386E-01 | 5292 | 114141 | 119433 |
| 594.2 | genitourinary | Calculus of lower urinary tract | 1.00 | 0.00 | 0.22 | -0.46 | 0.40 | 9.434E-01 | 1364 | 120108 | 121472 |
| 728.71 | musculoskeletal | Contracture of palmar fascia [Dupuytren's disease] | 1.00 | 0.00 | 0.15 | -0.31 | 0.29 | 9.587E-01 | 2586 | 118574 | 121160 |
| 198 | neoplasms | Secondary malignant neoplasm | 1.00 | 0.00 | 0.10 | -0.21 | 0.19 | 9.617E-01 | 6042 | 114090 | 120132 |
| 696.4 | dermatologic | Psoriasis | 1.00 | 0.00 | 0.11 | -0.23 | 0.21 | 9.659E-01 | 4857 | 115549 | 120406 |

**Supplementary Table 4:** Laboratory association study comparing EUR FECD cases and controls.

| LAB VALUE | OR | CI 2.5% | CI 97.5% | p | n cases | n control | n Total |
|---|---|---|---|---|---|---|---|
| Bicarbonate | 1.15 | 1.100 | 1.203 | 7.31E-10 | 2131 | 111095 | 113226 |
| Hematocrit (HCT) | 0.90 | 0.855 | 0.936 | 1.57E-06 | 2128 | 111201 | 113329 |
| Hemoglobin A1c | 1.11 | 1.064 | 1.163 | 2.47E-06 | 2023 | 104639 | 106662 |
| Hemoglobin | 0.90 | 0.857 | 0.939 | 2.60E-06 | 2129 | 111205 | 113334 |
| Serum Glucose | 1.08 | 1.034 | 1.127 | 5.54E-04 | 2136 | 111660 | 113796 |
| Mean Corpuscular Hemoglobin (MCH) | 0.93 | 0.890 | 0.972 | 1.31E-03 | 2128 | 111186 | 113314 |
| Total Cholesterol | 0.93 | 0.894 | 0.977 | 2.65E-03 | 2123 | 110747 | 112870 |
| Red Blood Cell (RBC) | 0.93 | 0.891 | 0.977 | 2.90E-03 | 2128 | 111168 | 113296 |
| Mean Corpuscular Volume (MCV) | 0.94 | 0.895 | 0.978 | 3.23E-03 | 2128 | 111186 | 113314 |
| Uric Acid | 0.91 | 0.847 | 0.975 | 7.65E-03 | 820 | 42044 | 42864 |
| Low Density Lipoprotein Cholesterol (LDL-C) | 0.94 | 0.903 | 0.985 | 8.76E-03 | 2129 | 111033 | 113162 |
| Lymphocyte - Absolute Value | 0.94 | 0.896 | 0.988 | 1.41E-02 | 1743 | 83111 | 84854 |
| Ferritin | 0.93 | 0.869 | 0.987 | 1.81E-02 | 990 | 45067 | 46057 |
| Total Iron | 1.10 | 1.016 | 1.189 | 1.82E-02 | 663 | 29460 | 30123 |
| Serum Glucose - Finger Stick | 1.09 | 1.013 | 1.168 | 2.05E-02 | 787 | 37112 | 37899 |
| Eosinophil - Fractional Value | 1.05 | 1.006 | 1.102 | 2.52E-02 | 1987 | 102483 | 104470 |
| White Blood Cell (WBC) | 0.95 | 0.911 | 0.995 | 2.87E-02 | 2061 | 106491 | 108552 |
| Serum Chloride | 0.95 | 0.911 | 0.995 | 2.88E-02 | 2134 | 111434 | 113568 |
| Serum_Calcium (mg/dL) | 0.95 | 0.912 | 0.999 | 4.32E-02 | 2025 | 106430 | 108455 |
| Monocyte - Absolute Value | 0.96 | 0.910 | 1.003 | 6.56E-02 | 1774 | 84718 | 86492 |
| Prostate Specific Antigen (PSA) | 0.96 | 0.912 | 1.005 | 7.77E-02 | 1786 | 98732 | 100518 |
| Mean Corpuscular Hemoglobin Concentration (MCHC) | 0.96 | 0.922 | 1.006 | 9.39E-02 | 2128 | 111186 | 113314 |
| Basophil - Fractional Value | 0.96 | 0.905 | 1.008 | 9.41E-02 | 1499 | 74056 | 75555 |
| Aspartate aminotransferase (AST) | 0.96 | 0.922 | 1.007 | 9.82E-02 | 2070 | 108184 | 110254 |
| Serum Albumin | 0.96 | 0.921 | 1.008 | 1.05E-01 | 2074 | 106917 | 108991 |
| High Density Lipoprotein Cholesterol (HDL-C) | 0.97 | 0.925 | 1.010 | 1.34E-01 | 2127 | 110979 | 113106 |

| | | | | | | | |
|---|---|---|---|---|---|---|---|
| Serum Magnesium | 1.04 | 0.985 | 1.098 | 1.55E-01 | 1438 | 69647 | 71085 |
| Alanine Aminotransferase (ALT) | 1.03 | 0.987 | 1.079 | 1.67E-01 | 2124 | 111016 | 113140 |
| Troponin - T | 0.83 | 0.609 | 1.121 | 2.07E-01 | 50 | 2285 | 2335 |
| Iron | 0.96 | 0.899 | 1.025 | 2.20E-01 | 924 | 42301 | 43225 |
| Red Cell Distribution Width (RDW) | 1.03 | 0.983 | 1.075 | 2.22E-01 | 2077 | 108228 | 110305 |
| Total Creatine Kinase (CK) | 1.04 | 0.976 | 1.109 | 2.28E-01 | 1017 | 47157 | 48174 |
| Prothrombin Time (PT) | 0.96 | 0.897 | 1.030 | 2.57E-01 | 885 | 38217 | 39102 |
| Creatine Kinase MB (CKMB) - Fractional Value | 0.88 | 0.703 | 1.104 | 2.64E-01 | 82 | 3471 | 3553 |
| Creatine Kinase MB (CKMB) - Absolute Value | 1.06 | 0.958 | 1.167 | 2.66E-01 | 410 | 18320 | 18730 |
| Blood Urea Nitrogen (BUN) - Blood, Serum, Plasma | 1.03 | 0.979 | 1.073 | 2.88E-01 | 2133 | 111589 | 113722 |
| Eosinophil - Absolute Value | 1.02 | 0.975 | 1.071 | 3.60E-01 | 1965 | 98735 | 100700 |
| Transferrin Saturation (TSAT) | 0.97 | 0.893 | 1.048 | 4.11E-01 | 637 | 27558 | 28195 |
| C-Reactive Protein (CRP) (mg/L) | 0.94 | 0.820 | 1.086 | 4.13E-01 | 213 | 10831 | 11044 |
| Erythrocyte Sedimentation Rate (ESR) | 0.97 | 0.899 | 1.047 | 4.36E-01 | 749 | 31252 | 32001 |
| Mean Platelet Volume (MPV) | 1.02 | 0.972 | 1.066 | 4.46E-01 | 1961 | 102941 | 104902 |
| Neutrophil - Absolute Value | 0.98 | 0.938 | 1.031 | 4.78E-01 | 1810 | 90542 | 92352 |
| Serum Creatinine | 1.02 | 0.971 | 1.064 | 4.83E-01 | 2137 | 111719 | 113856 |
| Neutrophil - Fractional Value | 1.02 | 0.968 | 1.068 | 4.98E-01 | 1711 | 88352 | 90063 |
| Serum Potassium | 1.02 | 0.971 | 1.061 | 5.05E-01 | 2134 | 111622 | 113756 |
| Troponin | 0.93 | 0.688 | 1.233 | 5.79E-01 | 74 | 2989 | 3063 |
| N-Terminal Prohormone of Brain Natriuretic Peptide (proBNP) | 0.97 | 0.859 | 1.096 | 6.17E-01 | 297 | 12412 | 12709 |
| Troponin - I | 1.02 | 0.925 | 1.129 | 6.54E-01 | 412 | 20779 | 21191 |
| Basophil - Absolute Value | 1.01 | 0.947 | 1.086 | 6.78E-01 | 1178 | 53303 | 54481 |
| Estimated Glomerular Filtration Rate (eGFR) | 0.99 | 0.948 | 1.039 | 7.57E-01 | 2106 | 109972 | 112078 |
| Monocyte - Fractional Value | 1.00 | 0.959 | 1.054 | 8.31E-01 | 1856 | 92732 | 94588 |

| | | | | | | | |
|---|---|---|---|---|---|---|---|
| International Normalized Ratio (INR) | 0.99 | 0.943 | 1.050 | 8.39E-01 | 1507 | 70993 | 72500 |
| Platelets | 1.00 | 0.961 | 1.049 | 8.51E-01 | 2128 | 111185 | 113313 |
| Brain Natriuretic Peptide (BNP) | 0.99 | 0.883 | 1.115 | 8.65E-01 | 296 | 15162 | 15458 |
| C-Reactive Protein (CRP) (md/dl) | 1.00 | 0.869 | 1.143 | 8.81E-01 | 230 | 9096 | 9326 |
| Triglycerides | 1.00 | 0.954 | 1.042 | 8.82E-01 | 2120 | 110366 | 112486 |
| Lymphocyte - Fractional Value | 1.00 | 0.951 | 1.045 | 8.88E-01 | 1837 | 91204 | 93041 |
| Serum Sodium | 1.00 | 0.957 | 1.047 | 9.44E-01 | 2135 | 111618 | 113753 |
